# The Effectiveness and Use of Digital Technology Interventions for Upper Limb Rehabilitation at Home in Acute and Subacute Stroke: A Systematic Review and Meta-Analysis

**DOI:** 10.1101/2025.11.14.25340164

**Authors:** Helen J Gooch, James E Hill, Stephanie P Jones, Rachel C Stockley

## Abstract

**Objective:** This systematic review and meta-analysis aimed to evaluate the effectiveness of digital technology interventions for upper limb rehabilitation at home in the acute and subacute phases post-stroke.

**Data sources:** A multi-database search was conducted in Medline, EMBASE, PsycINFO, CINAHL, AMED, the Cochrane Library and Scopus up to 28^th^ April 2025.

**Review methods:** This review was informed by Cochrane guidelines. Studies were grouped according to the nature of the digital technology in the intervention. Data was synthesised using narrative text, tabulation and where possible meta-analysis. Risk of bias and certainty of evidence were evaluated. The review was registered on PROSPERO (CRD420251027928).

**Results:** From 3498 records identified, twelve studies (787 participants) were included. Four groups of digital technology interventions were identified: virtual reality (n=7), electrical stimulation (n=2), wearable sensors (n=2), and mobile applications (n=1). Meta-analysis comparing virtual reality with time matched conventional exercise suggested no evidence of effect on either upper limb motor impairment (n=3, 241 participants, mean difference= - 0.23, 95% confidence interval (CI):-2.58 to 2.12) or activity limitation (n=5, 467 participants, standardised mean difference= 0.07, 95% CI:-0.25 to 0.38) with low certainty of evidence.

**Conclusion:** Evidence of the effectiveness of digital technology interventions for upper limb rehabilitation at home in the acute and subacute phases following stroke is limited. Further research is needed to understand if and how virtual reality interventions could provide an alternative therapy option. No summary conclusions could be drawn on the use of the other digital technologies identified in the review.

## Introduction

Management of disability following stroke presents a major challenge for global healthcare, with 12.2 million people worldwide experiencing a stroke each year.^1^ Upper limb motor impairments are reported by around three-quarters of stroke survivors,^2^ resulting in limitations of functional activity,^3–5^ and substantial impact on the lives of those affected.^6,7^

The amount of motor recovery is strongly associated with the use of the upper limb in everyday activities,^8–10^ therefore maximising this recovery is a key focus of rehabilitation.^11^ The potential for upper limb motor recovery is greatest within the acute and subacute phases (first six months) following a stroke,^3,12–15^ thus providing a temporal opportunity for enhancing rehabilitation impact. In order to promote motor recovery and functional activity, guidelines indicate that upper limb rehabilitation should be repetitive and task focused,^11,16^ and although the optimal dose remains unclear, higher doses are considered to facilitate greater recovery.^17,18^

Digital technologies are increasingly being explored as a means to enhance post-stroke rehabilitation provision. There are promising findings for the use of digital technologies to address motor impairment and activity limitation, demonstrated by meta-analyses, for a variety of digital technologies, including virtual reality,^19–23^ robotics,^24–26^ digital games,^27^ electrical stimulation, ^28–30^ and telerehabilitation.^31^ This current evidence indicates comparable effect, and at times greater effect, to usual care. Similarly, comparable and at times favourable effects are also suggested in the acute and subacute phases after stroke.^19,21,22,25–28,30^

Understanding how digital technologies can be utilised for acute and subacute rehabilitation within the United Kingdom (UK), where current drivers are moving the delivery of care in or closer to home through early supported discharge and community rehabilitation,^11,16,32–34^ needs further consideration. As the length of hospital stay decreases,^35^ stroke survivors are returning home with significant levels of disability and rehabilitation needs; this, coupled with reduced frequency of formal therapy sessions following discharge home,^36^ raises challenges around achieving sufficient dose of rehabilitation in the home environment.

The use of digital technologies in home-based upper limb rehabilitation is less well appraised than their use across a variety of settings. A recent scoping review has indicated the types of digital technologies used in the home are games, virtual reality, sensors, tablets, robotics and telerehabilitation.^37^ Literature focused on evaluating the effect of home-based digital technologies,^31,38–41^ indicates lower numbers of primary studies posing challenges for meta-analysis. Despite this there is indication of comparable effect, and at times greater effect, to usual care in line with the broader body of evidence. This literature however lacks any focus or investigation within the acute and subacute phases following stroke.

This systematic review focused on the use of digital technology interventions for upper limb rehabilitation at home in the acute and subacute phases following stroke. The objectives were to (1) determine the effectiveness of a range of digital technology interventions delivered in the home environment, that target motor impairment and activity limitation of the upper limb, in the acute and subacute phases (first six months) following stroke; (2) examine if the effectiveness of these interventions was influenced by the severity of the stroke participants or how much of the intervention was delivered; (3) identify if economic evaluation had been conducted for any of the interventions, and if available, describe the findings.

## Methods

This systematic review was informed by the current Cochrane guidelines,^42^ and reported with consideration of the Preferred Reporting Items for Systematic reviews and Meta-Analyses (PRISMA) guidelines.^43^ The review protocol was prospectively registered on PROSPERO (CRD420251027928).

### Eligibility Criteria

Publications and peer reviewed theses that reported the results of a randomised controlled trial (RCT) that met the Population, Intervention, Comparator, Outcome (PICO) criteria,^44^ were included in the review (Table 1).

**Table 1:** Inclusion criteria.

| PICO | Criteria |
| --- | --- |
| Population | Adults who have experienced a stroke within the last six months with upper limb involvement |
| Intervention | Digital technology rehabilitation intervention delivered in the home setting to target the upper limb |
| Comparator | Usual or conventional therapy, no therapy or a defined placebo intervention |
| Outcomes | In the context of the International Classification of Functioning, Disability and Health,<sup>45</sup> upper limb motor impairment (domain B7) and/or activity limitation (domain D4) |

Conference abstracts, protocols and trial registry entries without available published results, non-randomised trials and publications from trials where data from eligible stroke participants could not be ascertained were excluded. Trials in which a digital technology intervention was used for a single treatment session, was delivered to a group or delivered in an unknown location, and those in which the comparator group utilised a digital technology were not included. The search was limited to English language, due to study resources, and to publications from 2000, reflecting the main period of advancement in the use of digital technologies.^46^ Further detail on the eligibility criteria is available in Supplemental Table S1.

### Information Sources, Search Strategy and Selection Process

A multi-database search, developed in consultation with an information specialist, was conducted in Medline, EMBASE, PsycINFO, CINAHL, AMED, the Cochrane Library and Scopus. The search used a combination of keywords, and controlled vocabulary related to six distinct search streams related to the population (1. Stroke 2. Upper limb), the intervention (3. Technology, 4. Rehabilitation, 5. Home-based), and the study design (6. Randomised trials) which were then combined. Clinical trials registries (Clinical Trials, The UK Clinical Study Registry (ISRCTN), and the International Clinical Trials Registry Platform (ICTRP)) and ProQuest health and medicine databases (limited to dissertations and theses) were also searched. The final search strategy (Supplemental Table S2) was performed from the date of inception to the 28^th^ and 29^th^ April 2025.

The database and registry searches were collated and deduplicated in EndNote 21 software^47^ using a structured process.^48^ Title and abstract screening and then full text assessment was completed by two independent reviewers, facilitated by Rayyan software.^49^ Disagreements were resolved by discussion. Reasons for exclusion at full text assessment were recorded. Study publications were sought from potentially eligible conference abstracts, protocols and trial registry entries through online searching and email contact with authors.

Additional studies were identified from the full text searching process, from reference lists of relevant reviews,^37–39,41^ and all included studies, and through citation searching of included studies (completed on Scopus between 13^th^ June and 23^rd^ June 2025). Full texts were assessed by two independent reviewers and disagreements resolved by discussion.

### Data Collection Process and Data Items

Data for the included studies was extracted onto a custom made, piloted, data collection tool (Supplemental Table S3) created in Microsoft Excel software.^50^

Data was collected on study design, participants (including the PROGRESS framework equity factors),^51^ interventions, outcomes, and results. The intervention data was structured on the Template for Intervention Description and Replication: Extension for Rehabilitation (TIDieR-Rehab),^52,53^ with consideration of the Dose Articulation Framework.^54^ Outcome data for motor impairment and activity limitation was collected for all valid measurement tools, time points and analyses. Data was also collected for all valid tools measuring harmful effects, reporting of adverse events, intervention fidelity and economic evaluation.

One reviewer collected data from all the included studies, with a second reviewer independently collecting data for 40% of studies to enable cross checking for interpretation and accuracy, and all of the outcome data included in the meta-analysis to cross check accuracy. Any conflicts were resolved by review of the publications and discussion. An email request for further data on one study was made without success.

### Study Risk of Bias Assessment

In line with current Cochrane guidance, the revised Cochrane Risk of Bias Tool (RoB 2) was used,^55,56^ and available resources facilitated the assessment.^57^ For the studies included in the meta-analyses, the RoB 2 tool was completed for each outcome domain and specific time point included in the meta-analyses. For studies not included in the meta-analyses, a pragmatic decision was made to complete the RoB 2 for the domain of activity limitation. The assessment was completed independently by two reviewers with disagreements resolved by discussion.

### Synthesis Methods

The study information, participant characteristics, intervention characteristics, outcome measures and reported findings are summarised using narrative text, descriptive statistics (facilitated by IBM SPSS Statistics)^58^ and tabulation.

The statistical synthesis of the outcome domains of motor impairment and activity limitation were planned with consideration of the detail of the digital technology used in the intervention, the type of comparator intervention and the available outcome data. Studies were grouped after data collection according to the type of digital technology in the intervention, how it was being used and what the target of the intervention was. Each digital technology group was further split into studies with an active comparator intervention and studies with a usual care comparator. For each study a single outcome measurement tool, for each of the two outcome domains, was selected for the meta-analysis after data extraction. When possible the core outcome set recommendation was used (motor impairment: Fugel-Meyer Assessment for Upper Extremity, activity limitation: Action Research Arm Test)^59,60^ and if this was not reported, the measure considered most similar to the core outcome from the available measures was used. All measures included in the meta-analyses were assessor observed measures and not patient reported measures, and were collected directly post intervention or within the first month following the intervention, if post intervention data were not available. Only studies reporting mean data were included. Studies were included irrespective of the finding of the RoB 2 assessment.

### Meta-analysis

Computations were completed using Comprehensive Meta-Analysis Version 4.^61^ The meta-analyses were based on the effect of the assignment and used an inverse-variance with a random-effects model.^62–67^ The calculated effect size used was mean difference (MD) when all included studies used one outcome measurement tool and standardised mean difference (SMD) when different outcome measurement tools associated with the same underlying construct were used. A 95% confidence interval (CI) was calculated and the null hypothesis was tested using a criterion alpha value of 0.05. Heterogeneity was assessed through visual inspection of forest plots and calculation of the I² statistic.^64,68–71^

In line with Cochrane guidelines, if the mean was reported alongside a CI rather than the standard deviation (SD), then CI was used to calculate the SD.^72^ In addition for the meta-analysis of MDs, in the absence of post intervention data, the change from baseline data with its SD was used without the need for data conversion.^64^

The minimal clinically important difference was identified for relevant outcome data. To facilitate interpretation, a pooled SMD was re-expressed by multiplying the pooled SMD and its 95% confidence interval by a representative SD of the minimal clinically important difference, following Cochrane recommendations.^73^

Subgroup analyses exploring the severity of the stroke participants or how much of the intervention was delivered were planned to be conducted only if there were sufficient studies to do so.^64^ Meta-analysis data and findings are presented as narrative text, tables and forest plots.

### Reporting Bias Assessment

Selective non-publication was explored broadly for the whole systematic review through identification of potentially eligible studies in trial registries for which no publication could be found or for which the review authors had no confirmation that the study was still in progress. Funnel plots were produced and visually inspected for the studies included in each meta-analysis.

### Certainty Assessment

The Grading of Recommendations Assessment, Development, and Evaluation (GRADE) tool was completed to support interpretation of the review results.^74,75^ It was completed independently by two reviewers with disagreements resolved by discussion.

## Results

### Study Selection

The database and registry search yielded 3498 records. After deduplication, 2397 records were screened by title and abstract. From these records and those identified by other methods, 267 full texts were assessed and twelve studies included in the review.^76–87^ (Figure 1). The most frequent reasons for exclusion at full text assessment were that participants were more than six months post-stroke (106/255, 42%) and that the intervention was not conducted in the home environment (48/255, 19%). Six studies had additional publications associated with the main study publication including protocols and results from sub-studies.^88–95^

**Figure 1:**
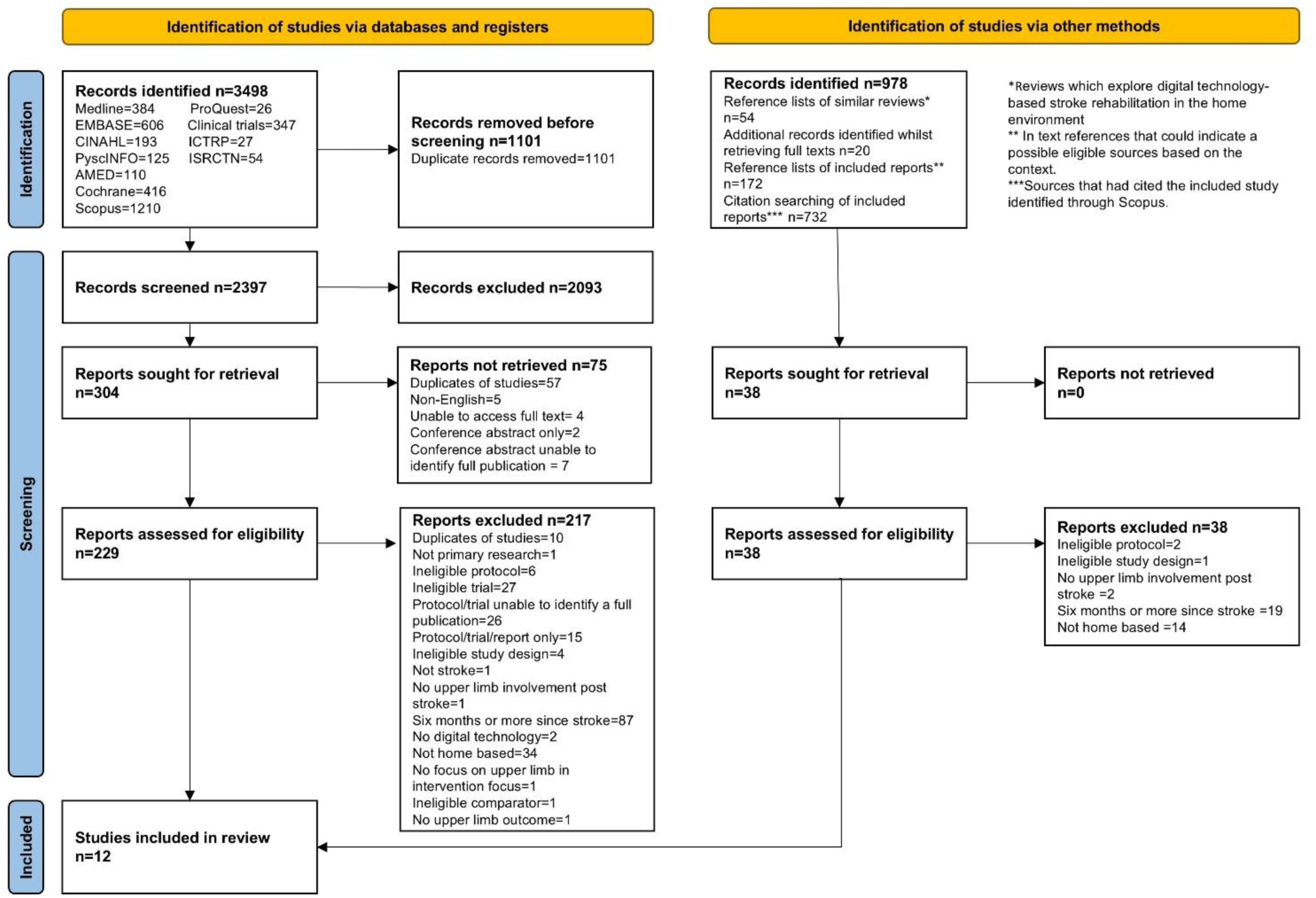
Preferred Reporting Items for Systematic Reviews and Meta-analyses (PRISMA) flowchart

### Study Characteristics

A summary of the included studies is provided in Table 2.

**Table 2:** Overview of included studies.

| Study | Participants |  |  |  | Intervention |  |  |  | Outcomes |  | Follow up period |
| --- | --- | --- | --- | --- | --- | --- | --- | --- | --- | --- | --- |
|  | Number n | Gender Female n (%) | Age (years) Mean (SD) *Median (IQR) | Limitation in upper limb activity Mean (SD) (or CI if stated) *Median (IQR) | Experimental | Comparator | Time-matched | Length | Upper limb impairment | Upper limb activity |  |
| Adie <sup>76</sup> | 235<br>E=117<br>C=118 | E=51 (44%)<br>C=53 (45%) | E=66.8 (14.6)<br>C=68.0 (11.9) | ARAT<br>E=41.2 (15.9)<br>C=41.0 (16.6) | Virtual reality (Wii™ games) | Exercise programme | Yes | 6 weeks | None | ARAT (MAL only at 6 months) | 6 months |
| Alon <sup>77</sup> | 15<br>E=7<br>C=8 | E=4 (57%)<br>C=3 (37.5%) | Not reported | BBT<br>E=5.9 (6.0)<br>C=5.3 (6.2) | Electrical stimulation and exercise programme | Exercise programme | No | 12 weeks | modified FMA-UE | BBT JHFT | None |
| Butcher <sup>78</sup> | 24<br>E=16<br>C=8 | E=13 (81%)<br>C=4 (50%) | E=66.5 (15)<br>C=64.6 (13.6) | ARAT<br>E=29.5 (21.0)<br>C=36.1 (22.7) | Virtual reality (Neurofenix platform) | Control (usual care including therapy) | No | 7 weeks | FMA-UE | ARAT MAL | None |
| Cramer <sup>79</sup> | 124<br>E=62<br>C=62 | E=14 (23%)<br>C=20 (32%) | E=62 (14)<br>C=60 (13) | BBT<br>E=21.3 (13.3)<br>C=23.8 (12.7) | Virtual reality , onscreen exercises and education, videoconferencing for supervised sessions | In-clinic supervised exercise and training and home exercise programme | Yes | 6 to 8 weeks | FMA-UE | BBT SIS (hand) | 30 days post intervention † |
| Da-Silva <sup>80</sup> | 33<br>E=14<br>C=19 | E=8 (57%)<br>C=12 (63%) | *E=73 (65–80)<br>*C=69 (61–80) | ARAT<br>*E=37 (16-46)<br>*C=15 (2-35) | Wearable sensor (CueS) and activity programme | Sham device and activity programme | Yes | 4 weeks | MI | ARAT MAL | 8 weeks |
| Emmerson <sup>81</sup> | 62<br>E=30<br>C=32 | E=13 (43%)<br>C=13 (41%) | E=68 (15)<br>C=63 (18) | WMFT (time in seconds)<br>E=39 (44)<br>C=49 (47) | Mobile application iPad exercise programme and reminders | Exercise programme | Yes | 4 weeks | Grip strength | WMFT | None |
| Fletcher-Smith <sup>82</sup> | 40<br>E=20<br>C=20 | E=13 (65%)<br>C=7 (35%) | *E=75 (66.5-80)<br>*E=67 (59-84) | ARAT<br>*E=0 (0-6)<br>*C=0 (0-5.5) | Electrical stimulation | Control (usual care including therapy) | No | 3 months | NIHSS arm | ARAT | 6 and 12 months |
| Standen <sup>83</sup> | 27<br>E=17<br>C=10 | E=9 (53%)<br>C=2 (20%) | E=59 (12.03)<br>C=63 (14.06) | WMFT (time in seconds)<br>*E=2.60 (1.65-6.00)<br>*C=3.34 (1.90-4.92) | Virtual reality (Virtual glove) | Control (usual care, no intensive therapy) | No | 8 weeks | Grip strength | WMFT NHPT MAL | None |
| Swanson <sup>84</sup> | 27<br>E=14 | E=0 (0%)<br>C=4 (31%) | E=50.3 (10.9)<br>C=52 (8.7) | BBT<br>E=25.4 (17.6) | Virtual reality (FitMi) | Exercise programme | Yes | 3 weeks | FMA-UE | BBT MAL | 1 month post |
|  | C=13 |  |  | C=23.5 (14.8) |  |  |  |  |  |  | intervention † |
| Wei <sup>85</sup> | 84<br>E=32<br>C1=25<br>C2=27 | E=7 (22%)<br>C1=5 (20%)<br>C2=4 (15%) | E=59.19 (11.25)<br>C1=60.44 (10.38)<br>C2=63.11 (10.27) | ARAT<br>E=34.00 (17.84)<br>C1=36.12 (21.84)<br>C2=35.96 (22.59) | Wearable sensor<br>(Remind to Move)<br>and exercise<br>programme | C1= Sham<br>device and<br>exercise<br>programme<br>C2=Control<br>(usual care<br>including<br>therapy) | No | 4 weeks | FMA-UE | ARAT<br>BBT<br>MAL | 8 and 12<br>weeks |
| Wilson <sup>86</sup> | 17<br>E=10<br>C=7 | E=3 (30%)<br>C=2 (29%) | E=69.9 (13.8)<br>C=77.3 (8.9) | BBT<br>E=23.5 (9.6)<br>C=15.7 (10.8) | Virtual reality<br>(EDNA) | Exercise<br>programme | Yes | 8 weeks | None | BBT<br>NHPT | 3<br>months<br>post<br>intervention † |
| Wolf <sup>87</sup> | 99<br>E=51<br>C=48 | E=26 (51%)<br>C=17 (35%) | E=59.1 (14.1)<br>C=54.7 (12.2) | ARAT<br>E=34.4 (95% CI:<br>24.7, 44.0)<br>C=31.1 (95% CI:<br>22.1,40.1) | Virtual reality using<br>a robotic device<br>(Hand Mentor Pro)<br>and exercise<br>programme | Exercise<br>programme | Yes | 8 to 12<br>weeks | FMA-UE | ARAT<br>WMFT<br>SIS (hand) | None |

**Abbreviations**
C: Comparator group, CI: Confidence Interval, E: Experimental group, FMA-UE: Fugl-Meyer Assessment for Upper Extremity, IQR: Interquartile Range, JHFT: Jebsen-Taylor Hand Function Test, MAL: Motor Activity Log, MI: Motricity Index, n: Number, NHPT: Nine Hole Peg Test, NIHSS: National Institute of Health Stroke Scale, SD: Standard Deviation, SIS: Stroke Impact Scale, WMFT: Wolf Motor Function Test
\*median and IQR reported, †follow up time measured from end of intervention

### Study design and participant characteristics

All twelve studies used a parallel design with two^76–84,86,87^ or three^85^ groups. Five studies (5/12, 42%) were feasibility/pilot trials,^77,78,80,82,83^ five (5/12, 42%) were definitive trials,^76,79,84,85,87^; the study type of the remaining two (2/12, 17%) was unclear.^81,86^ Seven studies (7/12, 58%) were multisite studies,^76,78–81,85,87^ with between two and eleven sites included. Five studies (5/12, 42%) were conducted in the UK,^76,78,80,82,83^ four (4/12, 33%) in the United States of America,^77,79,84,87^ two (2/12, 17%) in Australia,^81,86^ and one (1/12, 8%) in China.^85^

The median study sample size across the twelve studies was 36.5 (range 15 to 235) with a total of 787 participants, (307 females, 39% and 480 males, 61%). Participants’ ages were reported for all studies except for one,^77^ and ranged between a mean of 50.3 and 77.3 years. The average age was less than 70 years in eight studies (8/11, 73%).^76,78,79,81,83–85,87^ Baseline upper limb motor impairment was reported in nine studies (9/12, 75%),^77–82,84,85,87^ using a range of different measures with over half (5/9, 56%)^78,79,84,85,87^ reporting the Fugel-Meyer

Assessment for Upper Extremity. Values ranged between a mean of 33.3 and 52.0. All studies reported a baseline measure of upper limb activity limitation. Six studies (6/12, 50%) reported the Action Research Arm Test,^76,78,80,82,85,87^ and values ranged between a median of zero and a mean of 41.2. A summary of participant characteristics for each study is available in Supplemental Table S4.

The eight PROGRESS equity factors^51^ were poorly reported, with each factor being reported by a median of one study (range 0 to 12) (Supplemental Table S5). Gender was reported in all twelve studies. Only one study (1/12, 8%) reported employment data,^76^ and no studies reported on socioeconomic status.

### Intervention characteristics

The intervention characteristics of the digital technology intervention, reported against the TIDieR-Rehab checklist (excluding those relating to study findings), are summarised in Tables 3a and 3b.

**Table 3a:**
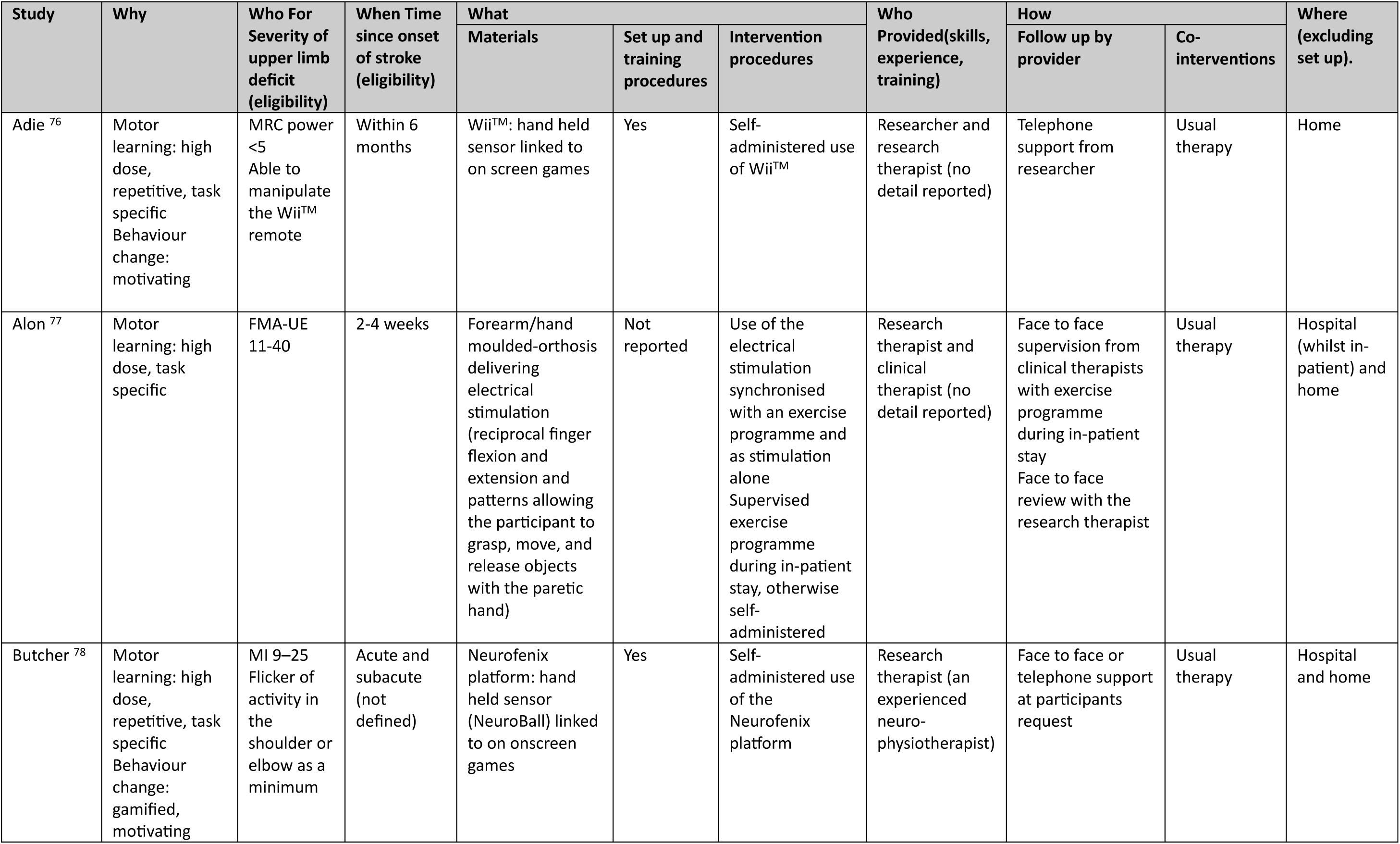
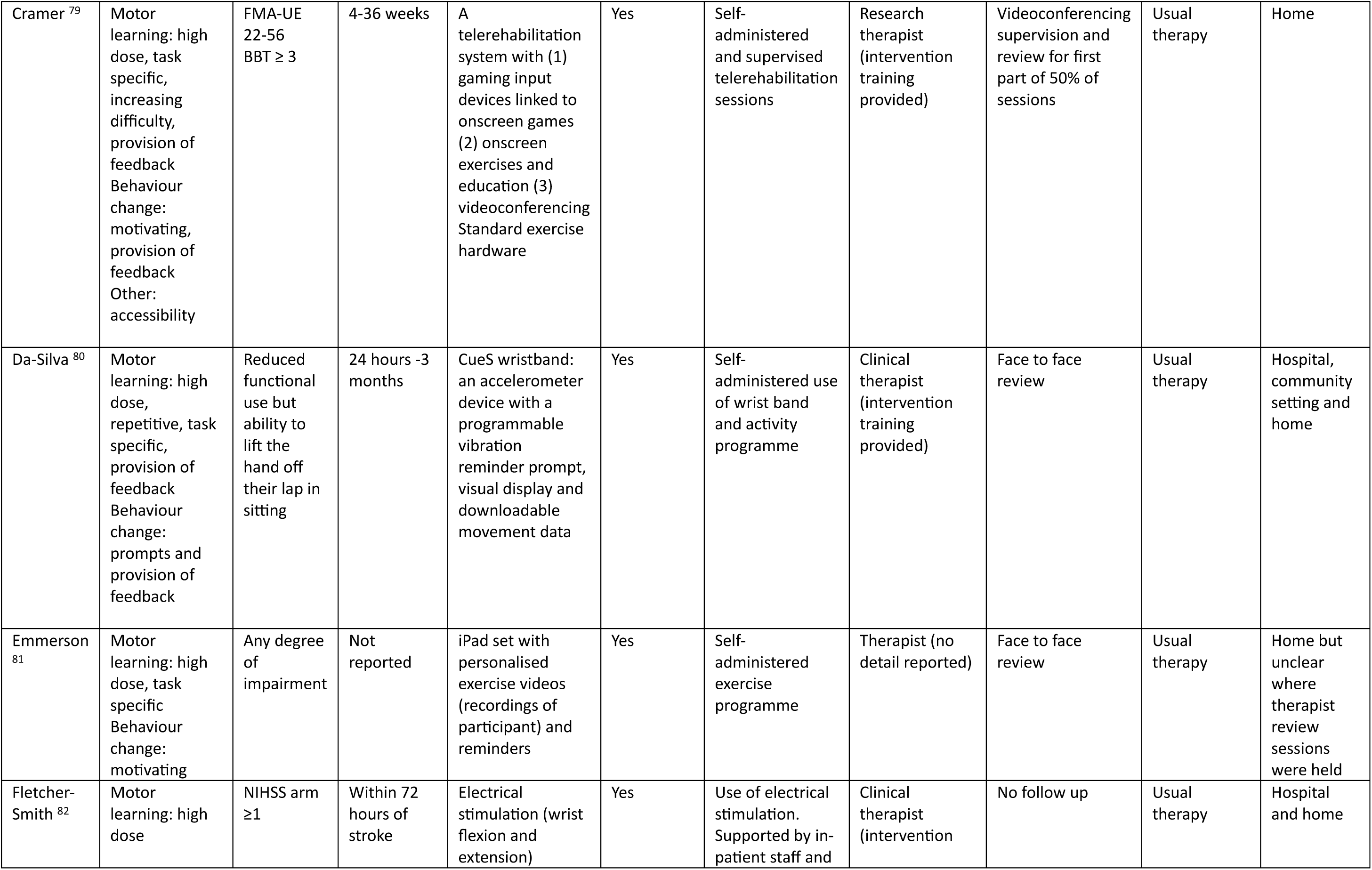
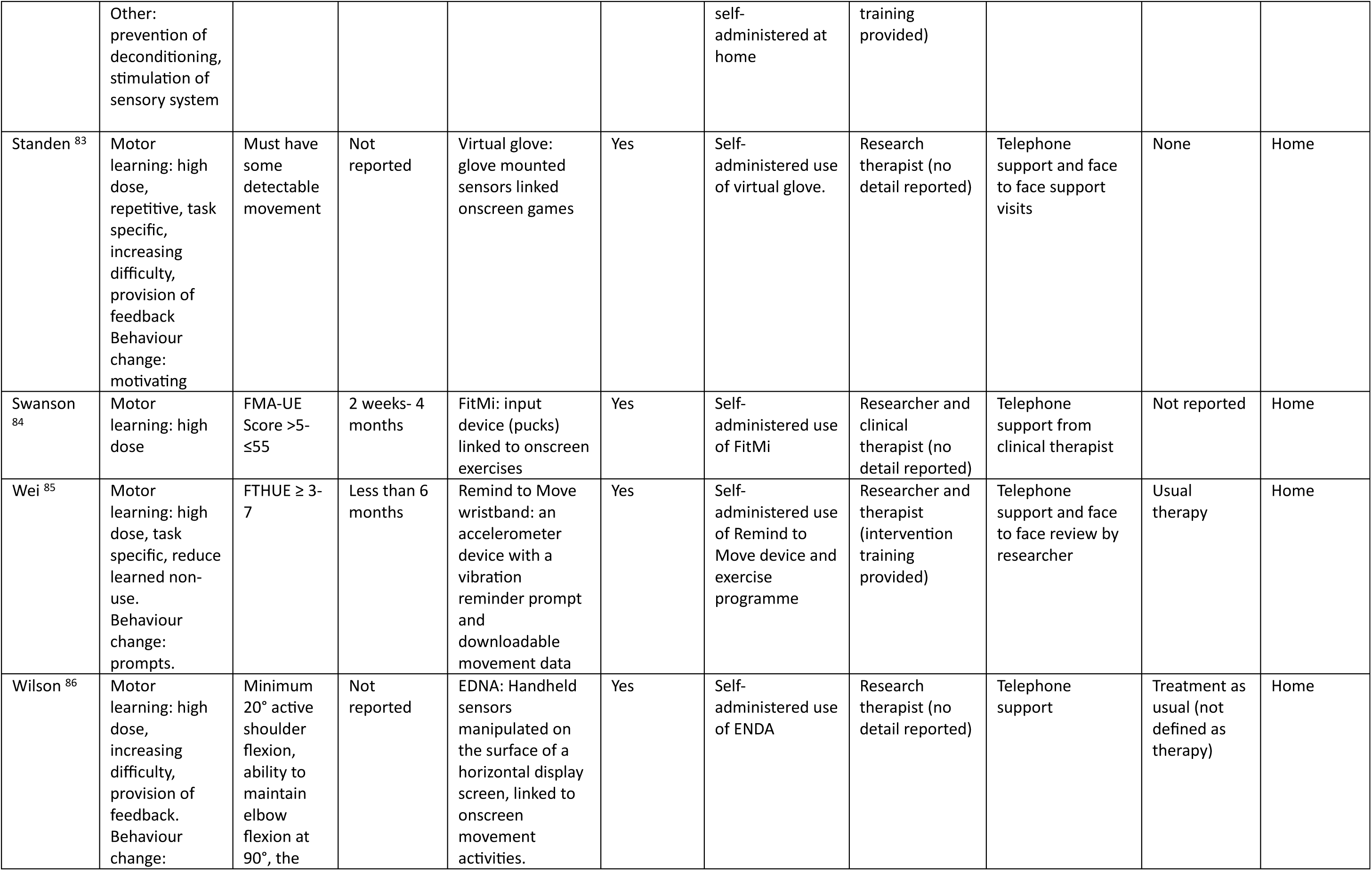
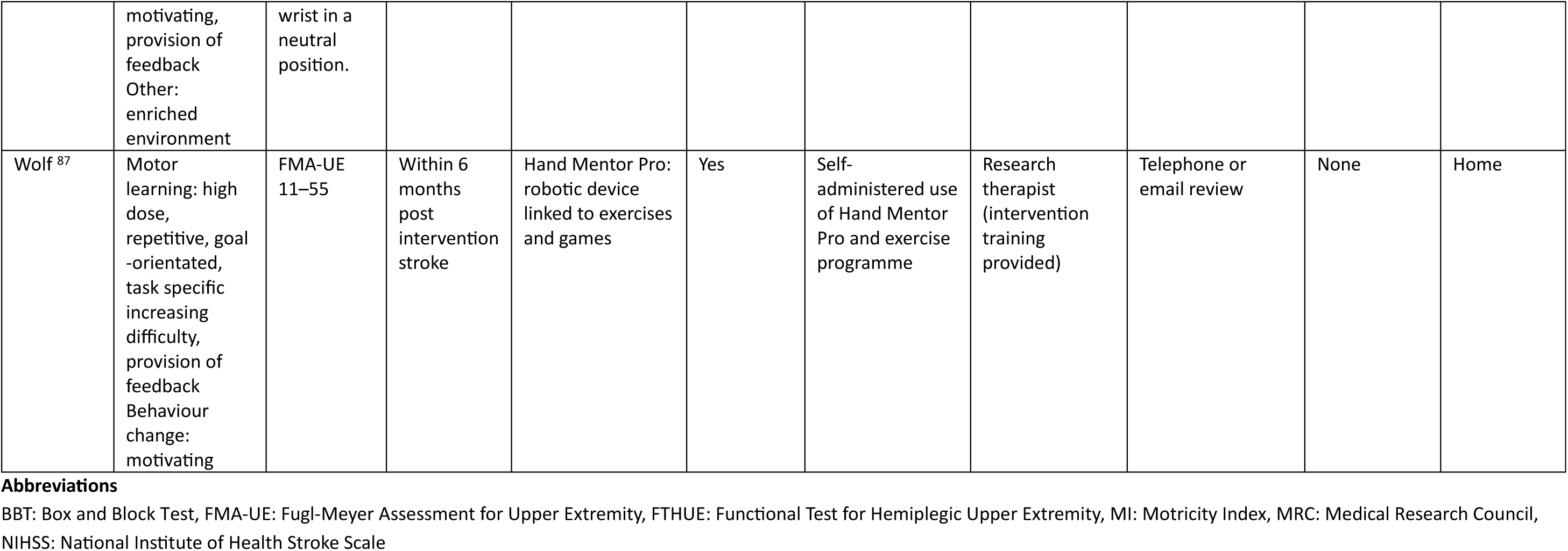
TIDieR-Rehab items (why, who for, when, what, who provided, how, where) for included studies.

**Table 3b:**
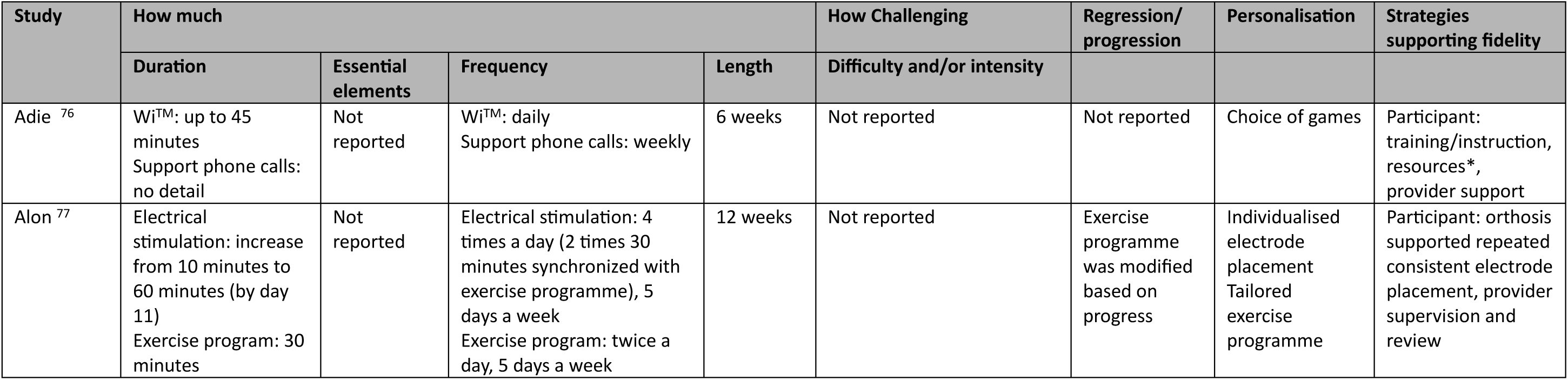
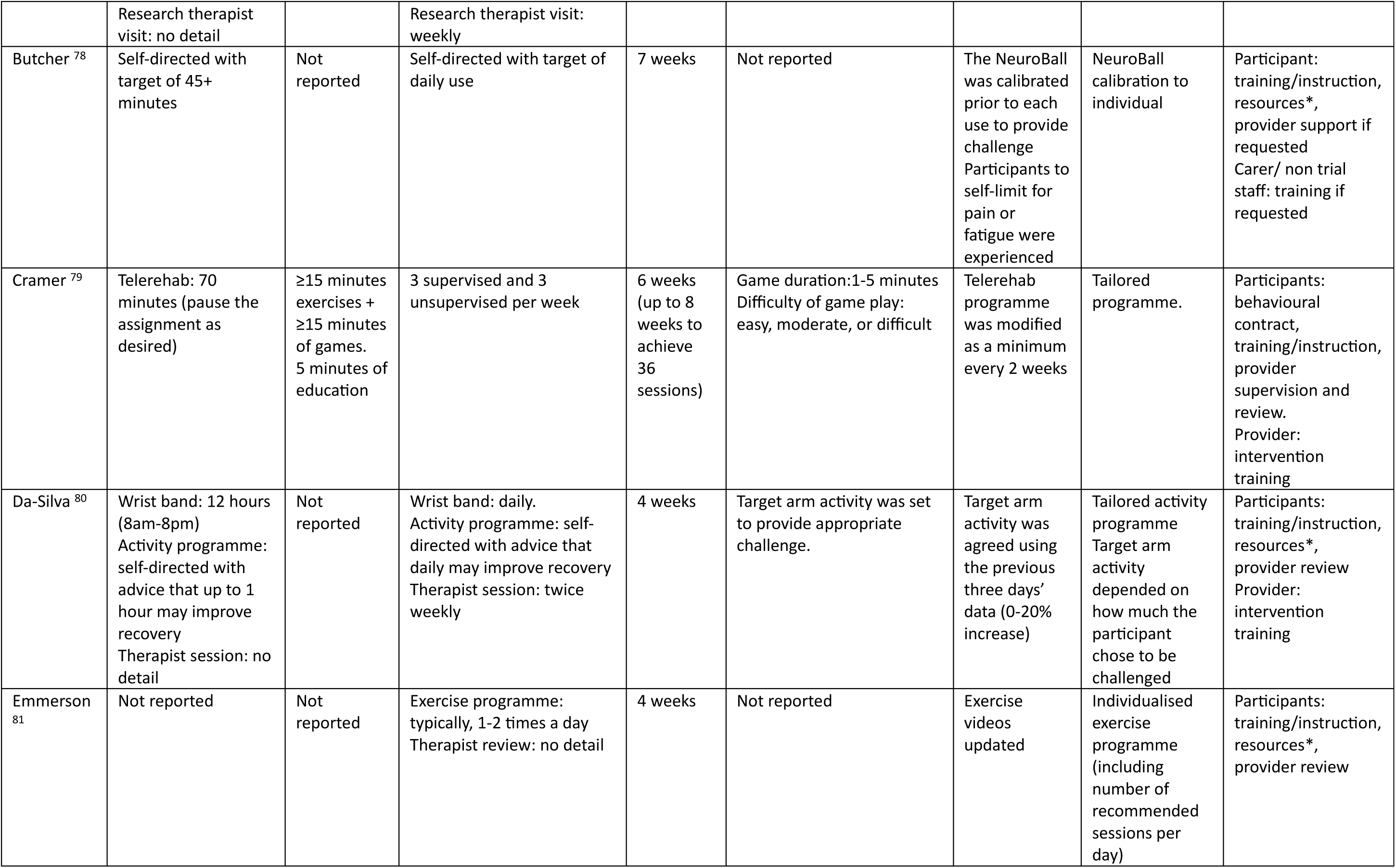
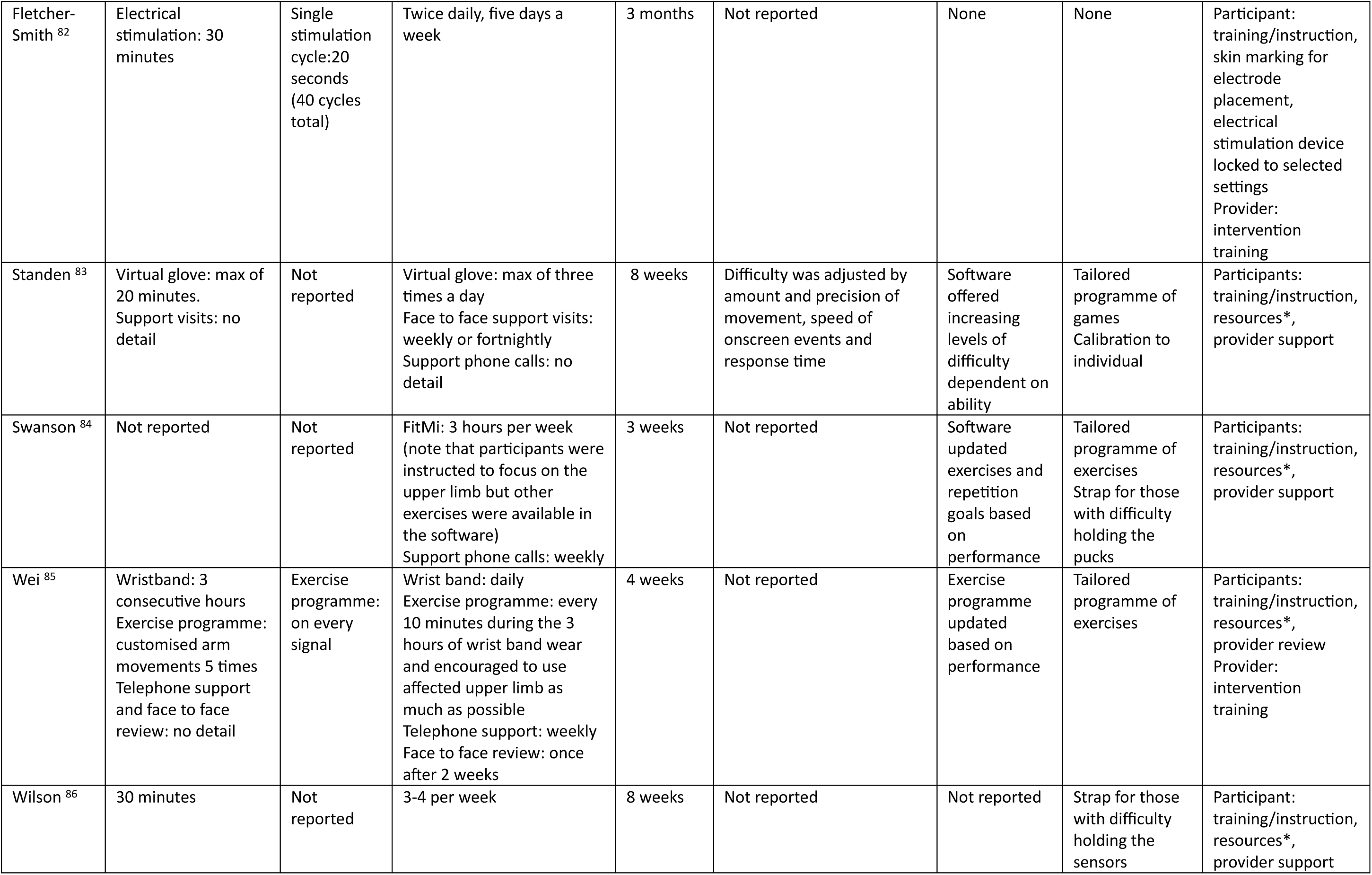
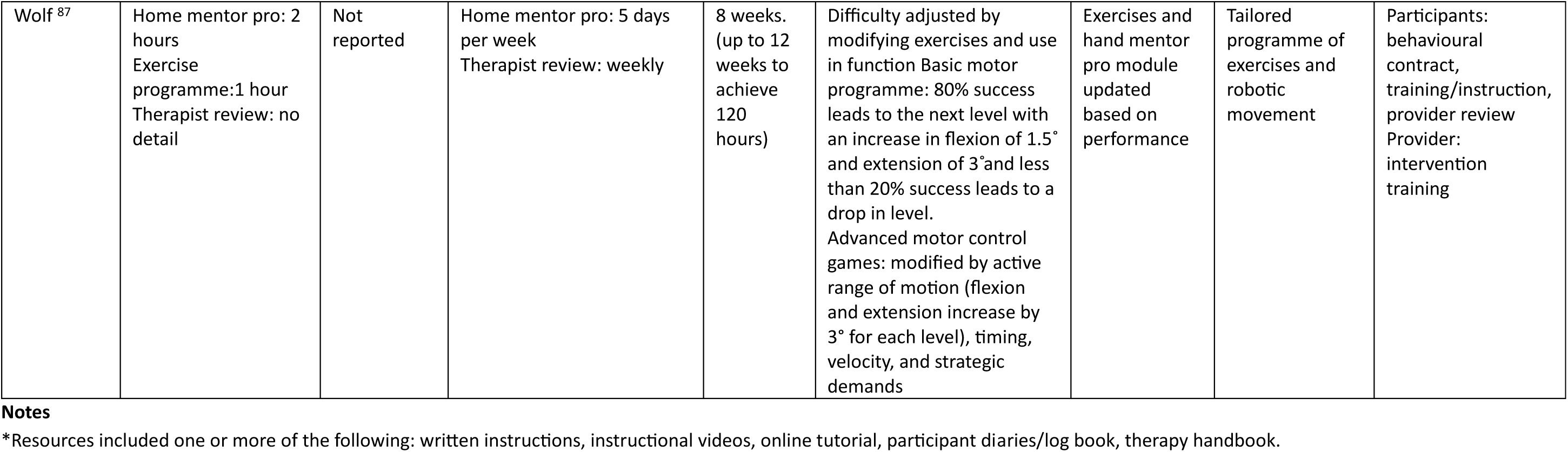
TIDieR-Rehab items (how much, how challenging, progression, personalisation, fidelity strategies) for included studies.

Relevant data were available from the majority of studies for ten of the twelve included TIDieR checklist items. Detail on the essential elements of the intervention were missing in nine studies (9/12, 75%),^76–78,80,81,83,84,86,87^ and detail on intervention challenge was missing in eight studies (8/12, 67%).^77,78,81,82,84–86,88^

### TIDieR item: Who for

All the interventions were designed to be delivered to stroke survivors with a range of severity of upper limb involvement with inclusion criteria based on a variety of different outcome measures as well as informal observation of activity. The majority of studies (10/12, 83%) excluded those with the most severe deficit.^76–80,83–87^

### TIDieR item: What

From the included studies the nature of the digital technology used within the experimental interventions was divided into four groups. Seven studies (7/12, 58%) used upper limb motion capture to interact with a digital screen in order to complete activities or game play,^76,78,79,83,84,86,87^ either as a complete intervention,^76,78,83,84,86^ or as part of an intervention.^79,87^ This group of interventions will hereinafter be referred to as virtual reality with acknowledgment that one study also incorporated videoconferencing and onscreen exercises and education,^79^ and one study incorporated a robotic device and exercise programme,^87^ into the intervention alongside the virtual reality component.

Two studies (2/12, 17%) used peripheral neuromuscular electrical stimulation,^77,82^ (hereafter referred to as electrical stimulation) either standalone,^82^ or as a combination of functional electrical stimulation (electrical stimulation synchronised with functional exercise) and electrical stimulation alone.^77^ Two studies (2/12, 17%) used wrist worn wearable sensor devices,^80,85^ (hereafter referred to as wearable sensors) which were able to collect movement data and provide a vibration cue for movement. The final study (1/12, 8%) used a mobile device to record a personalised exercise programmes and provide exercise reminders,^81^ (hereafter referred to as mobile applications).

The comparator groups included an active conventional exercise or activity programme for three-quarters of the studies (9/12 75%),^76,77,79–81,84–87^ with seven (7/9, 78%) time matched to the experimental group.^76,79–81,84,86,87^ Usual therapy was provided to all participants in three quarters of studies (9/12, 75%).^76–82,85,86^

### TIDieR item: How much

Detail of the planned dose was indicated through measures of time, i.e., session duration and frequency and overall length of the intervention, and varied within and between the digital technology groups. For the seven studies grouped as virtual reality,^76,78,79,83,84,86,87^ the planned intervention time per week in minutes varied from 120 to 900 with a median of 315 (Interquartile Range (IQR): 180-420). Interventions continued for between three and eight weeks (median:7 (IQR: 6-8)). The minutes of planned intervention per day (calculated from the available data as the total minutes of intervention divided by the total length of the intervention in days) ranged from 17 to 129 with a median of 45 minutes per day (IQR: 26-60)). The two electrical stimulation studies planned to deliver either 300 minutes,^82^ or 1200 minutes,^77^ per week, over a twelve-week period and the minutes of planned intervention per day (calculated as above) were 43 and 171 respectively. For the two wearable sensor studies, the device was planned to be worn for 1260 minutes,^85^ and 5040 minutes,^80^ per week, over a four-week period, with the minutes of planned wear per day calculated as 180 and 720 respectively. In the mobile application study,^81^ the intervention time per week was not reported over the four week intervention.

All seven studies looking at virtual reality also reported the actual time-based dose completed in a variety of formats (Table 4). Four studies (4/7, 57%) used data from the digital technology itself,^78,83,84,87^ three studies (3/7, 43%) used therapist and participant reporting,^79,87,88^ and one study (1/7, 14%) lacked detail of how data was collected.^86^ Generally, the time spent on the intervention was lower than the planned time, although one study^87^ reported a higher actual time compared with planned time. Only one study ^84^ reported a broader dimension of dose, in the form of repetitions, and although they identified a lower actual intervention time, they found that participants completed more repetitions than the theoretical target set for the intervention. One of the studies looking at electrical stimulation,^82^ one of the studies using wearable sensors,^80^ and the mobile application study,^81^ also reported that the actual intervention time was lower than the planned time, with only one study using the digital technology to collect this data. ^80^

**Table 4:** Actual dose versus planned dose.

| Technology group | Study | How data was collected | Actual dose | Planned dose |
| --- | --- | --- | --- | --- |
| Virtual reality | Adie <sup>76</sup> | Participant | Mean daily time: 37 minutes (SD:16.2) | Daily time: 45 minutes |
|  |  |  | Mean total time: 1020.2 minutes (SD:721) | Total time: 1890 minutes |
|  | Butcher <sup>78</sup> | Technology | Median total time: 11 (range 1–58) hours | Total time: 36.75 hours |
|  |  |  | Mean total days: 22.4 (SD:14.4) (range: 3–48 ) | Total days: 49 |
|  |  |  | Mean days trained per week: 3.2 (SD:1.9). | Days per week: 7 |
|  | Cramer <sup>79</sup> | Participant / therapist | Percentage of the 36 therapy sessions for which the patient completed 40 or more of the assigned 70 minutes: 98.3% (35.4/36) | Sessions and time: 36 sessions lasting 70 minutes |
|  | Standen <sup>83</sup> | Technology | Range of the percentage of recommended time of use (whilst device was in the participants home*): 1.46% - 70.6%. | Total time: 3360 minutes |
|  |  |  | Range of percentage of recommended days used (whilst device was in the participants home*): 10% - 100%. | Total days: 56 |
|  |  |  | Daily time (on days when used): For the participant with the highest use, the median duration was less than 60 minutes although the variation was large and indicates that there were days where use exceeded 90 minutes | Daily time: 60 minutes |
|  |  |  | Number of sessions (on days when used): 7 participants had days when they used the glove 3 times and all 13 had days where they used it twice. For 5 participants, there were days when they used the glove 4 times | Sessions per day: 3 |
|  | Swanson <sup>84</sup> | Technology | Median percentage of number of days used: 47% (range 23%-100%) | Total days: 21 |
|  |  |  | Mean total time: 5.4 (SD:4.1) hours | Total time: 9 hours |
|  |  |  | 9 out of 14 (64%) achieved theoretical target repetitions with 7 participants completing more than 3 times this amount<br>Mean total hand repetitions: 4051 (SD: 4986) (range: 130 -18617)<br>Mean total arm repetitions: 2744 (SD:3076) (range: 186 - 11262) | Theoretical target repetitions of at least 2700 |
|  | Wilson <sup>86</sup> | Unclear | Total sessions: 28 | Total sessions: 32 |
|  | Wolf <sup>87</sup> | Participant (time on activities/exercises). | Mean total time: 8052 (SD: 4042) minutes (range: 928 – 21195) | Total time: 7200 |
|  |  | Technology (time of use). | Mean total device use: 2172 (SD:1388) minutes (range: 12 – 5153) | Total device use: 4800 |
| Electrical stimulation | Alon <sup>77</sup> | Not measured | N/A |  |
|  | Fletcher-Smith <sup>82</sup> | Unclear | Mean number of sessions: 64.5(SD: 53) (range: 10- 166) | Number of sessions: 120 |
| Wearable Sensor | Da-Silva <sup>80</sup> | Technology | Median days worn: 25.0 (IQR: 21.8–28.0) | Number of days worn: 28 |
|  | Wei <sup>85</sup> | Not measured | N/A |  |
| Mobile application | Emmerson <sup>81</sup> | Participant | Mean percentage of exercise programme completed daily: 62% (SD:25)<br>Mean daily time spent completing HEP: 34 (SD:20) minutes | Lack of detail on daily content or time of planned exercise programme |

**Abbreviations**
IQR: Interquartile Range, SD: Standard Deviation
**Notes**
\*the time the device was in the participants homes was noted to have varied and this was considered in the calculation

### Risk of bias

RoB 2 assessment of all studies (using an activity limitation outcome) (Figure 2) identified ten studies (10/12, 83%) with some concerns ^78,79,81–88^. One study (1/12, 8%) was identified as low risk,^80^ and one study (1/12, 8%) as high risk.^77^ Across the included studies, domain five (selection of the reported result) raised the greatest number of concerns. The RoB 2 assessment specific to the completed meta-analyses is discussed below.

**Figure 2.**
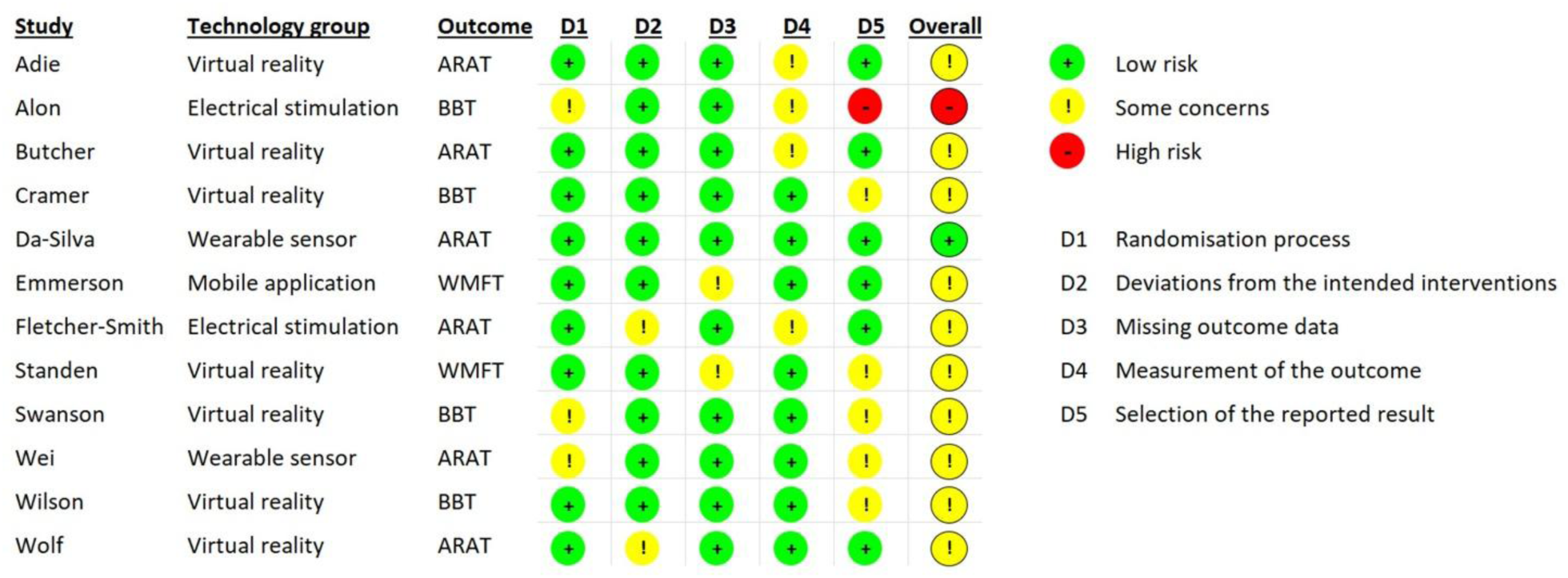
Risk of bias assessment using the RoB 2 tool

Regarding wider indicators of quality, one study (1/12, 8%) had no record of trial registration, provided no details of study approval processes and also had commercial funding sources aligned to the digital technology.^77^ Five studies (5/12, 42%) also declared conflicts not directed relating to study funding but which included associations with digital technology companies and product patents. ^78,79,84,85,87^

## Results of Syntheses

### Effectiveness of digital technology interventions: Motor impairment and activity limitation

A summary of the motor impairment and activity limitation findings for all studies is provided in Table 5. From the available studies and data, only two meta-analyses were completed, exploring the effectiveness of virtual reality on motor impairment and activity limitation outcomes. Subgroup analyses exploring the severity of the stroke population or how much of the intervention was delivered were not conducted as it was judged that there were insufficient studies for subgroup analysis to be meaningful.^64^ The data input into the meta-analysis is provided in Supplemental Table 6a-b and a summary of findings is provided in Table 6 which includes the GRADE judgement.

**Table 5:** Overview of activity limitation and impairment of function findings from studies.

| Technology group | Study | Impairment of function |  |  | Activity limitation |  |  |
| --- | --- | --- | --- | --- | --- | --- | --- |
|  |  | Outcome measure | Comparative analysis of groups | Included in meta-analysis | Outcome measure | Comparative analysis of groups | Included in meta-analysis |
| Virtual reality | Adie <sup>76</sup> | None measured |  |  | ARAT | Mean difference baseline to post intervention non-significant | Yes |
|  | Butcher <sup>78</sup> | FMA-UE | Mean difference baseline to post intervention non-significant | No | ARAT | Mean difference baseline to post intervention non-significant | No |
|  | Cramer <sup>79</sup> | FMA-UE | Mean difference baseline to 30 day follow up non-significant but indicating non-inferiority ((no post intervention data) | Yes | BBT | Mean difference baseline to 30 day follow up non-significant but indicating non-inferiority (no post intervention data) | Yes |
|  | Standen <sup>83</sup> | Grip strength | Difference baseline to post intervention non-significant | No | WMFT (time) | Difference baseline to post intervention non-significant | No |
|  |  | N/A |  |  | NHPT | Difference baseline to post intervention non-significant | No |
|  | Swanson <sup>84</sup> | FMA-UE | Difference baseline to 30 day follow-up **significant in favour of the experimental group** (significance p<0.05) (no post intervention experimental comparison) | Yes | BBT | Difference baseline to 30 day follow-up non-significant (no post intervention experimental comparison) | Yes |
|  | Wilson <sup>86</sup> | Non measured |  |  | BBT | Difference baseline to post intervention **significant in favour of the experimental group** (significance *p < Benjamini–Hochberg critical value) | Yes |
|  |  | N/A |  |  | NHPT | Difference baseline to post intervention non-significant | No |
|  | Wolf <sup>87</sup> | FMA-UE | Difference baseline to post intervention non-significant | Yes | ARAT | Difference baseline to post intervention non-significant | Yes |
|  |  | N/A |  |  | WMFT (time) | Difference baseline to post intervention **significant in favour of the comparator group** (significance p<0.05) | No |
| Electrical stimulation | Alon <sup>77</sup> | modified FMA-UE | Mean outcome data post intervention **significant in favour of the experimental group** (significance p<0.05) | No | BBT | Mean outcome data post intervention **significant in favour of the experimental group** (significance p<0.05) | No |
|  | Fletcher-Smith <sup>82</sup> | NIHSS arm | No comparative analysis | No | ARAT | No comparative analysis | No |
| Wearable Sensor | Da-Silva <sup>80</sup> | MI | No comparative analysis | No | ARAT | No comparative analysis | No |
|  | Wei <sup>85</sup> | FMA-UE | Difference baseline to 12 week follow-up non-significant (no post intervention comparison) | No | ARAT | Difference baseline to post intervention **significant in favour of the experimental group** (significance p=0.025) | No |
|  |  | N/A |  |  | BBT | Difference baseline to 12 week follow-up non-significant (no post intervention experimental comparison) | No |
| Mobile applications | Emmerson <sup>81</sup> | Grip strength | Mean difference baseline to post intervention non-significant | No | WMFT (time) | Mean difference baseline to post intervention non-significant | No |

**Abbreviations**
ARAT: Action Research Arm Test, BBT: Box and Block Test, FMA-UE: Fugl-Meyer Assessment for Upper Extremity, MI: Motricity Index, NHPT: Nine Hole Peg Test, NIHSS: National Institute of Health Stroke Scale, WMFT: Wolf Motor Function Test

**Table 6:** Summary of Findings.

| Virtual reality compared to conventional exercise therapy for upper limb rehabilitation at home in the first acute and subacute phases after stroke |  |  |  |  |  |
| --- | --- | --- | --- | --- | --- |
| Population: Adult participants with upper limb deficit as a result of a stroke within the previous six months |  |  |  |  |  |
| Setting: Experimental intervention, targeting the upper limb, delivered in the home setting |  |  |  |  |  |
| Experimental intervention: Virtual reality |  |  |  |  |  |
| Comparator Intervention: Conventional exercise therapy |  |  |  |  |  |
| Outcomes | Assumed effect<br>Comparator group | Corresponding effect<br>(95% CI)<br>Experimental group | Number of<br>participants<br>(studies) | Certainty of<br>the<br>evidence<br>(GRADE) | Notes |
| Post intervention impairment of motor function measured by FMA-UE reported as MD | Same amount of an alternative therapy approach | There was no apparent difference between groups, MD -0.23 (-2.58 to 2.12) | 241 (3 studies) | Low | The MD can be directly compared with the FMA-UE scale which has scores ranging from 0 to 66 with higher scores favourable. MCID scores of between 9 and 13 have been reported in acute and subacute stroke<sup>96–98</sup> |
| Post intervention limitation of activity measured by either the ARAT or BBT and reported as SMD | Same amount of an alternative therapy approach | There was no apparent difference between groups, SMD 0.07 (-0.25 to 0.38) | 467 (5 studies) | Low | The pooled SMD relates to an improvement of 0.99 on the ARAT (95% CI -3.76 to 5.77). ARAT MCID scores of 12 or 17 (depending on hand dominance) have been reported in subacute stroke<sup>99*</sup>. For both the ARAT and BBT higher scores are favourable |

**Abbreviations**
ARAT: Action Research Arm Test, BBT: Box and Block Test, CI; Confidence Interval, FMA-UE: Fugl-Meyer Assessment for Upper Extremity, MCID: Minimal Clinically Important Difference, MD: Mean difference, SMD: Standardised Mean Difference

### Virtual reality: Motor impairment

Of the five studies that reported an observed measure of motor impairment, ^78,79,83,84,87^ one (1/5, 20%) reported significant between group differences in favour of the experimental group using the virtual reality ^84^.

Two studies were ineligible for the meta-analysis due to lack of an active comparator,^78^ and data presented as median values.^83^ Three studies (241 participants) were included in the meta-analysis comparing a virtual reality intervention with a time matched conventional exercise programme.^79,84,87^ There was no evidence of differing effect on motor impairment (Fugl-Meyer Assessment for Upper Extremity) between the two groups (Figure 3; MD=-0.23, 95% CI:-2.58 to 2.12, p=0.85, I^2^=0%). The minimal clinically important difference for the Fugl-Meyer Assessment for Upper Extremity (reported between 9 and 13 in acute and subacute stroke)^96–98^ was not met by the pooled MD estimate nor the pooled 95% CI, indicating no clinically meaningful difference between the two groups. The RoB 2 assessment identified some concerns for all three studies (Supplemental Figure S1a). The certainty of the evidence was judged as low as it was downgraded for risk of bias and inconsistency (Supplemental Table S7). Whilst funnel plots were produced (Supplemental Figure S2a), interpretation was inconclusive due to small study numbers.^100^

**Figure 3:**
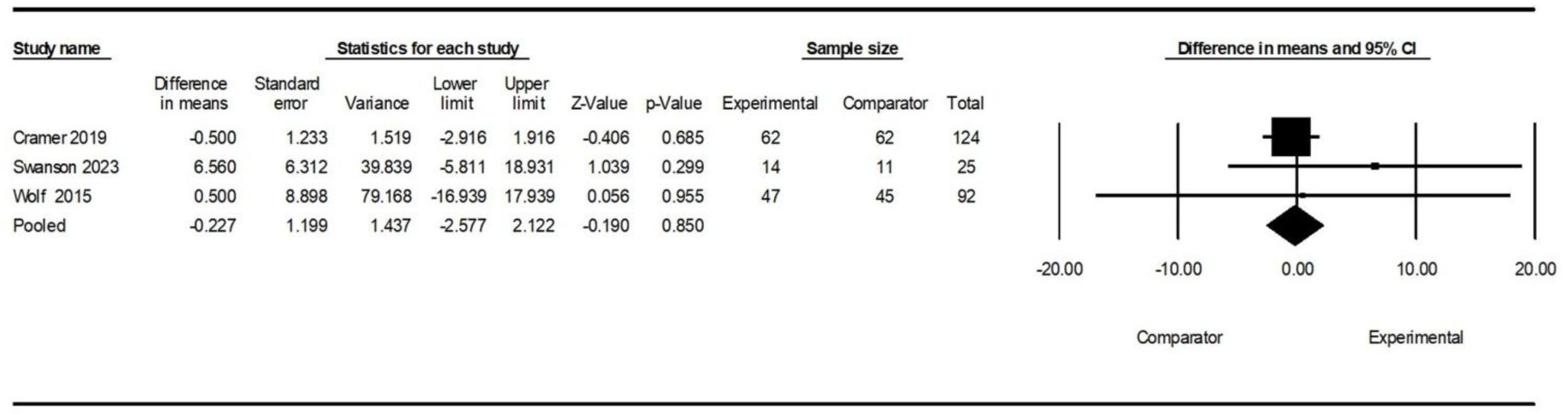
Forest plot for studies comparing the effects of virtual reality and conventional exercise programmes on motor impairment using the Fugel-Meyer Assessment for Upper Extremity

### Virtual reality: Activity limitation

Of the seven studies that reported an observed activity limitation measure, ^76,78,79,83,84,86,87^ two (2/7, 29%) reported significant between group changes, one study in favour of the virtual reality group (although a non-significant finding was reported for a second observed activity limitation measure),^86^ and one in favour of the comparator group which was conventional exercise.^87^

Two studies were ineligible for meta-analysis, ^78,83^ as described in the motor impairment section above and so five studies (467 participants) were included, comparing a virtual reality intervention with a time matched conventional exercise programme.^76,79,84,86,87^ There was no evidence of differing effect on activity limitation (Action Research Arm Test and Box and Block Test) between the virtual reality and conventional exercise groups (Figure 4; SMD= 0.07, 95% CI:-0.25 to 0.38, p=0.68, I^2^=54.87%). The pooled SMD was re-expressed in Action Research Arm Test points (using the representative SD of 15.3)^99^ and compared to the minimal clinically important difference (12 or 17 in subacute stroke, depending on hand dominance).^99^ The pooled SMD relates to an improvement of 0.99 on the Action Research Arm Test (95% CI −3.76 to 5.77), equivalent to 8% of one minimal important difference, indicating no clinically meaningful difference in change between the two groups.

**Figure 4:**
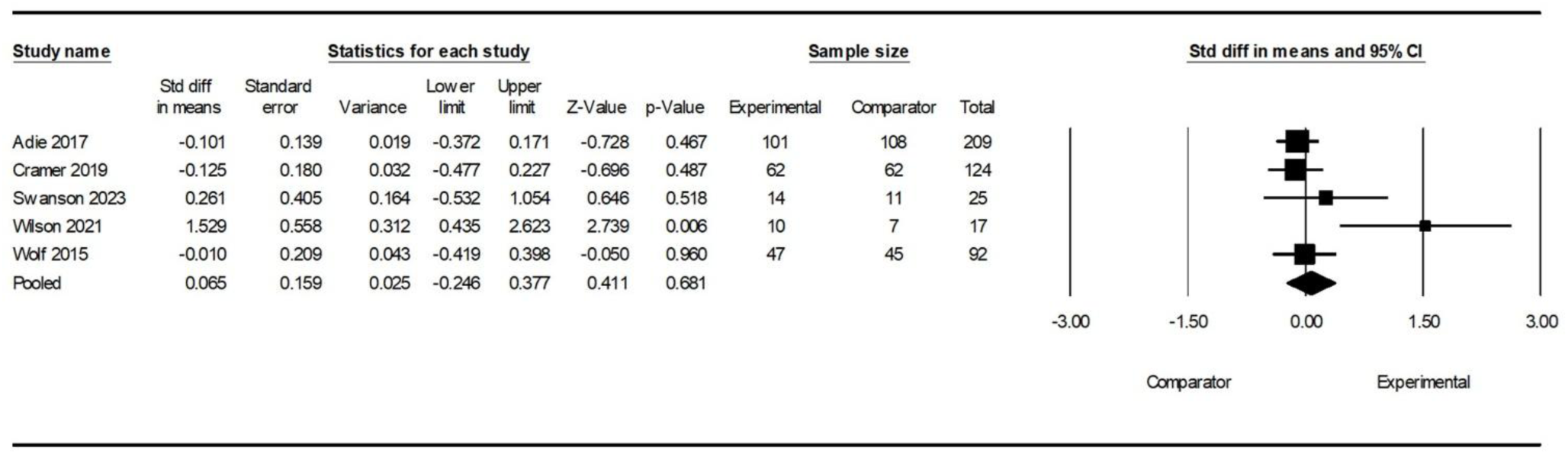
Forest plot for studies comparing the effects of virtual reality and conventional exercise programmes on combined measures of activity limitation (Action Research Arm Test and Box and Block Test)

The RoB 2 assessment identified some concerns for all five studies (Supplemental Figure S1b). The certainty of the evidence was judged as low with downgrading for risk of bias and inconsistency and funnel plots were inconclusive due to small study numbers (Supplemental Table S7 and Figure S2a).

### Other technology groups

Of the two studies that explored electrical stimulation,^77,82^ only one reported a comparative analysis with significant between group changes in favour of the experimental group for both the observed motor impairment outcome (modified Fugl-Meyer Assessment for Upper Extremity) and the observed activity limitation outcome (Box and Block Test).^77^ Meta-analysis was not completed as comparator groups were different in each study

Similarly, from the two studies investigating wearable sensors,^80,85^ only one reported a comparative analysis with significant between group changes in favour of the experimental group for the observed activity limitation outcome (ARAT) (although a non-significant finding was reported for a second observed activity limitation measure).^85^ Meta-analysis was not completed as one study presented data as median values.^80^

The final study, looking at a mobile application,^81^ did not find any significant between groups difference for either outcome domain.

### Effectiveness of digital technology interventions: Harm and adverse events

Only four studies (4/12, 33%) reported outcome measures for domains that may indicate harm including fatigue, spasticity and pain^78,80,82,84^ (Supplemental Table S8). Two of these (both investigating virtual reality) compared indicators of possible harm between groups,^78,84^ with no significant differences identified.

Adverse event data was collected in ten (10/12, 83%) of the studies.^76–81,84–87^ (Supplemental Table S8). No studies identified any serious adverse events associated with an intervention. Non-serious adverse events were only reported for three of the virtual reality studies. Two studies identified non-serious adverse events that were at least possibly associated with the intervention,^78,79^ and one study identified non-serious adverse events but without commenting on possible associations.^87^

### Economic evaluation

Two studies (2/12, 17%) reported the results of an economic evaluation; one exploring virtual reality,^76^ and one electrical stimulation^82^ (Supplemental Table S9). For the virtual reality intervention,^76^ there was no between group difference in mean quality adjusted life years at six months, however the virtual reality intervention group had higher health and social care costs, (although this higher cost was not solely related to the digital technology used). For the electrical stimulation intervention,^82^ only a small difference, in favour of the electrical stimulation group, in mean quality adjusted life years at twelve months was reported but the electrical stimulation group also had lower costs from a health and societal perspective. This needs to be considered in the context of a small sample size and minor discrepancy in participant ages which may could have influenced the between group different in cost of lost productivity.

## Discussion

This systematic review sought to determine the effectiveness of a range of digital technology interventions used to address motor impairment and activity limitation of the upper limb, at home, in the acute and subacute phases (first six months) following stroke. It is the first to focus on this specific area of stroke rehabilitation. Four broad groups of digital technologies were identified from the included studies; virtual reality, electrical stimulation, wearable sensors and mobile applications, however, only studies comparing a virtual reality intervention with a time-matched conventional exercise programme were able to be combined for meta-analysis. This was due to low numbers of studies, differences in comparator group interventions and the way summary data was presented.

The meta-analysis findings indicate that there was no evidence of a difference in motor impairment and activity limitation outcomes between a virtual reality intervention and a time-matched conventional exercise programme, when delivered at home in the acute and subacute phases (first six months) following stroke. This evidence was deemed as low certainty due to risk of bias and inconsistency. In the context of home-based rehabilitation our findings are in line with a meta-analysis evaluating the effect of digital games within the home environment,^39^ despite their inclusion of a broader group of comparator interventions. In comparison, another meta-analysis reported benefits of digital technology rehabilitation (digital games, robotics and virtual reality) at home.^38^ However this analysis pooled data from a broader range of outcome domains than our review, and multiple outcomes from individual studies, and also lacked clarity on the comparator interventions included. In the context of the acute and subacute phases after stroke, review findings follow a similar pattern. A recent Cochrane review and meta-analysis comparing virtual reality with an alternative therapy,^22^ reported a small effect in favour of virtual reality, however the subgroup analysis considering time post stroke onset suggested no evidence of effect in the first six months after stroke. A meta-analysis focusing on virtual reality in the first month after stroke,^21^ also reported no evidence of effect when compared to a conventional therapy or usual care. In contrast, in the first six months after stroke, a meta-analysis of digital games^27^ indicated benefit and a meta-analysis of virtual reality indicated improvements for motor impairment outcomes but not activity limitation outcomes.^19^ Both these reviews used comparator groups that included a mix of dose matched and non-dose matched conventional therapies or no therapy, and the data related to the time since stroke onset was explored in subgroup analyses. In addition to indicating no evidence of difference between virtual reality and conventional exercise, this review did not identify concerns of serious harm from the virtual reality. A limited number of attributable non-serious adverse events were reported in line with other review findings.^21,22^

Formal exploration of an association between the severity of upper limb motor impairment or activity limitation, or the dose of the intervention, on the effect of virtual reality was not completed due to the limited number of studies included in the meta-analysis. The influence of these moderators is therefore unknown but the review data raises some noteworthy points. Of significance is that all studies required participants to have some upper limb motor function and activity, and hence it can be concluded that these findings are not applicable to those with no motor function or activity in their upper limb. The calculated planned average daily intervention time for the seven virtual reality studies varied from 17 to 129 minutes per day with a median of 60 minutes. A recent study has indicated that up to 135 minutes a day could be safely tolerated in the acute phase after stroke. ^101^ Whilst all included studies planned lower daily intervention times than this, the intervention times reported do not reflect any usual care delivered alongside the intervention, and the 135 minutes is related to safe rather than efficacious intervention time. Therefore, it cannot be determined if an optimal intervention time was achieved in the included studies. Notably the adherence data indicates that the planned intervention time was not met in the majority of these virtual reality interventions.

There was insufficient data to complete meta-analyses of the effectiveness of the other groups of digital technologies (electrical stimulation, wearable sensors and mobile applications). Published meta-analyses on electrical stimulation indicate its benefit in the acute and subacute phases after stroke,^28,30^ but do not specifically consider its use at home. The review literature on wearable sensors and mobile applications is broad (across phases of stroke recovery and settings) and lacks statistical synthesis. A scoping review,^102^ identified that upper limb wearable sensors have potential use as an adjunct to increase adherence to interventions through sensory reminders and feedback. From three systematic reviews investigating mobile applications for multiple aspects of stroke rehabilitation,^103–105^ a lack of RCT evidence for upper limb mobile applications was identified, however individual studies focusing on upper limb rehabilitation did indicate potential for use.

The generalisability of these findings should be considered within the context of the population in the studies. Studies were found which directly met the PICO criteria however participants appeared somewhat younger than the general population of stroke survivors with eight (of the eleven studies reporting age) identifying a mean or median of less than 70 years. Although mean age of stroke onset is falling, it still remains just above 70.^106^ There was also little data provided on key indicators of education and socioeconomic status, despite their clear association with health status, and disparities in access to, use and outcomes from digital technologies.^107^

Reporting of the interventions against the TIDieR-Rehab checklist highlighted a lack of reporting for the essential amount and challenge aspects of dose, in line with a recent review of reporting quality in studies of robotic interventions for upper limb rehabilitation post-stroke.^108^ Reporting of these elements, that reflect the multidimensional nature of dose and not just time elements, are crucial to developing the understanding of the relationship between dose and outcomes and enable more effective rehabilitation.^54^ Digital technologies themselves offer opportunities to facilitate the collection of a diverse range of dose related data and reduce the burden on participant and therapist data collection which is prone to subjectivity,^54,102^ although this review suggests that digital technologies have not yet been used to their full potential to do this.

Moving forwards, whilst a firm conclusion of comparability of virtual reality and conventional exercise programmes cannot be drawn from the meta-analysis data, the authors cautiously suggest that the use of virtual reality technologies may provide an alternative intervention to conventional exercise therapy for home based rehabilitation in the acute and subacute phases following stroke. Further research is therefore needed to understand who may benefit from a virtual reality intervention, through high quality studies focusing on a specific subgroup or identifying subgroups within a larger study, and individual participant data meta-analysis. Research to improve understanding of the impact of the dose (considering all aspects of dose and not just time) of virtual reality interventions is also crucial to ensure sufficient dose is provided to maximise clinical outcomes. The absence of economic evaluation in all but one of the virtual reality studies in this review should be remedied in future work to inform clinical decision making in the context of limited financial resources. Further robust studies of the clinical and cost effectiveness of electrical stimulation, wearable sensors and mobile applications within the home environment in the acute and subacute phases post-stroke is also needed.

These findings need to be interpreted within the limitations of this review. Despite a comprehensive search strategy, and contact with trial registry and protocol authors, the number of studies identified is low especially for each different type of digital technology. This may be a reflection of the relative immaturity of evidence in this area but could have been influenced by the exclusion of unpublished work and studies not published in English; decisions which may increase the risk of reporting bias from both publication bias and language bias.^109^ The low number of studies limited the meta-analyses completed, and interpretation of publication bias. These alongside concerns raised by the risk of bias assessment reduce the certainty of the conclusions that can be drawn from the data. Whilst the meta-analyses and overall conclusions are limited, a strength of this review is the use of the TIDieR-Rehab checklist,^53^ to provide comprehensive and structured detail on the reported interventions. Our approach to the grouping of the digital technology interventions bought together interventions that served a similar function. The largest group included interventions which involved an element of real time, motion-based interaction with the digital devices. The review authors considered this group could be best described under the established umbrella term of virtual reality,^110^ but acknowledges that this is a very broad interpretation and therefore includes studies that other reviews focussing specifically on virtual reality may have actively excluded.^19,22^ Some studies in this group also included the use of additional conventional and digital technology components alongside the virtual reality, the effect of which remains unexplored.

In conclusion evidence of the effectiveness of using digital technology interventions to address motor impairment and activity limitation of the upper limb, at home in the acute and subacute phases (first six months) following stroke is limited. With regards to interventions which incorporate an element of virtual reality, there was no evidence of difference between the virtual reality and time-matched conventional exercise. No summary conclusions could be drawn regarding the other digital technologies; electrical stimulation, wearable sensors and mobile applications. The further research outlined above is needed to understand, if and how virtual reality interventions could provide an alternative therapy option, and the effectiveness of the other digital technologies in order to support clinical decision making.

### Clinical Messages

In the context of upper limb rehabilitation to address motor impairment and activity limitation, that is completed at home in the acute and subacute phases (first six months) following stroke:

- For interventions which incorporate an element of virtual reality, there was no evidence of difference between the virtual reality and time-matched conventional exercise.
- Virtual reality technologies may provide an alternative intervention to conventional exercise therapy but further research is needed to confirm this, including an understanding of who may benefit, what the optimal dose may be and what the economic implications are for this type of intervention.
- Evidence surrounding the use of electrical stimulation, wearable sensors and mobile applications is too limited to draw conclusions. Further effectiveness research is indicated in this area.

## Data Availability

All data can be found in the original articles. Data summaries pertinent to the review and further detail on GRADE judgements are provided in the supplemental material.

## References

1. GBD 2019 Stroke Collaborators. Global, regional, and national burden of stroke and its risk factors, 1990–2019: a systematic analysis for the Global Burden of Disease Study 2019. Lancet Neurol 2021; 20: 795–820.

2. Lawrence ES, Coshall C, Dundas R, et al. Estimates of the prevalence of acute stroke impairments and disability in a multiethnic population. Stroke 2001; 32: 1279–1284.

3. Broeks JG, Lankhorst GJ, Rumping K, et al. The long-term outcome of arm function after stroke: results of a follow-up study. Disabil Rehabil 1999; 21: 357–364.

4. Kwakkel G, Kollen BJ, Van der Grond J, et al. Probability of regaining dexterity in the flaccid upper limb. Stroke 2003; 34: 2181–2186.

5. Lai SM, Studenski S, Duncan PW, et al. Persisting consequences of stroke measured by the stroke impact scale. Stroke 2002; 33: 1840–1844.

6. Doyle SD, Bennett S, Dudgeon B. Upper limb post-stroke sensory impairments: the survivor’s experience. Disabil Rehabil 2014; 36: 993–1000.

7. Purton J, Sim J and Hunter SM. The experience of upper-limb dysfunction after stroke: a phenomenological study. Disabil Rehabil 2021; 43: 3377–3386.

8. Harris JE and Eng JJ. Paretic upper-limb strength best explains arm activity in people with stroke. Phys Ther 2007; 87: 88–97.

9. Sveen U, Bautz-Holter E, Sødring KM, et al. Association between impairments, self-care ability and social activities 1 year after stroke. Disabil Rehabil 1999; 21: 372–377.

10. Veerbeek JM, Kwakkel G, Van Wegen EE, et al. Early prediction of outcome of activities of daily living after stroke: a systematic review. Stroke 2011; 42: 1482–1488.

11. Intercollegiate Stroke Working Party. National clinical guideline for stroke for the UK and Ireland. London: Intercollegiate Stroke Working Party, 2023. Available from www.strokeguideline.org.

12. Bernhardt J, Hayward KS, Kwakkel G, et al. Agreed definitions and a shared vision for new standards in stroke recovery research: The Stroke Recovery and Rehabilitation Roundtable taskforce. Int J Stroke 2017; 12: 444–450.

13. Borschmann KN and Hayward KS. Recovery of upper limb function is greatest early after stroke but does continue to improve during the chronic phase: a two-year, observational study. Physiotherapy 2020; 107: 216–223.

14. Cortes JC, Goldsmith J, Harran MD, et al. A short and distinct time window for recovery of arm motor control early after stroke revealed with a global measure of trajectory kinematics. Neurorehabil Neural Repair 2017; 31: 552–560.

15. Krakauer JW, Carmichael ST, Corbett D, et al. Getting neurorehabilitation right: what can be learned from animal models? Neurorehabil Neural Repair 2012; 26: 923–931.

16. National Institute for Health and Care Excellence. Stroke rehabilitation in adults: NICE guideline [NG236]. London: NICE, 2023. Available from www.nice.org.uk/guidance/ng236.

17. Lin DJ, Cramer SC, Boyne P, et al. High-dose, high-intensity stroke rehabilitation: why aren’t we giving it? Stroke 2025; 56: 1351–1364.

18. Schneider EJ, Lannin NA, Ada L, et al. Increasing the amount of usual rehabilitation improves activity after stroke: a systematic review. J Physiother 2016; 62: 182–187.

19. Chen J, Or CK and Chen T. Effectiveness of using virtual reality–supported exercise therapy for upper extremity motor rehabilitation in patients with stroke: systematic review and meta-analysis of randomized controlled trials. J Med Internet Res 2022; 24: e24111.

20. Domínguez-Téllez P, Moral-Muñoz JA, Salazar A, et al. Game-based virtual reality interventions to improve upper limb motor function and quality of life after stroke: systematic review and meta-analysis. Games Health J 2020; 9: 1–10.

21. Hao J, Yao Z, Harp K, et al. Effects of virtual reality in the early-stage stroke rehabilitation: a systematic review and meta-analysis of randomized controlled trials. Physiother Theory Pract 2023; 39: 2569–2588.

22. Laver KE, Lange B, George S, et al. Virtual reality for stroke rehabilitation. Cochrane Database Syst Rev 2025; 6: CD008349.

23. Leong SC, Tang YM, Toh FM, et al. Examining the effectiveness of virtual, augmented, and mixed reality (VAMR) therapy for upper limb recovery and activities of daily living in stroke patients: a systematic review and meta-analysis. J NeuroEngineering Rehabil 2022; 19: 93.

24. Mehrholz J, Pohl M, Platz T, et al. Electromechanical and robot-assisted arm training for improving activities of daily living, arm function, and arm muscle strength after stroke. Cochrane Database Syst Rev 2018; 9: CD006876.

25. Veerbeek JM, Langbroek-Amersfoort AC, Van Wegen EEH, et al. Effects of robot-assisted therapy for the upper limb after stroke: a systematic review and meta-analysis. Neurorehabil Neural Repair 2017; 31: 107–121.

26. Yang X, Shi X, Xue X, et al. Efficacy of robot-assisted training on rehabilitation of upper limb function in patients with stroke: a systematic review and meta-analysis. Arch Phys Med Rehabil 2023; 104: 1498–1513.

27. Doumas I, Everard G, Dehem S, et al. Serious games for upper limb rehabilitation after stroke: a meta-analysis. J Neuroeng Rehabil 2021; 18: 100.

28. Eraifej J, Clark W, France B, et al. Effectiveness of upper limb functional electrical stimulation after stroke for the improvement of activities of daily living and motor function: a systematic review and meta-analysis. Syst Rev 2017; 6: 40.

29. Kristensen MGH, Busk H and Wienecke T. Neuromuscular electrical stimulation improves activities of daily living post stroke: a systematic review and meta-analysis. Arch Rehabil Res Clin Transl 2022; 4: 100167.

30. Yang JD, Liao CD, Huang SW, et al. Effectiveness of electrical stimulation therapy in improving arm function after stroke: a systematic review and a meta-analysis of randomised controlled trials. Clin Rehabil 2019; 33: 1286–1297.

31. Laver KE, Adey-Wakeling Z, Crotty M, et al. Telerehabilitation services for stroke. Cochrane Database Syst Rev 2020; 1: CD010255.

32. Darzi A. Independent investigation of the National Health Service in England. London: GOV.UK, 2024. Available from www.gov.uk/government/publications/independent-investigation-of-the-nhs-in-england.

33. National Institute for Health and Care Excellence. Stroke in adults: quality standard [QS2]. London: NICE, 2010.Available from www.nice.org.uk/guidance/QS2.

34. UK Government. Fit for the future: the 10 year health plan for England. London: GOV.UK, 2025. Available from www.gov.uk/government/publications/10-year-health-plan-for-england-fit-for-the-future.

35. Sentinel Stroke National Audit Programme. SSNAP clinical audit April 2013 to March 2018 annual public report. London: SSNAP, 2019. Available from www.hqip.org.uk/wp-content/uploads/2019/06/Ref-142-SSNAP-Annual-Report-FINAL.pdf.

36. Sentinel Stroke National Audit programme. SSNAP spotlight report: stroke rehabilitation. London: SSNAP, 2024. Available from www.strokeaudit.org/SupportFiles/Documents/Annual-report/2024/SSNAP-Annual-Report-2024-Therapy-Spotlight-Report.aspx

37. Gebreheat G, Goman A and Porter-Armstrong A. The use of home-based digital technology to support post-stroke upper limb rehabilitation: a scoping review. Clin Rehabil 2024; 38: 60–71.

38. Bok SK, Song Y, Lim A, et al. High-tech home-based rehabilitation after stroke: a systematic review and meta-analysis. J Clin Med 2023; 12: 2668.

39. Gelineau A, Perrochon A, Robin L, et al. Measured and perceived effects of upper limb home-based exergaming Interventions on activity after stroke: a systematic review and meta-analysis. Int J Environ Res Public Health 2022; 19: 9112.

40. Hestetun-Mandrup AM, Toh ZA, Oh HX, et al. Effectiveness of digital home rehabilitation and supervision for stroke survivors: a systematic review and meta-analysis. Digit Health 2024; 10: 20552076241256861.

41. Huang J, Wei Y, Zhou P, et al. Effect of home-based virtual reality training on upper extremity recovery in patients with stroke: systematic review. J Med Internet Res 2025; 27: e69003.

42. Higgins JPT, Thomas J, Chandler J, et al. (eds). Cochrane handbook for systematic reviews of interventions. Version 6.5 (updated August 2024). Cochrane, 2024. Available from www.cochrane.org/authors/handbooks-and-manuals/handbook/current.

43. Page MJ, McKenzie JE, Bossuyt PM, et al. The PRISMA 2020 statement: an updated guideline for reporting systematic reviews. BMJ 2021; 372: n71.

44. Thomas J, Kneale D, McKenzie JE, et al. Chapter 2: Determining the scope of the review and the questions it will address [last updated August 2023]. In: Higgins JPT, Thomas J, Chandler J, et al. (eds). Cochrane handbook for systematic reviews of interventions. Version 6.5 (updated August 2024). Cochrane, 2024. Available from www.cochrane.org/authors/handbooks-and-manuals/handbook/current.

45. World Health Organisation. International classification of functioning, disability and health (ICF). Geneva: World Health Organisation, 2001 Available from www.who.int/standards/classifications/international-classification-of-functioning-disability-and-health.

46. Hillyer M. Here’s how technology has changed the world since 2000. World Economic Forum, (2020, accessed 20 February 2025).

47. The EndNote Team. EndNote version 21. Philadelphia PA: Clarivate, 2013.

48. Falconer J. Removing duplicates from an EndNote library. Library, Archive & Open Research Services Blog, https://blogs.lshtm.ac.uk/library/2018/12/07/removing-duplicates-from-an-endnote-library/ (2018, accessed 24 February 2025).

49. Ouzzani M, Hammady H, Fedorowicz Z, et al. Rayyan—a web and mobile app for systematic reviews. Syst Rev 2016; 5: 210.

50. Microsoft Corporation. Microsoft Excel version 2510. Redmond WA: Microsoft Corporation, 2021.

51. O’Neill J, Tabish H, Welch V, et al. Applying an equity lens to interventions: using PROGRESS ensures consideration of socially stratifying factors to illuminate inequities in health. J Clin Epidemiol 2014; 67: 56–64.

52. Hoffmann TC, Glasziou PP, Boutron I, et al. Better reporting of interventions: template for intervention description and replication (TIDieR) checklist and guide. BMJ 2014; 348: g1687.

53. Signal N, Gomes E, Olsen S, et al. Enhancing the reporting quality of rehabilitation interventions through an extension of the Template for Intervention Description and Replication (TIDieR): the TIDieR-Rehab checklist and supplementary manual. BMJ Open 2024; 14: e084320.

54. Hayward KS, Churilov L, Dalton EJ, et al. Advancing stroke recovery through improved articulation of nonpharmacological intervention dose. Stroke 2021; 52: 761–769.

55. Higgins JPT, Savović J, Page MJ, et al. Chapter 8: Assessing risk of bias in a randomized trial [last updated October 2019]. In: Higgins JPT, Thomas J, Chandler J, et al. (eds). Cochrane handbook for systematic reviews of interventions. Version 6.5 (updated August 2024). Cochrane, 2024. Available from www.cochrane.org/authors/handbooks-and-manuals/handbook/current.

56. Sterne JAC, Savović J, Page MJ, et al. RoB 2: a revised tool for assessing risk of bias in randomised trials. BMJ 2019; 366: l4898.

57. Risk of bias.info authors. Current version of RoB 2. Risk of bias.info, https://www.riskofbias.info/welcome/rob-2-0-tool/current-version-of-rob-2 (2025, accessed 2 September 2025).

58. IBM Corporation. IBM SPSS Statistics version 31.0.0.0. Armonk NY: IBM Corporation, 2025.

59. Kwakkel G, Lannin NA, Borschmann K, et al. Standardized measurement of sensorimotor recovery in stroke trials: consensus-based core recommendations from the Stroke Recovery and Rehabilitation Roundtable. Int J Stroke 2017; 12: 451–461.

60. Pohl J, Held JPO, Verheyden G, et al. Consensus-based core set of outcome measures for clinical motor rehabilitation after stroke—a delphi study. Front Neurol 2020; 11: 875.

61. Borenstein M, Hedges L, Higgins J, et al. Comprehensive Meta-Analysis version 4. Englewood NJ: Biostat, 2022.

62. Borenstein M, Hedges LV, Higgins JPT, et al. A basic introduction to fixed-effect and random-effects models for meta-analysis. Res Synth Methods 2010; 1: 97–111.

63. Borenstein M, Hedges LV, Higgins JPT, et al. Introduction to meta-analysis. 2nd ed. Hoboken: Wiley, 2021.

64. Deeks JJ, Higgins JPT, Altman DG, et al. Chapter 10: Analysing data and undertaking meta-analyses [last updated November 2024]. In: Higgins JPT, Thomas J, Chandler J, et al. (eds). Cochrane handbook for systematic reviews of interventions. Version 6.5 (updated August 2024). Cochrane, 2024. Available from www.cochrane.org/authors/handbooks-and-manuals/handbook/current.

65. DerSimonian R and Laird N. Meta-analysis in clinical trials. Control Clin Trials 1986; 7: 177–188.

66. DerSimonian R and Laird N. Meta-analysis in clinical trials revisited. Contemp Clin Trials 2015; 45: 139–145.

67. Hedges LV and Vevea JL. Fixed-and random-effects models in meta-analysis. Psychol Methods 1998; 3: 486–504.

68. Borenstein M. In a meta-analysis, the I-squared statistic does not tell us how much the effect size varies. J Clin Epidemiol 2022; 152: 281–284.

69. Higgins JPT and Thompson SG. Quantifying heterogeneity in a meta-analysis. Stat Med 2002; 21: 1539–1558.

70. Higgins JPT. Commentary: heterogeneity in meta-analysis should be expected and appropriately quantified. Int J Epidemiol 2008; 37: 1158–1160.

71. Higgins JPT, Thompson SG, Deeks JJ, et al. Measuring inconsistency in meta-analyses. BMJ 2003; 327: 557–560.

72. Higgins JPT, Li T and Deeks JJ. Chapter 6: Choosing effect measures and computing estimates of effect [last updated August 2023]. In: Higgins JPT, Thomas J, Chandler J, et al. (eds). Cochrane handbook for systematic reviews of interventions. Version 6.5 (updated August 2024). Cochrane, 2024. Available from www.cochrane.org/authors/handbooks-and-manuals/handbook/current.

73. Schünemann HJ, Vist GE, Higgins JPT, et al. Chapter 15: Interpreting results and drawing conclusions [last updated August 2023]. In: Higgins JPT, Thomas J, Chandler J, et al. (eds). Cochrane handbook for systematic reviews of interventions. Version 6.5 (updated August 2024). Cochrane, 2024. Available from www.cochrane.org/authors/handbooks-and-manuals/handbook/current.

74. Schünemann HJ, Higgins JPT, Vist, GE, et al. Chapter 14: Completing ‘summary of findings’ tables and grading the certainty of the evidence [last updated August 2023]. In: Higgins JPT, Thomas J, Chandler J, et al. (eds). Cochrane handbook for systematic reviews of interventions. Version 6.5 (updated August 2024). Cochrane, 2024. Available from www.cochrane.org/authors/handbooks-and-manuals/handbook/current.

75. Schünemann H, Brożek J, Guyatt G, et al. (eds). GRADE handbook for grading quality of evidence and strength of recommendations. Updated October 2013. The GRADE Working Group, 2013. Available from www.guidelinedevelopment.org/handbook.

76. Adie K, Schofield C, Berrow M, et al. Does the use of Nintendo Wii Sports^TM^ improve arm function? Trial of Wii^TM^ in Stroke: a randomized controlled trial and economics analysis. Clin Rehabil 2017; 31: 173–185.

77. Alon G, Levitt AF and McCarthy PA. Functional electrical stimulation enhancement of upper extremity functional recovery during stroke rehabilitation: a pilot study. Neurorehabil Neural Repair 2007; 21: 207–215.

78. Butcher T, Warland A, Stewart V, et al. Rehabilitation using virtual gaming for Hospital and hOMe-Based training for the Upper limb in acute and subacute Stroke (RHOMBUS II): results of a feasibility randomised controlled trial. BMJ Open 2025; 15: e089672.

79. Cramer SC, Dodakian L, Le V, et al. Efficacy of home-based telerehabilitation vs in-clinic therapy for adults after stroke: a randomized clinical trial. JAMA Neurol 2019; 76: 1079–1087.

80. Da-Silva RH, Moore SA, Rodgers H, et al. Wristband Accelerometers to motiVate arm Exercises after Stroke (WAVES): a pilot randomized controlled trial. Clin Rehabil 2019; 33: 1391–1403.

81. Emmerson KB, Harding KE and Taylor NF. Home exercise programmes supported by video and automated reminders compared with standard paper-based home exercise programmes in patients with stroke: a randomized controlled trial. Clin Rehabil 2017; 31: 1068–1077.

82. Fletcher-Smith JC, Walker D-M, Allatt K, et al. The ESCAPS study: a feasibility randomized controlled trial of early electrical stimulation to the wrist extensors and flexors to prevent post-stroke complications of pain and contractures in the paretic arm. Clin Rehabil 2019; 33: 1919–1930.

83. Standen PJ, Threapleton K, Richardson A, et al. A low cost virtual reality system for home based rehabilitation of the arm following stroke: a randomised controlled feasibility trial. Clin Rehabil 2017; 31: 340–350.

84. Swanson VA, Johnson C, Zondervan DK, et al. Optimized home rehabilitation technology reduces upper extremity impairment compared to a conventional home exercise program: a randomized, controlled, single-blind trial in subacute stroke. Neurorehabil Neural Repair 2023; 37: 53–65.

85. Wei WXJ, Fong KNK, Chung RCK, et al. “Remind-to-Move” for promoting upper extremity recovery using wearable devices in subacute stroke: a multi-center randomized controlled study. IEEE Trans Neural Syst Rehabil Eng 2019; 27: 51–59.

86. Wilson PH, Rogers JM, Vogel K, et al. Home-based (virtual) rehabilitation improves motor and cognitive function for stroke patients: a randomized controlled trial of the Elements (EDNA-22) system. J NeuroEngineering Rehabil 2021; 18: 165.

87. Wolf SL, Sahu K, Bay RC, et al. The HAAPI (Home Arm Assistance Progression Initiative) trial: a novel robotics delivery approach in stroke rehabilitation. Neurorehabil Neural Repair 2015; 29: 958–968.

88. Adie K, Schofield C, Berrow M, et al. Does the use of Nintendo Wii Sports^TM^ improve arm function and is it acceptable to patients after stroke? Publication of the Protocol of the trial of Wii^TM^ in Stroke – TWIST. Int J Gen Med 2014; 7: 475–481.

89. Fletcher-Smith JC, Walker D-M, Sprigg N, et al. ESCAPS study protocol: a feasibility randomised controlled trial of ‘Early electrical stimulation to the wrist extensors and wrist flexors to prevent the post-stroke complications of pain and contractures in the paretic arm’. BMJ Open 2016; 6: e010079.

90. Kilbride C, Warland A, Stewart V, et al. Rehabilitation using virtual gaming for Hospital and hOMe-Based training for the Upper limb post Stroke (RHOMBUS II): protocol of a feasibility randomised controlled trial. BMJ Open 2022; 12: e058905.

91. Linder SM, Rosenfeldt AB, Reiss A, et al. The home stroke rehabilitation and monitoring system trial: A randomized controlled trial. Int J Stroke 2013; 8: 46–53.

92. Linder SM, Rosenfeldt AB, Bay RC, et al. Improving quality of life and depression after stroke through telerehabilitation. Am J Occup Ther 2015; 69: 6902290020p1-6902290020p10.

93. Moore SA, Da-Silva R, Balaam M, et al. Wristband Accelerometers to motiVate arm Exercise after Stroke (WAVES): study protocol for a pilot randomized controlled trial. Trials 2016; 17: 508.

94. Standen PJ, Brown DJ, Battersby S, et al. A study to evaluate a low cost virtual reality system for home based rehabilitation of the upper limb following stroke. Int J Disabil Hum Dev 2011; 10: 337–341.

95. Standen PJ, Threapleton K, Connell L, et al. Patients’ use of a home-based virtual reality system to provide rehabilitation of the upper limb following stroke. Phys Ther 2015; 95: 350–359.

96. Hiragami S, Inoue Y and Harada K. Minimal clinically important difference for the Fugl-Meyer Assessment of the upper extremity in convalescent stroke patients with moderate to severe hemiparesis. J Phys Ther Sci 2019; 31: 917–921.

97. Arya KN, Verma R and Garg RK. Estimating the minimal clinically important difference of an upper extremity recovery measure in subacute stroke patients. Top Stroke Rehabil 2011; 18: 599–610.

98. Huynh BP, DiCarlo JA, Vora I, et al. Sensitivity to change and responsiveness of the upper extremity Fugl-Meyer Assessment in individuals with moderate to severe acute stroke. Neurorehabil Neural Repair 2023; 37: 545–553.

99. Lang CE, Edwards DF, Birkenmeier RL, et al. Estimating minimal clinically important differences of upper-extremity measures early after stroke. Arch Phys Med Rehabil 2008; 89: 1693–1700.

100. Sterne JAC, Sutton AJ, Ioannidis JPA, et al. Recommendations for examining and interpreting funnel plot asymmetry in meta-analyses of randomised controlled trials. BMJ 2011; 343: d4002.

101. Triccas LT, Hallet P, Cardeynaels S, et al. Higher doses of intensive upper limb rehabilitation for moderate to severe impairment in acute, subacute stroke: phase I dose escalation study. Neurorehabil Neural Repair 2025; 39: 752–764.

102. Kim GJ, Parnandi A, Eva S, et al. The use of wearable sensors to assess and treat the upper extremity after stroke: a scoping review. Disabil Rehabil 2022; 44: 6119–6138.

103. Rintala A, Kossi O, Bonnechère B, et al. Mobile health applications for improving physical function, physical activity, and quality of life in stroke survivors: a systematic review. Disabil Rehabil 2023; 45: 4001–4015.

104. Szeto SG, Wan H, Alavinia M, et al. Effect of mobile application types on stroke rehabilitation: a systematic review. J Neuroeng Rehabil 2023; 20: 12.

105. Zhou X, Du M and Zhou L. Use of mobile applications in post-stroke rehabilitation: a systematic review. Top Stroke Rehabil 2018; 25: 489–499.

106. Public Health England. Briefing document: first incidence of stroke estimates for England 2007 to 2016. London: PHE publications, 2018. Available from https://assets.publishing.service.gov.uk/media/5a82ab52e5274a2e8ab58bb5/Stroke_incidence_briefing_document_2018.pdf.

107. Lythreatis S, Singh SK, El-Kassar A-N. The digital divide: a review and future research agenda. Technol Forecast Soc Change 2022; 175: 121359.

108. Gomes E, Alder G, Boardsworth K, et al. Innovative but difficult to replicate: a systematic review of the reporting quality of robotic and conventional upper-limb interventions in stroke rehabilitation randomized controlled trials using the TIDieR-Rehab checklist. Appl Sci 2025; 15: 8487.

109. Sedgwick P. Meta-analyses: how to read a funnel plot. BMJ 2013; 346: f1342.

110. Abbas JR, O’Connor A, Ganapathy E, et al. What is virtual reality? A healthcare-focused systematic review of definitions. Health Policy Technol 2023; 12: 100741.

