## Supplemental material for "The Effectiveness and Use of Digital Technology Interventions for Upper Limb Rehabilitation at Home in Acute and Subacute Stroke: A Systematic Review and Meta-Analysis"

### Table S1. Eligibility Criteria

| Criteria Category | | Inclusion | Exclusion | Exclusion Code$ |
|---|---|---|---|---|
| **Evidence Source** | **Date** | 2000-2025 | Pre 2000 | 1 |
| | **Language** | English | Non-English | 2 |
| | **Source** | • Full text published peer reviewed work<br>• Theses | • Standalone conference abstracts*<br>• Standalone protocols and trial registries* | 3 Conference<br>3 Protocol/trial - ineligible (note reason)<br>3 Protocol/trial – ongoing study<br>3 Protocol/trial -no full publication available or accessed |
| | **Study Design** | All types of randomised trials where individuals are studied including<br>• Quasi-randomised trials<br>• Pilot and feasibility trials that use a randomised trial design<br>• Cluster randomised trials | • Quantitative research that does not use randomisation<br>• Qualitative research | 4 |
| **Population** | **Age** | Adults (18 years and above) | • Children (0-17 years)<br>• Child and adult mixed groups where the data from the subset of eligible participants cannot be retrieved | 5 |
| | **Condition** | Diagnosis of stroke (using clinical or radiological criteria) | • Acquired brain injury used as a general term without a specific subgroup for stroke<br>• Transient ischaemic attack<br>• Other neurological conditions<br>• Mixed conditions where the data from the subset of eligible participants cannot be retrieved | 6 |

| | | | | |
|---|---|---|---|---|
| | **Sequalae of stroke** | As a result of the stroke presence of either or both of the following<br>• Impairment of upper limb motor functions (ICF domain B7 impairment of neuromusculoskeletal and movement related function)<br>• Upper limb activity limitation (ICF domain D4 mobility)<br>Note- stroke severity or the presence of other impairment/activity limitation will not influence the inclusion criteria | • Impairment of other body functions or other activity limitations without upper limb motor impairment<br>• Mixed impairment and activity limitation which includes the upper limb but where the data from the subset of eligible participants cannot be retrieved | 7 |
| | **Time since stroke** | Six months or less following stroke (based on the mean-time post stroke recorded in the participant baseline characteristics) | • More than six months post stroke<br>• Mixed time since stroke but where the data from the subset of eligible participants cannot be retrieved | 8<br>8 Mixed |
| **Intervention** | **Digital technology-based rehabilitation intervention** | Digital technology-based rehabilitation intervention delivered over more than one session<br>Electrical stimulation will be included as a digital technology<br>Note-The presence of the digital technology is a core feature but it can sit within a multicomponent intervention | • Single session use<br>• An intervention where the digital technology is not a core part of the intervention e.g., a mobile phone being used solely for phone calls to support an intervention, videos used to deliver action observation | 9 |
| | **Individual** | Delivered to an individual | Delivered to a group | 10 |
| | **Home-based** | The intervention is delivered in the home environment for at least part of the study duration | • Delivered outside the home for the total duration of the study including in other community settings<br>• Unknown intervention location | 11<br>11 Unknown |
| | **Focus** | The intervention: | • Rehabilitation of other impairments and activity limitations | 12 |

| | | | | |
|---|---|---|---|---|
| | | • Must actively reduce upper limb motor impairment and increase activity<br>• Can be delivered alone or as an adjunct to usual therapy<br>• Can be delivered by any intervention provider<br>• Can be supervised or unsupervised<br>• Can be delivered by any mode (e.g. face to face or remotely)<br>• Can be delivered over any period of time using any schedule and for any duration, dose and intensity | • Rehabilitation of a mix of impairments and activity limitations which includes the upper but where the data from the subset of eligible interventions cannot be retrieved<br>• Digital technology which is used for assessment only or as a fully assistive intervention | |
| **Comparator** | | • Usual or conventional therapy<br>• No therapy<br>• Defined placebo intervention | Alternative (non-conventional) therapy such as a different type of technology or variations on a type of technology | 13 |
| **Outcome** | | Planned to assess upper limb measures which address<br>• upper limb motor impairment (ICF domain B7 impairment of neuromusculoskeletal and movement related function) and/or<br>• Upper limb activity limitation (ICF domain D4 mobility) | Outcomes solely addressing<br>• Lower limb impairment and/or activity limitation<br>• Other ICF domains | 14 |

**Abbreviations**

ICF: International Classification of Functioning, Disability and Health[1]

**Notes**

* Eligible conference abstracts and protocols and trial registry entries will be included at title and abstract screening and used to search for full text publications of completed studies

$ Codes for full text screening only

Table S2a-k. Full search strategy for all bibliographic and trial registry databases

| a) Ovid Medline (Search completed 28th April 2025) | | |
|---|---|---|
| 1 | digital health/ or telemedicine/ or remote consultation/ or telerehabilitation/ or therapy, computer assisted/ or digital technology/ | 58738 |
| 2 | computers/ or computer systems/ or exp microcomputers/ or mobile applications/ or exp user-computer interface/ or exp video games/ or gamification/ or computer simulation/ or augmented reality/ or exp virtual reality/ or robotics/ or orthotic devices/ or exp computers, handheld/ or remote sensing technology/ or accelerometry/ or exp cell phone/ or videoconferencing/ or wireless technology/ or exp wearable electronic devices/ or electric stimulation therapy/ or electric stimulation/ or transcutaneous electric nerve stimulation/ or haptic interfaces/ | 597385 |
| 3 | (((digital or health or mobile) adj4 technolog*) or e-health or m-health or ehealth or mhealth or e-therapy or etherapy or mtherapy or m-therapy or erehab* or e-rehab* or mrehab* or m-rehab* or mobile health* or digital health* or digital medicine or telemedicine or tele-medicine or telehealth* or tele-health* or telecare or tele-care or telemanagement or tele-management or telerehab* or tele-rehab* or teleconsult* or tele-consult* or telestroke or tele-stroke or teletherap* or tele-therap* or ((computer-based or internet-based) adj4 (therap* or medicine)) or ((remote or electronic or video or tele or virtual) adj4 (consult* or rehab* or therap* or treatment or physio* or occupational therap* or monitor* or care)) or health informatics or medical informatics or biomedical technolog* or patient portal or digital monitor*).ab,kw,ti. | 164116 |
| 4 | (digital device* or smart-phone* or smart phone* or mobile or iPod or iPad or android or apple or blackberry or tablet* or smart-watch* or smart watch* or computer* or laptop* or PC or microcomputer* or Fitbit or pedometer* or activity tracker* or X-Box or Kinect or Wii or wireless or Nintendo or PlayStation or gam* console* or acceleromet* or inertial measurement unit* or IMU or motion capture or personal digital assistant or activity monitor* or sensor or remote sensing technolog* or (wearable adj4 (device* or sensor* or technolog*)) or interactive technolog* or virtual reality or VR or (virtual adj4 (environment* or object* or system* or program*)) or augmented reality or AR or mixed reality or leap motion or robot* or orthos* or orthot* or computer-aided or computer aided or computer assisted or computer-assisted or electromechanical or electro-mechanical or exoskeleton* or end-effector or ARMin or IntelliArm or NEUROExos or Mit-Manus or ((electr* or musc* or neuromusc* or nerve or neuro) adj4 stimulat*) or electrotherapy or artificial intelligence or AI or haptic* or digital interaction or app*1 or application* or internet or web-based or online or video or YouTube or skype or zoom or e-mail or electronic mail or text messag* or SMS or MMS or Wi-Fi or audio-video or ((tele or video) adj4 conferenc*) or ((serious or applied or health or rehab* or video or interactive or computer) adj4 gam*) or exergam* or gaming).ab,kw,ti. | 3565151 |
| 5 | 1 or 2 or 3 or 4 | 3956027 |
| 6 | physical therapy modalities/ or exercise movement techniques/ or exp exercise therapy/ or exercise/ or "activit* of daily living"/ or recreation therapy/ or "recovery of function"/ or "physical and rehabilitation medicine"/ or rehabilitation/ or neurological rehabilitation/ or stroke rehabilitation/ | 343765 |
| 7 | allied health occupations/ or occupational therapy/ or allied health personnel/ or physical therapist assistants/ or occupational therapists/ or physical therapists/ | 33850 |
| 8 | (rehab* or neurorehab* or neurological rehab* or stroke rehab* or mobili#ation or (activit* adj4 daily living) or functional activit* or functional training or exercis* or training or physiotherap* or physical therap* or recreational therap* or occupational therap* or allied health worker* or allied health professional*).ab,kw,ti. | 1338021 |
| 9 | 6 or 7 or 8 | 1485964 |
| 10 | cerebrovascular disorders/ or exp basal ganglia cerebrovascular disease/ or exp brain ischemia/ or exp carotid artery diseases/ or carotid artery, internal, dissection/ or cerebral small vessel diseases/ or exp intracranial arterial diseases/ or exp cerebral arterial diseases/ or exp "intracranial embolism and thrombosis"/ or intracranial | 493156 |

| | | |
|---|---|---|
| | arteriovenous malformations/ or exp intracranial hemorrhages/ or exp cerebral hemorrhage/ or stroke/ or exp brain infarction/ or stroke, lacunar/ or vasospasm, intracranial/ or brain injuries/ or hemiplegia/ or paresis/ or exp cerebrovasular trauma/ | |
| 11 | (stroke or apoplex* or cva or poststroke or post-stroke or ((cerebr* or brain) adj4 vasc*) or hemipar* or hemipleg* or paresis or paretic or ((brain or cerebr* or cerebell* or hemispher* or intracran* or intracerebral or intercran* or vertebrobasilar or middle cerebral artery or MCA or anterior circulation or posterior circulation or basilar artery or vertebral artery or carotid artery or intraventricular or basal ganglia or internal capsule or subarchnoid or hemispher*) adj4 (isch?emi* or infarct* or thromb* or embol* or occlus* or hypoxi* or h?emorrhag* or h?ematom* or bleed*))).ab,kw,ti. | 543710 |
| 12 | 10 or 11 | 754752 |
| 13 | exp upper extremity/ | 195796 |
| 14 | ((upper adj1 (extremit* or limb*)) or arm*1 or shoulder* or axilla* or hand*1 or elbow* or forearm* or finger* or wrist* or palm* or thumb*).ab,kw,ti. | 1266398 |
| 15 | 13 or 14 | 1311138 |
| 16 | home environment/ or community health services/ or exp community participation/ or home care services/ or hospital to home transition/ | 119808 |
| 17 | (home or home-based or community or outreach or in-home or domiciliary or domicile or domestic or house or household or residence).ab,kw,ti. | 1286563 |
| 18 | 16 or 17 | 1347286 |
| 19 | exp randomized controlled trial/ | 639373 |
| 20 | controlled clinical trial.pt. | 95699 |
| 21 | (randomi#ed or placebo or randomly or trial or groups).ab. | 3953512 |
| 22 | therapy.fs. | 2271939 |
| 23 | 19 or 20 or 21 or 22 | 5996982 |
| 24 | exp animals/ not humans.sh. | 5333638 |
| 25 | 23 not 24 | 5395510 |
| 26 | 5 and 9 and 12 and 15 and 18 and 25 | 385 |
| 27 | limit 26 to yr="2000 -Current" | 385 |
| 28 | limit 27 to english language | 384 |

| b) Ovid EMBASE (Search completed 28th April 2025) | | |
|---|---|---|
| 1 | exp digital health/ or telemedicine/ or teleconsultation/ or telerehabilitation/ or teletherapy/ or computer assisted therapy/ or digital technology/ | 85597 |
| 2 | computer/ or computer system/ or microcomputer/ or exp mobile application/ or computer interface/ or exp video game/ or gamification/ or computer simulation/ or augmented reality/ or virtual reality/ or robotics/ or orthosis/ or personal digital assistant/ or remote sensing/ or accelerometry/ or exp mobile phone/ or videoconferencing/ or wireless communication/ or exp wearable computer/ or electrotherapy/ or nerve stimulation/ or haptic interface/ | 580003 |
| 3 | (((digital or health or mobile) adj4 technolog*) or e-health or m-health or ehealth or mhealth or e-therapy or etherapy or mtherapy or m-therapy or erehab* or e-rehab* or mrehab* or m-rehab* or mobile health* or digital health* or digital medicine or telemedicine or tele-medicine or telehealth* or tele-health* or telecare or tele-care or telemanagement or tele-management or telerehab* or tele-rehab* or teleconsult* or tele-consult* or telestroke or tele-stroke or teletherap* or tele-therap* or ((computer-based or internet-based) adj4 (therap* or medicine)) or ((remote or electronic or video or tele or virtual) adj4 (consult* or rehab* or therap* or treatment or physio* or occupational therap* or monitor* or care)) or health informatics or medical informatics or biomedical technolog* or patient portal or digital monitor*).ab,kw,ti. | 208373 |
| 4 | (digital device* or smart-phone* or smart phone* or mobile or iPod or iPad or android or apple or blackberry or tablet* or smart-watch* or smart watch* or computer* or | 4647961 |

| | | |
|---|---|---|
| | laptop* or PC or microcomputer* or Fitbit or pedometer* or activity tracker* or X-Box or Kinect or Wii or wireless or Nintendo or PlayStation or gam* console* or acceleromet* or inertial measurement unit* or IMU or motion capture or personal digital assistant or activity monitor* or sensor* or remote sensing technolog* or (wearable adj4 (device* or sensor* or technolog*)) or interactive technolog* or virtual reality or VR or (virtual adj4 (environment* or object* or system* or program*)) or augmented reality or AR or mixed reality or leap motion or robot* or orthos* or orthot* or computer-aided or computer aided or computer assisted or computer-assisted or electromechanical or electro-mechanical or exoskeleton* or end-effector or ARMin or IntelliArm or NEUROExos or Mit-Manus or ((electr* or musc* or neuromusc* or nerve or neuro) adj4 stimulat*) or electrotherapy or artificial intelligence or AI or haptic* or digital interaction or app*1 or application* or internet or web-based or online or video or YouTube or skype or zoom or e-mail or electronic mail or text messag* or SMS or MMS or Wi-Fi or audio-video or ((tele or video) adj4 conferenc*) or ((serious or applied or health or rehab* or video or interactive or computer) adj4 gam*) or exergam* or gaming).ab,kw,ti. | |
| 5 | 1 or 2 or 3 or 4 | 4977941 |
| 6 | exercise movement techniques/ or exp kinesiotherapy/ or exercise/ or daily life activity/ or recreation therapy/ or convalescence/ or rehabilitation/ or neurorehabilitation/ or stroke rehabilitation/ or rehabilitation medicine/ | 755699 |
| 7 | occupational therapy/ or physiotherapy/ or physiotherapist assistant/ or occupational therapist/ or physiotherapist/ | 164602 |
| 8 | (rehab* or neurorehab* or neurological rehab* or stroke rehab* or mobili#ation or (activit* adj4 daily living) or functional activit* or functional training or exercis* or training or physiotherap* or physical therap* or recreational therap* or occupational therap* or allied health worker* or allied health professional*).ab,kw,ti. | 1766395 |
| 9 | 6 or 7 or 8 | 2072205 |
| 10 | cerebrovascular disease/ or exp basal ganglion hemorrhage/ or exp brain hematoma/ or exp brain hemorrhage/ or exp brain infarction/ or exp brain ischemia/ or exp carotid artery disease/ or vertebral artery dissection/ or exp cerebral artery disease/ or exp cerebrovascular accident/ or exp ischemic stroke/ or cerebral atherosclerosis/ or brain arteriovenous malformation/ or thromboembolism/ or exp intracranial aneurysm/ or exp occlusive cerebrovascular disease/ or vertebrobasilar insufficiency/ or brain vasospasm/ or brain injury/ or hemiplegia/ or hemiparesis/ | 1133672 |
| 11 | (stroke or apoplex* or cva or poststroke or post-stroke or ((cerebr* or brain) adj4 vasc*) or hemipar* or hemipleg* or paresis or paretic or ((brain or cerebr* or cerebell* or hemispher* or intracran* or intracerebral or intercran* or vertebrobasilar or middle cerebral artery or MCA or anterior circulation or posterior circulation or basilar artery or vertebral artery or carotid artery or intraventricular or basal ganglia or internal capsule or subarchnoid or hemispher*) adj4 (isch?emi* or infarct* or thromb* or embol* or occlus* or hypoxi* or h?emorrhag* or h?ematom* or bleed*))).ab,kw,ti. | 815520 |
| 12 | 10 or 11 | 1324517 |
| 13 | exp upper limb/ or exp arm/ or exp finger/ or exp shoulder/ | 386913 |
| 14 | ((upper adj1 (extremit* or limb*)) or arm*1 or shoulder* or axilla* or hand*1 or elbow* or forearm* or finger* or wrist* or palm* or thumb*).ab,kw,ti. | 1704964 |
| 15 | 13 or 14 | 1786625 |
| 16 | home environment/ or home rehabilitation/ or home physiotherapy/ or community participation/ or home care/ or hospital to home transition/ | 89366 |
| 17 | (home or home-based or community or outreach or in-home or domiciliary or domicile or domestic or house or household or residence).ab,kw,ti. | 1652652 |
| 18 | 16 or 17 | 1675358 |
| 19 | exp randomized controlled trial/ or controlled clinical trial/ or randomization/ or intermethod comparison/ or double blind procedure/ or human experiment/ | 2083978 |
| 20 | (random* or placebo or compare or compared or comparison or (open adj label) or ((double or single or doubly or singly) adj (blind or blinded or blindly)) or parallel | 10175542 |

| | | |
|---|---|---|
| | group*1 or crossover or cross over or ((assign* or match or matched or allocation) adj5 (alternate or group*1 or intervention*1 or patient*1 or subject*1 or participant*1)) or assigned or allocated or (controlled adj7 (study or design or trial)) or volunteer or volunteers).ti,ab. | |
| 21 | ((evaluated or evaluate or evaluating or assessed or assess) and (compare or compared or comparing or comparison)).ab. | 3123865 |
| 22 | trial.ti. | 455407 |
| 23 | 19 or 20 or 21 or 22 | 11002263 |
| 24 | (random* adj sampl* adj7 ("cross section*" or questionnaire*1 or survey* or database*1)).ti,ab. not (comparative study/ or controlled study/ or randomi#ed controlled.ti,ab. or randomly assigned.ti,ab.) | 10410 |
| 25 | cross-sectional study/ not (exp randomized controlled trial/ or controlled clinical trial/ or controlled study/ or randomi?ed controlled.ti,ab. or control group*1.ti,ab.) | 438502 |
| 26 | (((case adj control$) and random$) not randomi?ed controlled).ti,ab. | 23474 |
| 27 | systematic review.ti,ab. not (trial or study).ti. | 398808 |
| 28 | (non random* not random*).ti,ab. | 0 |
| 29 | random field*.ti,ab. | 3150 |
| 30 | (random cluster adj3 sample*).ti,ab. | 235 |
| 31 | (review.ab. and review.pt.) not trial.ti. | 1279193 |
| 32 | we searched.ab. and (review.ti. or review.pt.) | 57693 |
| 33 | update review.ab. | 153 |
| 34 | (databases adj4 searched).ab. | 75720 |
| 35 | (rat or rats or mouse or mice or swine or porcine or murine or sheep or lambs or pigs or piglets or rabbit or rabbits or cat or cats or dog or dogs or cattle or bovine or monkey or monkeys or trout or marmoset*1).ti. and animal experiment/ | 1297403 |
| 36 | animal experiment/ not (human experiment/ or human/) | 2735373 |
| 37 | or/24-36 | 4824048 |
| 38 | 23 not 37 | 9592571 |
| 39 | 5 and 9 and 12 and 15 and 18 and 38 | 613 |
| 40 | limit 39 to yr="2000 -Current" | 611 |
| 41 | limit 40 to english language | 606 |

| c) Ebsco CINAHL (Search completed 28th April 2025) | | | |
|---|---|---|---|
| S1 | (MH "Digital Health") OR (MH "Telehealth") OR (MH "Telemedicine") OR (MH "Remote Consultation") OR (MH "Telerehabilitation") OR (MH "Digital Technology") OR (MH "Therapy, Computer Assisted") | Expanders - Apply equivalent subjects. Search modes - Proximity | 49339 |
| S2 | (MH "Video Games+") OR (MH "Virtual Reality+") OR (MH "Mobile Applications") OR (MH "Web Browsers") OR (MH "Minicomputers") OR (MH "Smart Glasses") OR (MH "Smartphone") OR (MH "Computers, Hand-Held") OR (MH "Computers, Portable") OR (MH "Microcomputers") OR (MH "Computers, Mainframe") OR (MH "Communications Software") OR (MH "User-Computer Interface") OR (MH "Games+") OR (MH "Augmented Reality") OR (MH "Computer Simulation") OR (MH "Robotics+") OR (MH "Orthoses") OR (MH "Accelerometry") OR (MH "Electrical Stimulation, Functional") OR (MH "Electrical Stimulation, Neuromuscular") OR (MH "Electric Stimulation") OR (MH "Cellular Phone") OR (MH "Videoconferencing") OR (MH | Expanders - Apply equivalent subjects. Search modes - Proximity | 124095 |

| | | | |
|---|---|---|---|
| | "Text Messaging") OR (MH "Wearable Sensors") OR (MH "Accelerometers") | | |
| S3 | XB ((digital or health or mobile) N4 technolog*) or e-health or m-health or ehealth or mhealth or e-therapy or etherapy or mtherapy or m-therapy or erehab* or e-rehab* or mrehab* or m-rehab* or “mobile health*” or “digital health*” or “digital medicine” or telemedicine or tele-medicine or telehealth* or tele-health* or telecare or tele-care or telemanagement or tele-management or telerehab* or tele-rehab* or teleconsult* or tele-consult* or telestroke or tele-stroke or teletherap* or tele-therap* or ((computer-based or internet-based or “computer assisted”) N4 (therap* or medicine)) or ((remote or electronic or video or tele or virtual) N4 (consult* or rehab* or therap* or treatment or physio* or "occupational therap*" or monitor* or care)) or “health informatics” or “medical informatics” or “biomedical technolog*” or “patient portal” or “digital monitor*”) | Expanders - Apply equivalent subjects. Search modes - Proximity | 110293 |
| S4 | XB (“digital device*” or smart-phone* or “smart phone*” or mobile or iPod or iPad or android or apple or blackberry or tablet* or smart-watch* or “smart watch*” or computer* or laptop* or PC or microcomputer* or Fitbit or pedometer* or “activity tracker*" or X-Box or Kinect or Wii or wireless or Nintendo or PlayStation or “gam* console*” or acceleromet* or “inertial measurement unit*” or IMU or “motion capture” or “personal digital assistant” or “activity monitor*” or sensor* or “remote sensing technolog*” or (wearable N4 (device* or sensor* or technolog*)) or “interactive technolog*” or “virtual reality” or VR or (virtual N4 (environment* or object* or system* or program*)) or “augmented reality” or AR or “mixed reality” or “leap motion” or robot* or orthos* or orthot* or computer-aided or “computer aided” or “computer assisted” or computer-assisted or electromechanical or electro-mechanical or exoskeleton* or end-effector or ARMin or IntelliArm or NEUROExos or Mit-Manus or ((electr* or musc* or neuromusc* or nerve or neuro) N4 stimulat*) or electrotherapy or “artificial intelligence” or AI or haptic* or “digital interaction” or app# or application# or internet or web-based or online or video or YouTube or skype or zoom or e-mail or “electronic mail” or “text messag*” or SMS or MMS or Wi-Fi or audio-video or ((tele or video) N4 conferenc*) or ((serious or applied or health or rehab* or video or interactive or computer) N4 gam*) or exergam* or gaming) | Expanders - Apply equivalent subjects. Search modes - Proximity | 639225 |
| S5 | S1 OR S2 OR S3 OR S4 | Expanders - Apply equivalent subjects. Search modes - Proximity | 761660 |
| S6 | (MH "Occupational Therapy") OR (MH "Physical Therapy") OR (MH Physical Therapy Assisting) OR (MH "Allied Health Professions") OR (MH "Physical Therapist Assistants") OR (MH "Occupational Therapy Assistants") OR (MH "Occupational Therapists") OR (MH "Physical Therapists") | Expanders - Apply equivalent subjects. Search modes - Proximity | 85331 |

| | | | |
|---|---|---|---|
| S7 | (MH "Activities of Daily Living") OR (MH "Rehabilitation") OR (MH "Therapeutic Exercise+") OR (MH "Recreational Therapy") | Expanders - Apply equivalent subjects. Search modes - Proximity | 129166 |
| S8 | XB (rehab* or neurorehab* or "neurological rehab*" or "stroke rehab*" or mobili?ation or (activit* N4 "daily living") or "functional activit*" or "functional training" or exercis* or training or physiotherap* or "physical therap*" or "recreational therap*" or "occupational therap*" or "allied health worker*" or "allied health professional*") | Expanders - Apply equivalent subjects. Search modes - Proximity | 548648 |
| S9 | S6 OR S7 OR S8 | Expanders - Apply equivalent subjects. Search modes - Proximity | 634686 |
| S10 | (MH "Cerebrovascular Disorders") OR (MH "Basal Ganglia Cerebrovascular Disease+") OR (MH "Hypoxia-Ischemia, Brain") OR (MH "Carotid Artery Diseases+") OR (MH "Cerebral Ischemia") OR (MH "Intracranial Embolism and Thrombosis+") OR (MH "Intracranial Hemorrhage+") OR (MH "Stroke+") OR (MH "Vertebral Artery Dissections") OR (MH "Brain Injuries") OR (MH "Hemiplegia") OR (MH "Cerebral Arterial Diseases+") OR (MH "Intracranial Arterial Diseases") OR (MH "Cerebral Small Vessel Diseases") OR (MH "Stroke, Lacunar") OR (MH "Cerebral Vasospasm") OR (MH "Cerebral Ischemia, Transient") | Expanders - Apply equivalent subjects. Search modes - Proximity | 155370 |
| S11 | XB (stroke or apoplex* or cva or poststroke or post-stroke or ((cerebr* or brain) N4 vasc*) or hemipar* or hemipleg* or paresis or paretic or ((brain or cerebr* or cerebell* or hemispher* or intracran* or intracerebral or intercran* or vertebrobasilar or "middle cerebral artery" or MCA or "anterior circulation" or "posterior circulation" or "basilar artery" or "vertebral artery" or "carotid artery" or intraventricular or "basal ganglia" or "internal capsule" or subarchnoid or hemispher*) N4 (isch#emi* or infarct* or thromb* or embol* or occlus* or hypoxi* or h#emorrhag* or h#ematom* or bleed*))) | Expanders - Apply equivalent subjects. Search modes - Proximity | 155240 |
| S12 | S10 OR S11 | Expanders - Apply equivalent subjects. Search modes - Proximity | 220695 |
| S13 | (MH "Upper Extremity+") | Expanders - Apply equivalent subjects. Search modes - Proximity | 48334 |
| S14 | XB ((upper N1 (extremit* or limb*)) or arm# or shoulder* or axilla* or hand# or elbow* or forearm* or finger* or wrist* or palm* or thumb*) | Expanders - Apply equivalent subjects. Search modes - Proximity | 241508 |
| S15 | S13 OR S14 | Expanders - Apply equivalent subjects. Search modes - Proximity | 252177 |
| S16 | (MH "Home Health Care") OR (MH "Rehabilitation, Community-Based") OR (MH "Home Rehabilitation+") OR (MH "Home Visits") OR (MH "Home Environment") OR (MH "Hospital to Home Transition") | Expanders - Apply equivalent subjects. Search modes - Proximity | 51383 |

| | | | |
|---|---|---|---|
| S17 | XB (home or home-based or community or outreach or in-home or domiciliary or domicile or domestic or house or household or residence) | Expanders - Apply equivalent subjects. Search modes - Proximity | 561262 |
| S18 | S16 OR S17 | Expanders - Apply equivalent subjects. Search modes - Proximity | 576944 |
| S19 | (MH "Double-Blind Studies") OR (MH "Single-Blind Studies") OR (MH "Randomized Controlled Trials+") OR (MH "Pretest-Posttest Design") OR (MH "Random Assignment") OR (MH "Cluster Sample") OR (MH "Placebos") OR (MH "Crossover Design") OR (MH "Comparative Studies") | Expanders - Apply equivalent subjects. Search modes - Proximity | 707830 |
| S20 | TI (randomised OR randomized OR trial) | Expanders - Apply equivalent subjects. Search modes - Proximity | 236197 |
| S21 | AB random* OR (cluster W3 RCT) OR (control W5 group) | Expanders - Apply equivalent subjects. Search modes - Proximity | 515383 |
| S22 | MH Sample Size AND AB (assigned OR allocated OR control) | Expanders - Apply equivalent subjects. Search modes - Proximity | 4516 |
| S23 | PT "randomized controlled trial" | Expanders - Apply equivalent subjects. Search modes - Proximity | 163148 |
| S24 | (MH "Animals+") OR (MH "Animal Studies") | Expanders - Apply equivalent subjects. Search modes - Proximity | 247238 |
| S25 | TI "animal model*" | Expanders - Apply equivalent subjects. Search modes - Proximity | 3746 |
| S26 | S24 OR S25 | Expanders - Apply equivalent subjects. Search modes - Proximity | 248494 |
| S27 | (MH "Human") | Expanders - Apply equivalent subjects. Search modes - Proximity | 2906082 |
| S28 | S26 NOT S27 | Expanders - Apply equivalent subjects. Search modes - Proximity | 213758 |
| S29 | S19 OR S20 OR S21 OR S22 OR S23 | Expanders - Apply equivalent subjects. Search modes - Proximity | 1102780 |
| S30 | S29 NOT S28 | Expanders - Apply equivalent subjects. | 1053933 |

| | | | |
|---|---|---|---|
| | | Search modes - Proximity | |
| S31 | S5 AND S9 AND S12 AND S15 AND S18 AND S30 | Expanders - Apply equivalent subjects. Search modes - Proximity | 195 |
| S32 | S31 | Limiters - Publication Year: 2000-. Expanders - Apply equivalent subjects. Search modes - Proximity | 195 |
| S33 | S32 | Limiters - English language; Language: English. Expanders - Apply equivalent subjects. Search modes - Proximity | 193 |

| d) Ebsco PyscINFO (Search completed 28th April 2025) | | | |
|---|---|---|---|
| S1 | DE "Electronic Health Services" OR DE "Telemedicine" OR DE "Telerehabilitation" OR DE "Computer Assisted Therapy" OR DE "Digital Technology" OR DE "Digital Interventions" | Expanders - Apply equivalent subjects. Search modes - Proximity | 20,630 |
| S2 | DE "Human Robot Interaction" OR DE "Mobile Applications" OR DE "Mobile Health Applications" OR DE "Text Messaging" OR DE "Mobile Devices" OR DE "Mobile Phones" OR DE "Tablet Computers" OR DE "Digital Computers" OR DE "Microcomputers" OR DE "Robotics" OR DE "Online Therapy" OR DE "Teleconferencing" OR DE "Videoconferencing" OR DE "Teleconsultation" OR DE "Digital Game-Based Learning" OR DE "Digital Gaming" OR DE "Gamification" OR DE "Virtual Environment" OR DE "Computer Games" OR DE "Avatars" OR DE "Computers" OR DE "Laptop Computers" OR DE "Virtual Reality" OR DE "Augmented Reality" OR DE "Internet" OR DE "Smartphones" OR DE "Wireless Technologies" OR DE "Wearable Devices" OR DE “Human Computer Interaction” OR DE "Sensor Technology" OR DE "Electrical Stimulation" OR DE "Nerve Stimulation" | Expanders - Apply equivalent subjects. Search modes - Proximity | 132,037 |
| S3 | XB (((digital or health or mobile) N4 technolog*) or e-health or m-health or ehealth or mhealth or e-therapy or etherapy or mtherapy or m-therapy or erehab* or e-rehab* or mrehab* or m-rehab* or “mobile health*” or “digital health*” or “digital medicine” or telemedicine or tele-medicine or telehealth* or tele-health* or telecare or tele-care or telemanagement or tele-management or telerehab* or tele-rehab* or teleconsult* or tele-consult* or telestroke or tele-stroke or teletherap* or tele-therap* or ((computer-based or internet-based or “computer assisted”) N4 (therap* or medicine)) or ((remote or electronic or video or tele or virtual) N4 (consult* or rehab* or therap* or treatment or physio* or "occupational therap*" or monitor* or care)) or “health informatics” or “medical informatics” or “biomedical technolog*” or “patient portal” or “digital monitor*”) | Expanders - Apply equivalent subjects. Search modes - Proximity | 38,541 |

| | | | |
|---|---|---|---|
| S4 | XB ("digital device*" or smart-phone* or "smart phone*" or mobile or iPod or iPad or android or apple or blackberry or tablet* or smart-watch* or "smart watch*" or computer* or laptop* or PC or microcomputer* or Fitbit or pedometer* or "activity tracker*" or X-Box or Kinect or Wii or wireless or Nintendo or PlayStation or "gam* console*" or acceleromet* or "inertial measurement unit*" or IMU or "motion capture" or "personal digital assistant" or "activity monitor*" or sensor* or "remote sensing technolog*" or (wearable N4 (device* or sensor* or technolog*)) or "interactive technolog*" or "virtual reality" or "VR" or (virtual N4 (environment* or object* or system* or program*)) or "augmented reality" or "AR" or "mixed reality" or "leap motion" or robot* or orthos*or orthot* or computer-aided or "computer aided" or "computer assisted" or computer-assisted or electromechanical or electro-mechanical or exoskeleton* or end-effector or ARMin or IntelliArm or NEUROExos or Mit-Manus or ((electr* or musc* or neuromusc* or nerve or neuro) N4 stimulat*) or electrotherapy or "artificial intelligence" or "AI" or haptic* or "digital interaction" or app# or application# or internet or web-based or online or video or YouTube or skype or zoom or e-mail or "electronic mail" or "text messag*" or SMS or MMS or Wi-Fi or audio-video or ((tele or video) N4 conferenc*) or ((serious or applied or health or rehab* or video or interactive or computer) N4 gam*) or exergam* or gaming) | Expanders - Apply equivalent subjects. Search modes - Proximity | 727,755 |
| S5 | S1 OR S2 OR S3 OR S4 | Expanders - Apply equivalent subjects. Search modes - Proximity | 762,657 |
| S6 | DE "Exercise" OR DE "Physical Activity" OR DE "Recreation Therapy" OR DE "Movement Therapy" OR DE "Physical Treatment Methods" OR DE "Exercise Therapy" OR DE "Activities of Daily Living" OR DE "Neurorehabilitation" OR DE "Rehabilitation" | Expanders - Apply equivalent subjects. Search modes - Proximity | 110,612 |
| S7 | DE "Allied Health Personnel" DE "Occupational Therapy" OR DE "Physical Therapy" | Expanders - Apply equivalent subjects. Search modes - Proximity | 4,028 |
| S8 | XB (rehab* or neurorehab* or "neurological rehab*" or "stroke rehab*" or mobili?ation or (activit* N4 "daily living") or "functional activit*" or "functional training" or exercis* or training or physiotherap* or "physical therap*" or "recreational therap*" or "occupational therap*" or "allied health worker*" or "allied health professional*") | Expanders - Apply equivalent subjects. Search modes - Proximity | 481,251 |
| S9 | S6 OR S7 OR S8 | Expanders - Apply equivalent subjects. Search modes - Proximity | 527,517 |
| S10 | DE "Brain Injuries" OR DE "Brain Disorders" OR DE "Cerebral Hemorrhage" OR DE "Subarachnoid Hemorrhage" OR DE "Cerebral Ischemia" OR DE "Cerebral Infarction" OR DE "Cerebral Small Vessel Disease" OR DE "Cerebrovascular Disorders" OR DE "Cerebral Small Vessel Disease" OR DE | Expanders - Apply equivalent subjects. Search modes - Proximity | 57,646 |

| | | | |
|---|---|---|---|
| | "Cerebrovascular Accidents" OR DE "Hemiplegia" OR DE "Hemiparesis" OR DE "Cerebral Arteriosclerosis" | | |
| S11 | XB (stroke or apoplex* or cva or poststroke or post-stroke or ((cerebr* or brain) N4 vasc*) or hempar* or hemipleg* or paresis or paretic or ((brain or cerebr* or cerebell* or hemispher* or intracran* or intracerebral or intercran* or vertebrobasilar or "middle cerebral artery" or MCA or "anterior circulation" or "posterior circulation" or "basilar artery" or "vertebral artery" or "carotid artery" or intraventricular or "basal ganglia" or "internal capsule" or subarchnoid or hemispher*) N4 (isch#emi* or infarct* or thromb* or embol* or occlus* or hypoxi* or h#emorrhag* or h#ematom* or bleed*))) | Expanders - Apply equivalent subjects. Search modes - Proximity | 57,739 |
| S12 | S10 OR S11 | Expanders - Apply equivalent subjects. Search modes - Proximity | 80,951 |
| S13 | DE "Arm (Anatomy)" OR DE "Elbow (Anatomy)" OR DE "Shoulder (Anatomy)" OR DE "Wrist" | Expanders - Apply equivalent subjects. Search modes - Proximity | 4,473 |
| S14 | XB ((upper N1 (extremit* or limb*)) or arm# or shoulder* or axilla* or hand# or elbow* or forearm* or finger* or wrist* or palm* or thumb*) | Expanders - Apply equivalent subjects. Search modes - Proximity | 175,327 |
| S15 | S13 OR S14 | Expanders - Apply equivalent subjects. Search modes - Proximity | 175,570 |
| S16 | DE "Home Care" OR DE "Hospital Discharge" OR DE "Community Services" | Expanders - Apply equivalent subjects. Search modes - Proximity | 30,320 |
| S17 | XB (home or home-based or community or outreach or in-home or domiciliary or domicile or domestic or house or household or residence) | Expanders - Apply equivalent subjects. Search modes - Proximity | 584,664 |
| S18 | S16 OR S17 | Expanders - Apply equivalent subjects. Search modes - Proximity | 590,471 |
| S19 | DE "Between Groups Design" OR DE "Clinical Trials" OR DE "Random Sampling" OR DE "Randomized Controlled Trials" OR DE "Randomized Clinical Trials" OR DE "Treatment Effectiveness Evaluation" OR DE "Placebo" OR DE "Experimental Subjects" | Expanders - Apply equivalent subjects. Search modes - Proximity | 55,907 |
| S20 | XB (double-blind or single-blind or randomi?ed or random* or control* or trial* or "pretest-posttest design" or "random assignment" or group* or placebo* or "crossover design" or "comparative stud*" or therapy) | Expanders - Apply equivalent subjects. Search modes - Proximity | 2,014,620 |
| S21 | S19 OR S20 | Expanders - Apply equivalent subjects. Search modes - Proximity | 2,026,636 |
| S22 | S5 AND S9 AND S12 AND S15 AND S18 AND S21 | Expanders - Apply equivalent subjects. | 127 |

| | | | |
|---|---|---|---|
| | | Search modes - Proximity | |
| S23 | S22 | Limiters - Publication Year: 2000-. Expanders - Apply equivalent subjects. Search modes - Proximity | 127 |
| S24 | S23 | Limiters - English language; Language: English. Expanders - Apply equivalent subjects. Search modes - Proximity | 125 |

| e) Ebsco AMED (Search completed 28th April 2025) | | | |
|---|---|---|---|
| S1 | ((((ZU "telemedicine") or (ZU "telerehabilitation")) or ((ZU "remote consultation"))) or ((ZU "computer assisted therapy"))) or ((ZU "technology")) | Expanders - Apply equivalent subjects. Search modes - Proximity | 1,970 |
| S2 | ((((((ZU "computers") or (ZU "computer systems") or (ZU "microcomputers") or (ZU "mobile applications") or (ZU "user computer interface") or (ZU "video games") or (ZU "computer simulation") or (ZU "virtual reality") or (ZU "robotics") or (ZU "orthotic devices") or (ZU "computers handheld") or (ZU "smartphone") or (ZU "smartphones") or (ZU "text messaging") or (ZU "fitness trackers")) or ((ZU "wireless technology") or (ZU "accelerometry") or (ZU "cell phone") or (ZU "cell phones"))) or ((ZU "videoconferencing"))) or ((ZU "wearable electronic devices"))) or ((ZU "electric stimulation therapy"))) or ((ZU "electric stimulation")) | Expanders - Apply equivalent subjects. Search modes - Proximity | 8,010 |
| S3 | ((digital or health or mobile) N4 technolog*) or e-health or m-health or ehealth or mhealth or e-therapy or etherapy or mtherapy or m-therapy or erehab* or e-rehab* or mrehab* or m-rehab* or “mobile health*” or “digital health*” or “digital medicine” or telemedicine or tele-medicine or telehealth* or tele-health* or telecare or tele-care or telemanagement or tele-management or telerehab* or tele-rehab* or teleconsult* or tele-consult* or telestroke or tele-stroke or teletherap* or tele-therap* or ((computer-based or internet-based or “computer assisted”) N4 (therap* or medicine)) or ((remote or electronic or video or tele or virtual) N4 (consult* or rehab* or therap* or treatment or physio* or "occupational therap*" or monitor* or care)) or “health informatics” or “medical informatics” or “biomedical technolog*” or “patient portal” or “digital monitor*” | Expanders - Apply equivalent subjects. Search modes - Proximity | 3,246 |
| S4 | “digital device*” or smart-phone* or “smart phone*” or mobile or iPod or iPad or android or apple or blackberry or tablet* or smart-watch* or “smart watch*” or computer* or laptop* or PC or microcomputer* or Fitbit or pedometer* or “activity tracker*” or X-Box or Kinect or Wii or wireless or Nintendo or PlayStation or “gam* console*” or acceleromet* or “inertial measurement unit*” or IMU or “motion capture” or “personal digital assistant” or “activity | Expanders - Apply equivalent subjects. Search modes - Proximity | 23,560 |

| | | | |
|---|---|---|---|
| | monitor*" or sensor or "remote sensing technolog*" or (wearable N4 (device* or sensor* or technolog*)) or "interactive technolog*" or "virtual reality" or VR or (virtual N4 (environment* or object* or system* or program*)) or "augmented reality" or "AR" or "mixed reality" or "leap motion" or robot* or orthos*or orthot* or computer-aided or "computer aided" or "computer assisted" or computer-assisted or electromechanical or electro-mechanical or exoskeleton* or end-effector or ARMin or IntelliArm or NEUROExos or Mit-Manus or ((electr* or musc* or neuromusc* or nerve or neuro) N4 stimulat*) or electrotherapy or "artificial intelligence" or AI or haptic* or "digital interaction" or app# or application#" or internet or web-based or online or video or YouTube or skype or zoom or e-mail or "electronic mail" or "text messag*" or SMS or MMS or Wi-Fi or audio-video or ((tele or video) N4 conferenc*) or ((serious or applied or health or rehab* or video or interactive or computer) N4 gam*) or exergam* or gaming | | |
| S5 | S1 OR S2 OR S3 OR S4 | Expanders - Apply equivalent subjects. Search modes - Proximity | 27,683 |
| S6 | ((((((((ZU "physical therapy modalities") or (ZU "exercise") or (ZU "exercise movement techniques") or (ZU "exercise therapy")) or ((ZU "endurance training") or (ZU "muscle stretching exercises") or (ZU "plyometric exercise") or (ZU "resistance training"))) or ((ZU "activities of daily living"))) or ((ZU "recreation therapy"))) or ((ZU "recovery of function"))) or ((ZU "physical and rehabilitation medicine"))) or ((ZU "rehabilitation"))) or ((ZU "stroke rehabilitation"))) or ((ZU "neurological rehabilitation")) | Expanders - Apply equivalent subjects. Search modes - Proximity | 85,925 |
| S7 | (((ZU "allied health") or (ZU "allied health personnel")) or ((ZU "occupational therapy"))) or ((ZU "physical therapy speciality") or (ZU "physical therapists") or (ZU "occupational therapists")) | Expanders - Apply equivalent subjects. Search modes - Proximity | 10,114 |
| S8 | rehab* or neurorehab* or "neurological rehab*" or "stroke rehab*" or mobili?ation or (activit* N4 "daily living") or "functional activit*" or "functional training" or exercis* or training or physiotherap* or "physical therap*" or "recreational therap*" or "occupational therap*" or "allied health worker*" or "allied health professional*" | Expanders - Apply equivalent subjects. Search modes - Proximity | 150,045 |
| S9 | S6 OR S7 OR S8 | Expanders - Apply equivalent subjects. Search modes - Proximity | 150,652 |
| S10 | (((ZU "cerebrovascular accident") or (ZU "cerebrovascular disorders") or (ZU "cerebral arterial diseases") or (ZU "cerebral arteriosclerosis") or (ZU "cerebral hemorrhage") or (ZU "cerebral infarction") or (ZU "cerebral ischemia") or (ZU "cerebral ischemia transient") or (ZU "cerebral small vessel diseases")) or ((ZU "brain injuries") or (ZU "brain ischemia") or (ZU "brain infarction") or (ZU "hypoxia ischemia brain") or (ZU "carotid artery diseases") or (ZU "carotid artery injuries") or (ZU "carotid artery internal dissection") or (ZU "moyamoya disease") or (ZU | Expanders - Apply equivalent subjects. Search modes - Proximity | 15,780 |

| | | | |
|---|---|---|---|
| | "intracranial aneurysm") or (ZU "subarachnoid haemorhage") or (ZU "subarachnoid hemorrhage") or (ZU "stroke") or (ZU "vertebral artery dissection") or (ZU "hemiparesis") or (ZU "hemiplegia") or (ZU "paresis"))) or ((ZU "infarction middle cerebral artery") or (ZU "basal ganglia diseases")) | | |
| S11 | stroke or apoplex* or cva or poststroke or post-stroke or ((cerebr* or brain) N4 vasc*) or hemipar* or hemipleg* or paresis or paretic or ((brain or cerebr* or cerebell* or hemispher* or intracran* or intracerebral or intercran* or vertebrobasilar or “middle cerebral artery” or MCA or “anterior circulation” or “posterior circulation” or “basilar artery” or “vertebral artery” or “carotid artery” or intraventricular or “basal ganglia” or “internal capsule” or subarchnoid or hemispher*) N4 (isch#emi* or infarct* or thromb* or embol* or occlus* or hypoxi* or h#emorrhag* or h#ematom* or bleed*)) | Expanders - Apply equivalent subjects. Search modes - Proximity | 12,962 |
| S12 | S10 OR S11 | Expanders - Apply equivalent subjects. Search modes - Proximity | 18,137 |
| S13 | (((ZU "arm")) or ((ZU "axilla") or (ZU "elbow") or (ZU "forearm") or (ZU "wrist") or (ZU "hand") or (ZU "shoulder") or (ZU "fingers"))) or ((ZU "thumb")) | Expanders - Apply equivalent subjects. Search modes - Proximity | 7,980 |
| S14 | (upper N1 (extremit* or limb*)) or arm# or shoulder* or axilla* or hand# or elbow* or forearm* or finger* or wrist* or palm* or thumb* | Expanders - Apply equivalent subjects. Search modes - Proximity | 24,879 |
| S15 | S13 OR S14 | Expanders - Apply equivalent subjects. Search modes - Proximity | 24,879 |
| S16 | (((ZU "community health services")) or ((ZU "community participation") or (ZU "patient participation"))) or ((ZU "home care services")) | Expanders - Apply equivalent subjects. Search modes - Proximity | 5,367 |
| S17 | home or home-based or community or outreach or in-home or domiciliary or domicile or domestic or house or household or residence | Expanders - Apply equivalent subjects. Search modes - Proximity | 27,174 |
| S18 | S16 OR S17 | Expanders - Apply equivalent subjects. Search modes - Proximity | 28,123 |
| S19 | double-blind or single-blind or randomi?ed or random* or control* or trial* or “pretest-posttest design” or “random assignment” or group* or placebo* or "crossover design" or “comparative stud*" or therapy | Expanders - Apply equivalent subjects. Search modes - Proximity | 197,726 |
| S20 | ((((ZU "randomized controlled trials")) or ((ZU "placebos"))) or ((ZU "comparative study"))) or ((ZU "control groups")) | Expanders - Apply equivalent subjects. Search modes - Proximity | 9,587 |
| S21 | S19 OR S20 | Expanders - Apply equivalent subjects. | 197,726 |

| | | | |
|---|---|---|---|
| | | Search modes - Proximity | |
| S22 | S5 AND S9 AND S12 AND S15 AND S18 AND S21 | Expanders - Apply equivalent subjects. Search modes - Proximity | 115 |
| S23 | S22 | Limiters - Publication Year: 2000-. Expanders - Apply equivalent subjects. Search modes - Proximity | 112 |
| S24 | S23 | Limiters - English language; Language: English. Expanders - Apply equivalent subjects. Search modes - Proximity | 110 |

| f) Cochrane (Search completed 28th April 2025) | | |
|---|---|---|
| #1 | [mh ^"digital health"] or [mh ^telemedicine] or [mh ^"remote consultation"] or [mh ^telerehabilitation] or [mh ^"therapy, computer assisted"] or [mh ^"digital technology"] | 6664 |
| #2 | [mh ^computers] or [mh ^"computer systems"] or [mh microcomputers] or [mh ^"mobile applications"] or [mh "user-computer interface"] or [mh "video games"] or [mh ^"gamification"] or [mh ^"computer simulation"] or [mh ^"augmented reality"] or [mh "virtual reality"] or [mh ^robotics] or [mh ^"orthotic devices"] or [mh "computers, handheld"] or [mh ^"remote sensing technology"] or [mh ^accelerometry] or [mh "cell phone"] or [mh videoconferencing] or [mh ^"wireless technology"] or [mh "wearable electronic devices"] or [mh ^"electric stimulation therapy"] or [mh ^"electric stimulation"] or [mh ^"transcutaneous electric nerve stimulation"] or [mh ^"haptic interface"] | 23065 |
| #3 | (((digital or health or mobile) NEAR/4 technolog*) or e-health or m-health or ehealth or mhealth or e-therapy or etherapy or mtherapy or m-therapy or erehab* or e-rehab* or mrehab* or m-rehab* or (mobile NEXT health*) or (digital NEXT health*) or "digital medicine" or telemedicine or tele-medicine or telehealth* or tele-health* or telecare or tele-care or telemanagement or tele-management or telerehab* or tele-rehab* or teleconsult* or tele-consult* or telestroke or tele-stroke or teletherap* or tele-therap* or (remote NEAR/4 (rehab* or therap* or treatment or physio*)) or (video NEAR/4 (rehab* or therap* or treatment or physio*)) or (tele NEAR/4 (rehab* or therap* or treatment or physio*)) or (virtual NEAR/4 (rehab* or therap* or treatment or physio*)) or "health informatics" or "medical informatics" or ((computer or internet) NEAR/4 therap*) or (biomedical NEXT technolog*) or "patient portal" or (digital NEXT monitor*)):ti,ab,kw | 32172 |
| #4 | ((digital NEXT device*) or (smart NEXT phone*) or mobile or iPod or iPad or android or apple or blackberry or tablet* or (smart NEXT watch*) or computer* or laptop* or PC or microcomputer* or Fitbit or pedometer* or (activity NEXT tracker*) or X-Box or Kinect or Wii or wireless or Nintendo or PlayStation or (gam* NEXT console*) or acceleromet* or "inertial measurement" or IMU or "motion capture" or "personal digital assistant" or (activity NEXT monitor*) or sensor* or "remote sensing technology" or (wearable NEAR/4 (device* or sensor* or technolog*)) or (interactive NEXT technolog*) or "virtual reality" or VR or (virtual NEAR/4 (environment* or object* or system* or program*)) or augmented reality or AR or "mixed reality" or "leap motion" or robot* or orthos* or orthot* or "computer aided" or "computer assisted" or electromechanical or electro-mechanical or exoskeleton* or end-effector or ARMin or IntelliArm or NEUROExos or Mit-Manus or ((electr* or musc* or | 385464 |

| | | |
|---|---|---|
| | neuromusc* or nerve or neuro) NEAR/4 stimulat*) or electrotherapy or "artificial intelligence" or AI or haptic* or "digital interaction" or app? or application? or internet or web-based or online or video or YouTube or skype or zoom or e-mail or "electronic mail" or (text NEXT messag*) or SMS or MMS or Wi-Fi or audio-video or ((tele or video) NEAR/4 conferenc*) or ((serious or applied or health or rehab* or video or interactive or computer) NEAR/4 gam*) or exergam* or gaming):ti,ab,kw | |
| #5 | #1 or #2 or #3 or #4 | 394852 |
| #6 | [mh ^"physical therapy modalities"] or [mh ^"exercise movement techniques"] or [mh "exercise therapy"] or [mh ^exercise] or [mh ^"activities of daily living"] or [mh ^"recreation therapy"] or [mh ^"recovery of function"] or [mh ^"physical and rehabilitation medicine"] or [mh ^"rehabilitation"] or [mh ^"neurological rehabilitation"] or [mh ^"stroke rehabilitation"] | 60273 |
| #7 | [mh ^"allied health occupations"] or [mh ^"occupational therapy"] or [mh ^"allied health personnel"] or [mh ^"physical therapist assistants"] or [mh ^"occupational therapists"] or [mh ^"physical therapists"] | 1712 |
| #8 | (rehab* or neurorehab* or (neurological NEXT rehab*) or (stroke NEXT rehab*) or mobili?ation or (activit* NEAR/4 "daily living") or (functional NEXT activit*) or "functional training" or exercis* or training or physiotherap* or (physical NEXT therap*) or (recreational NEXT therap*) or (occupational NEXT therap*) or "allied health"):ti,ab,kw | 307588 |
| #9 | #6 or #7 or #8 | 310798 |
| #10 | [mh ^"cerebrovascular disorders"] or [mh "basal ganglia cerebrovascular disease"] or [mh "brain ischemia"] or [mh "carotid artery diseases"] or [mh ^"carotid artery, internal, dissection"] or [mh ^"cerebral small vessel diseases"] or [mh "intracranial arterial diseases"] or [mh "cerebral arterial diseases"] or [mh "intracranial embolism and thrombosis"] or [mh ^"intracranial arteriovenous malformations"] or [mh "intracranial hemorrhages"] or [mh "cerebral hemorrhage"] or [mh ^"stroke"] or [mh "brain infarction"] or [mh ^"stroke, lacunar"] or [mh ^"vasospasm, intracranial"] or [mh ^"brain injuries"] or [mh ^"hemiplegia"] or [mh ^"paresis"] or [mh "cerebrovasular trauma"] | 27912 |
| #11 | (stroke or apoplex* or cva or poststroke or post-stroke or ((cerebr* or brain) NEAR/4 vasc*) or hemipar* or hemipleg* or paresis or paretic or ((brain or cerebr* or cerebell* or hemispher* or intracran* or intracerebral or intercran* or vertebrobasilar or "middle cerebral artery" or MCA or "anterior circulation" or "posterior circulation" or "basilar artery" or "vertebral artery" or intraventricular or "basal ganglia" or "internal capsule" or subarchnoid or hemispher*) NEAR/4 (isch?emi* or infarct* or thromb* or embol* or occlus* or hypoxi* or h?emorrhag* or h?ematom* or bleed*))):ti,ab,kw | 93776 |
| #12 | #10 or #11 | 98599 |
| #13 | [mh "upper extremity"] | 10677 |
| #14 | ((upper NEAR/1 (extremit* or limb*)) or arm? or shoulder* or axilla* or hand? or elbow* or forearm* or finger* or wrist* or palm* or thumb*):ti,ab,kw | 264398 |
| #15 | #13 or #14 | 264406 |
| #16 | [mh ^"home environment"] or [mh ^"community health services"] or [mh ^"exp community participation"] or [mh ^"home care services"] or [mh ^"hospital to home transition"] | 3747 |
| #17 | (home or "home based" or community or outreach or "in home" or domiciliary or domicile or domestic or house or household or residence):ti,ab,kw | 129478 |
| #18 | #16 or # 17 | 246691 |
| #19 | [mh "randomized controlled trial"] | 34 |
| #20 | ("controlled clinical trial"):pt | 0 |
| #21 | double-blind or single-blind or randomi?ed or random* or control* or trial* or "pretest posttest design" or "random assignment" or group* or placebo* or "crossover design" or (comparative NEXT stud*) or therapy | 2239930 |

| | | |
|---|---|---|
| #22 | #19 or #20 or #21 | 2239930 |
| #23 | [mh "animals"] not [mh ^"humans"] | 3281 |
| #24 | #22 not #23 | 2236649 |
| #25 | #5 and #9 and #12 and #15 and #18 and #24 | 444 |
| #26 | #25 with Publication Year from 2000 to 2025, with Cochrane Library publication date Between Jan 2000 and Apr 2025, in Trials | 416 |

| g) Scopus (Search completed 29th April 2025) | | |
|---|---|---|
| 1 | TITLE-ABS-KEY ( ( ( digital OR health OR mobile ) W/4 technolog* ) OR e-health OR m-health OR ehealth OR mhealth OR e-therapy OR etherapy OR mtherapy OR m-therapy OR erehab* OR e-rehab* OR mrehab* OR m-rehab* OR "mobile health*" OR "digital health*" OR {digital medicine} OR telemedicine OR tele-medicine OR telehealth* OR tele-health* OR telecare OR tele-care OR telemanagement OR tele-management OR telerehab* OR tele-rehab* OR teleconsult* OR tele-consult* OR telestroke OR tele-stroke OR teletherap* OR tele-therap* OR ( remote W/4 ( rehab* OR therap* OR treatment OR physio* ) ) OR ( video W/4 ( rehab* OR therap* OR treatment OR physio* ) ) OR ( tele W/4 ( rehab* OR therap* OR treatment OR physio* ) ) OR ( virtual W/4 ( rehab* OR therap* OR treatment OR physio* ) ) OR {health informatics} OR {medical informatics} OR ( ( computer OR internet ) W/4 therap* ) OR "biomedical technolog*" OR {patient portal} OR "digital monitor*" OR "digital device" OR "smart phone" OR mobile OR ipod OR ipad OR android OR apple OR blackberry OR tablet OR "smart watch" OR computer OR laptop OR pc OR microcomputer OR fitbit OR pedometer OR "activity tracker" OR x-box OR kinect OR wii OR wireless OR nintendo OR playstation OR "gam* console" OR acceleromet* OR {inertial measurement} OR imu OR {motion capture} OR {personal digital assistant} OR "activity monitor" OR sensor OR {remote sensing technology} OR ( wearable W/4 ( device* OR sensor* OR technolog* ) ) OR "interactive technolog*" OR {virtual reality} OR vr OR ( virtual W/4 ( environment* OR object* OR system* OR program* ) ) OR {augmented reality} OR ar OR {mixed reality} OR {leap motion} OR robot* OR orthos* OR orthot* OR "computer aided" OR "computer assisted" OR electromechanical OR electro-mechanical OR exoskeleton OR end-effector OR armin OR intelliarm OR neuroexos OR mit-manus OR ( ( electr* OR musc* OR neuromusc* OR nerve OR neuro ) W/4 stimulat* ) OR electrotherapy OR {artificial intelligence} OR ai OR haptic* OR {digital interaction} OR app OR application OR internet OR web-based OR online OR video OR youtube OR skype OR zoom OR e-mail OR {electronic mail} OR "text messag*" OR sms OR mms OR wi-fi OR audio-video OR ( ( tele OR video ) W/4 conferenc* ) OR ( ( serious OR applied OR health OR rehab* OR video OR interactive OR computer ) W/4 gam* ) OR exergam* OR gaming ) | 19428635 |
| 2 | TITLE-ABS-KEY ( rehab* OR neurorehab* OR "neurological rehab*" OR "stroke rehab*" OR mobili*ation OR ( activit* W/4 "daily living" ) OR "functional activit*" OR {functional training} OR exercis* OR physiotherap* OR "physical therap*" OR "recreational therap*" OR "occupational therap*" OR {allied health} ) | 1772737 |
| 3 | TITLE-ABS-KEY ( stroke OR apoplex* OR cva OR poststroke OR post-stroke OR ( ( cerebr* OR brain ) W/4 vasc* ) OR hemipar* OR hemipleg* OR paresis OR paretic OR ( ( brain OR cerebr* OR cerebell* OR hemispher* OR intracran* OR intracerebral OR intercran* OR intraventricular OR "basal ganglia" OR "internal capsule" OR subarchnoid OR hemispher* ) W/4 ( ischemi* OR ischaemi* OR infarct* OR thromb* OR embol* OR occlus* OR hypoxi* OR hemorrhag* OR haemorrhag* OR hematom* OR haematom* OR bleed* ) ) ) | 957954 |
| 4 | TITLE-ABS-KEY ( "upper extremit*" OR "upper limb" OR arm OR shoulder OR axilla OR hand OR elbow OR forearm OR finger OR wrist OR palm OR thumb ) | 2759409 |
| 5 | TITLE-ABS-KEY ( home OR "home based" OR community OR outreach OR "in home" OR domiciliary OR domicile OR domestic OR house OR household OR residence ) | 4380280 |

| | | |
|---|---|---|
| 6 | TITLE-ABS-KEY ( double-blind OR single-blind OR randomi*ed OR random* OR control* OR trial* OR {pretest-posttest design} OR {random assignment} OR group* OR placebo* OR {crossover design} OR "comparative study" OR therapy ) | 32257642 |
| 7 | ( TITLE-ABS-KEY ( ( ( digital OR health OR mobile ) W/4 technolog* ) OR e-health OR m-health OR ehealth OR mhealth OR e-therapy OR etherapy OR mtherapy OR m-therapy OR erehab* OR e-rehab* OR mrehab* OR m-rehab* OR "mobile health*" OR "digital health*" OR {digital medicine} OR telemedicine OR tele-medicine OR telehealth* OR tele-health* OR telecare OR tele-care OR telemanagement OR tele-management OR telerehab* OR tele-rehab* OR teleconsult* OR tele-consult* OR telestroke OR tele-stroke OR teletherap* OR tele-therap* OR ( remote W/4 ( rehab* OR therap* OR treatment OR physio* ) ) OR ( video W/4 ( rehab* OR therap* OR treatment OR physio* ) ) OR ( tele W/4 ( rehab* OR therap* OR treatment OR physio* ) ) OR ( virtual W/4 ( rehab* OR therap* OR treatment OR physio* ) ) OR {health informatics} OR {medical informatics} OR ( ( computer OR internet ) W/4 therap* ) OR "biomedical technolog*" OR {patient portal} OR "digital monitor*" OR "digital device" OR "smart phone" OR mobile OR ipod OR ipad OR android OR apple OR blackberry OR tablet OR "smart watch" OR computer OR laptop OR pc OR microcomputer OR fitbit OR pedometer OR "activity tracker" OR x-box OR kinect OR wii OR wireless OR nintendo OR playstation OR "gam* console" OR acceleromet* OR {inertial measurement} OR imu OR {motion capture} OR {personal digital assistant} OR "activity monitor" OR sensor OR {remote sensing technology} OR ( wearable W/4 ( device* OR sensor* OR technolog* ) ) OR "interactive technolog*" OR {virtual reality} OR vr OR ( virtual W/4 ( environment* OR object* OR system* OR program* ) ) OR {augmented reality} OR ar OR {mixed reality} OR {leap motion} OR robot* OR orthos* OR orthot* OR "computer aided" OR "computer assisted" OR electromechanical OR electro-mechanical OR exoskeleton OR end-effector OR armin OR intelliarm OR neuroexos OR mit-manus OR ( ( electr* OR musc* OR neuromusc* OR nerve OR neuro ) W/4 stimulat* ) OR electrotherapy OR {artificial intelligence} OR ai OR haptic* OR {digital interaction} OR app OR application OR internet OR web-based OR online OR video OR youtube OR skype OR zoom OR e-mail OR {electronic mail} OR "text messag*" OR sms OR mms OR wi-fi OR audio-video OR ( ( tele OR video ) W/4 conferenc* ) OR ( ( serious OR applied OR health OR rehab* OR video OR interactive OR computer ) W/4 gam* ) OR exergam* OR gaming ) ) AND ( TITLE-ABS-KEY ( rehab* OR neurorehab* OR "neurological rehab*" OR "stroke rehab*" OR mobili*ation OR ( activit* W/4 "daily living" ) OR "functional activit*" OR {functional training} OR exercis* OR physiotherap* OR "physical therap*" OR "recreational therap*" OR "occupational therap*" OR {allied health} ) ) AND ( TITLE-ABS-KEY ( "upper extremit*" OR "upper limb" OR arm OR shoulder OR axilla OR hand OR elbow OR forearm OR finger OR wrist OR palm OR thumb ) ) AND ( TITLE-ABS-KEY ( stroke OR apoplex* OR cva OR poststroke OR post-stroke OR ( ( cerebr* OR brain ) W/4 vasc* ) OR hemipar* OR hemipleg* OR paresis OR paretic OR ( ( brain OR cerebr* OR cerebell* OR hemispher* OR intracran* OR intracerebral OR intercran* OR intraventricular OR "basal ganglia" OR "internal capsule" OR subarchnoid OR hemispher* ) W/4 ( ischemi* OR ischaemi* OR infarct* OR thromb* OR embol* OR occlus* OR hypoxi* OR hemorrhag* OR haemorrhag* OR hematom* OR haematom* OR bleed* ) ) ) ) ) AND ( TITLE-ABS-KEY ( home OR "home based" OR community OR outreach OR "in home" OR domiciliary OR domicile OR domestic OR house OR household OR residence ) ) AND ( TITLE-ABS-KEY ( double-blind OR single-blind OR randomi*ed OR random* OR control* OR trial* OR {pretest-posttest design} OR {random assignment} OR group* OR placebo* OR {crossover design} OR "comparative study" OR therapy ) ) | 1248 |
| 8 | 7 AND PUBYEAR > 1999 AND PUBYEAR < 2026 | 1238 |
| 9 | 8 AND ( LIMIT-TO ( LANGUAGE , "English" ) ) | 1210 |

| h) ProQuest Health and Medicine Databases (Search completed 28th April 2025) | | |
|---|---|---|
| S1 | (MAINSUBJECT.EXACT("Digital technology") OR MAINSUBJECT.EXACT("Video teleconferencing") OR MAINSUBJECT.EXACT("Smartwatches") OR MAINSUBJECT.EXACT("Handheld computers") OR MAINSUBJECT.EXACT("Personal computers") OR MAINSUBJECT.EXACT("Minicomputers") OR MAINSUBJECT.EXACT("Portable computers") OR MAINSUBJECT.EXACT("Wearable computers") OR MAINSUBJECT.EXACT("Electronic games") OR MAINSUBJECT.EXACT("Computer & video games") OR MAINSUBJECT.EXACT("Robotics") OR MAINSUBJECT.EXACT("Virtual reality") OR MAINSUBJECT.EXACT("Computer simulation") OR MAINSUBJECT.EXACT("Software") OR MAINSUBJECT.EXACT("Neuromuscular electrical stimulation") OR MAINSUBJECT.EXACT("Electric stimulation therapy") OR MAINSUBJECT.EXACT("Telemedicine")) AND stype.exact("Dissertations & Theses") | 778 |
| S2 | noft(((digital OR health OR mobile) NEAR/4 technolog*) OR e-health OR m-health OR ehealth OR mhealth OR e-therapy OR etherapy OR mtherapy OR m-therapy OR erehab* OR e-rehab* OR mrehab* OR m-rehab* OR "mobile health*" OR "digital health*" OR "digital medicine" OR telemedicine OR tele-medicine OR telehealth* OR tele-health* OR telecare OR tele-care OR telemanagement OR tele-management OR telerehab* OR tele-rehab* OR teleconsult* OR tele-consult* OR telestroke OR tele-stroke OR teletherap* OR tele-therap* OR ((computer-based OR internet-based OR "computer assisted") NEAR/4 (therap* OR medicine)) OR ((remote OR electronic OR video OR tele OR virtual) NEAR/4 (consult* OR rehab* OR therap* OR treatment OR physio* OR "occupational therap*" OR monitor* OR care)) OR "health informatics" OR "medical informatics" OR "biomedical technolog*" OR "patient portal" OR "digital monitor*") AND stype.exact("Dissertations & Theses") | 2812 |
| S3 | noft(("digital device*" OR smart-phone* OR "smart phone*" OR mobile OR iPod OR iPad OR android OR apple OR blackberry OR tablet* OR smart-watch* OR "smart watch*" OR computer* OR laptop* OR PC OR microcomputer* OR Fitbit OR pedometer* OR "activity tracker*" OR X-Box OR Kinect OR Wii OR wireless OR Nintendo OR PlayStation OR "gam* console*" OR acceleromet* OR "inertial measurement unit*" OR IMU OR "motion capture" OR "personal digital assistant" OR "activity monitor*" OR sensor* OR "remote sensing technolog*" OR (wearable NEAR/4 (device* OR sensor* OR technolog*)) OR "interactive technolog*" OR "virtual reality" OR VR OR (virtual NEAR/4 (environment* OR object* OR system* OR program*)) OR "augmented reality" OR AR OR "mixed reality" OR "leap motion" OR robot* OR orthos* OR orthot* OR computer-aided OR "computer aided" OR "computer assisted" OR computer-assisted OR electromechanical OR electro-mechanical OR exoskeleton* OR end-effector OR ARMin OR IntelliArm OR NEUROExos OR Mit-Manus OR ((electr* OR musc* OR neuromusc* OR nerve OR neuro) NEAR/4 stimulat*) OR electrotherapy OR "artificial intelligence" OR AI OR haptic* OR "digital interaction" OR app OR application OR internet OR web-based OR online OR video OR YouTube OR skype OR zoom OR e-mail OR "electronic mail" OR "text messag*" OR SMS OR MMS OR Wi-Fi OR audio-video OR ((tele OR video) NEAR/4 conferenc*) OR ((serious OR applied OR health OR rehab* OR video OR interactive OR computer) NEAR/4 gam*) OR exergam* OR gaming)) AND stype.exact("Dissertations & Theses") | 28006 |
| S4 | [S1] OR [S2] OR [S3] | 29373 |
| S5 | (MAINSUBJECT.EXACT("Therapy") OR MAINSUBJECT.EXACT("Physical therapy") OR MAINSUBJECT.EXACT("Occupational therapy") OR MAINSUBJECT.EXACT("Rehabilitation") OR MAINSUBJECT.EXACT("Exercise") OR MAINSUBJECT.EXACT("Activities of daily living")) AND stype.exact("Dissertations & Theses") | 8075 |
| S6 | noft(rehab* OR neurorehab* OR "neurological rehab*" OR "stroke rehab*" OR mobilisation OR (activit* NEAR/4 "daily living") OR "functional activit*" OR "functional training" OR exercis* OR training OR physiotherap* OR "physical therap*" OR "recreational therap*" OR "occupational therap*" OR "allied health | 24130 |

| | | |
|---|---|---|
| | worker*" OR "allied health professional*") AND stype.exact("Dissertations & Theses") | |
| S7 | [S5] OR [S6] | 25388 |
| S8 | (MAINSUBJECT.EXACT("Cerebrovascular disease") OR MAINSUBJECT.EXACT("Stroke")) AND stype.exact("Dissertations & Theses") | 344 |
| S9 | noft(stroke OR apoplex* OR cva OR poststroke OR post-stroke OR ((cerebr* OR brain) NEAR/4 vasc*) OR hemipar* OR hemipleg* OR paresis OR paretic OR ((brain OR cerebr* OR cerebell* OR hemispher* OR intracran* OR intracerebral OR intercran* OR vertebrobasilar OR "middle cerebral artery" OR MCA OR "anterior circulation" OR "posterior circulation" OR "basilar artery" OR "vertebral artery" OR "carotid artery" OR intraventricular OR "basal ganglia" OR "internal capsule" OR subarchnoid OR hemispher*) NEAR/4 (ischemi* OR infarct* OR thromb* OR embol* OR occlus* OR hypoxi* OR hemorrhag* OR hematom* OR bleed*))) AND stype.exact("Dissertations & Theses") | 2051 |
| S10 | [S8] OR [S9] | 2051 |
| S11 | noft("upper limb" OR "upper extremity" OR arm OR shoulder OR axilla OR elbow OR forearm OR wrist OR hand OR finger OR palm OR thumb) AND stype.exact("Dissertations & Theses") | 20430 |
| S12 | (MAINSUBJECT.EXACT("Community health care") OR MAINSUBJECT.EXACT("Home health care")) AND stype.exact("Dissertations & Theses") | 421 |
| S13 | noft(home OR home-based OR community OR outreach OR in-home OR domiciliary OR domicile OR domestic OR house OR household OR residence) AND stype.exact("Dissertations & Theses") | 30250 |
| S14 | [S12] OR [S13] | 30250 |
| S15 | MAINSUBJECT.EXACT("Clinical trials") AND stype.exact("Dissertations & Theses") | 582 |
| S16 | noft(double-blind OR single-blind OR randomised OR random* OR control* OR trial* OR "pretest-posttest design" OR "random assignment" OR group* OR placebo* OR "crossover design" OR "comparative stud*" OR therapy) AND stype.exact("Dissertations & Theses") | 78324 |
| S17 | [S15] OR [S16] | 78324 |
| S18 | [S4] AND [S7] AND [S10] AND [S11] AND [S14] AND [S17] | 26 |

| i) Clinical Trials (Search completed 28th April 2025) | | |
|---|---|---|
| 1 | ("digital technology" OR "health technology" OR "digital therapy" OR "digital rehabilitation" OR teletherapy OR telerehabilitation OR "computer assisted" OR games OR gamification OR "virtual reality" OR "augmented reality" OR application OR computer OR "smart phone" OR robotic OR "electrical stimulation" OR videoconferencing OR "wearable sensors") | 77959 |
| 2 | ("physical therapy" OR physiotherapy OR "occupational therapy" OR "neurological rehabilitation" OR "stroke rehabilitation" OR exercise OR "activities of daily living" OR "functional activity") | 62177 |
| 3 | (stroke OR "cerebrovascular accident" OR "brain infarct" OR "brain ischemia" OR "brain hemorrhage" OR hemiplegia OR "brain injury") | 26961 |
| 4 | ("upper limb" OR "upper extremity" OR arm OR shoulder OR elbow OR wrist OR hand) | 116115 |

| | | |
|---|---|---|
| 5 | (home OR community OR domiciliary) | 45779 |
| 6 | ("randomized trial" OR "controlled trial" OR random OR control OR placebo) | 282927 |
| 7 | ("digital technology" OR "health technology" OR "digital therapy" OR "digital rehabilitation" OR teletherapy OR telerehabilitation OR "computer assisted" OR games OR gamification OR "virtual reality" OR "augmented reality" OR application OR computer OR "smart phone" OR robotic OR "electrical stimulation" OR videoconferencing OR "wearable sensors") AND ("physical therapy" OR physiotherapy OR "occupational therapy" OR "neurological rehabilitation" OR "stroke rehabilitation" OR exercise OR "activities of daily living" OR "functional activity") AND (stroke OR "cerebrovascular accident" OR "brain infarct" OR "brain ischemia" OR "brain hemorrhage" OR hemiplegia OR "brain injury") AND ("upper limb" OR "upper extremity" OR arm OR shoulder OR elbow OR wrist OR hand) AND (home OR community OR domiciliary) AND ("randomized trial" OR "controlled trial" OR random OR control OR placebo) | 347 |

| j) International Clinical Trials Registry Platform (ICTRP) (Search completed 28th April 2025) | | |
|---|---|---|
| 1 | ("digital technology" OR "health technology" OR "digital therapy" OR "digital rehabilitation" OR teletherapy OR tele-therapy OR telerehabilitation OR tele-rehabilitation OR "computer assisted" OR computer-assisted OR games OR gamification OR "virtual reality" OR "augmented reality" OR application OR computer OR "smart phone" OR smart-phone OR robotic OR "electrical stimulation" OR videoconferencing OR video-conferencing OR "wearable sensors") | 43327 |
| 2 | ("physical therapy" OR physiotherapy OR "occupational therapy" OR "neurological rehabilitation" OR "stroke rehabilitation" OR exercise OR "activities of daily living" OR "functional activity") | 43007 |
| 3 | (stroke OR "cerebrovascular accident" OR "brain infarct" OR "brain ischemia" OR "brain ischaemia" OR "brain hemorrhage" OR "brain haemorrhage" OR hemiplegia OR "brain injury") | 23660 |
| 4 | ("upper limb" OR "upper extremity" OR arm OR shoulder OR elbow OR wrist OR hand) | 59689 |
| 5 | (home OR community OR domiciliary) | 24867 |
| 6 | ("randomized trial" OR "randomised trial" OR "controlled trial" OR random* OR control* OR placebo) | 485107 |
| 7 | ("digital technology" OR "health technology" OR "digital therapy" OR "digital rehabilitation" OR teletherapy OR tele-therapy OR telerehabilitation OR tele-rehabilitation OR "computer assisted" OR computer-assisted OR games OR gamification OR "virtual reality" OR "augmented reality" OR application OR computer OR "smart phone" OR smart-phone OR robotic OR "electrical stimulation" OR videoconferencing OR video-conferencing OR "wearable sensors") AND ("physical therapy" OR physiotherapy OR "occupational therapy" OR "neurological rehabilitation" OR "stroke rehabilitation" OR exercise OR "activities of daily living" OR "functional activity") AND (stroke OR "cerebrovascular accident" OR "brain infarct" OR "brain ischemia" OR "brain ischaemia" OR "brain hemorrhage" OR "brain haemorrhage" OR hemiplegia OR "brain injury") AND ("upper limb" OR "upper extremity" OR arm OR shoulder OR elbow OR wrist OR hand) AND (home OR community OR domiciliary) AND ("randomized trial" OR "randomised trial" OR "controlled trial" OR random* OR control* OR placebo) | 28 records for 27 trials |

| | k) ISRCTN -The UK’s Clinical Study Registry (Search completed 28th April 2005) | |
|---|---|---|
| 1 | ("digital technology" OR teletherapy OR telerehabilitation OR "computer assisted" OR games OR gamification OR "virtual reality" OR "augmented reality" OR application OR computer OR "smart phone" OR smart-phone OR robotic OR "electrical stimulation" OR videoconferencing OR "wearable sensors") | 4815 |
| 2 | ("physical therapy" OR physiotherapy OR "occupational therapy" OR rehabilitation OR therapy OR exercise) | 11132 |
| 3 | (stroke OR "cerebrovascular accident" OR "brain infarct" OR "brain ischaemia" OR "brain ischemia" OR "brain hemorrhage" OR "brain haemorrhage") | 2080 |
| 4 | ("upper limb" OR "upper extremity" OR arm) | 6573 |
| 5 | (home OR community) | 7036 |
| 6 | (randomized OR randomised OR controlled) | 23355 |
| 7 | ("digital technology" OR teletherapy OR telerehabilitation OR "computer assisted" OR games OR gamification OR "virtual reality" OR "augmented reality" OR application OR computer OR "smart phone" OR smart-phone OR robotic OR "electrical stimulation" OR videoconferencing OR "wearable sensors") AND ("physical therapy" OR physiotherapy OR "occupational therapy" OR rehabilitation OR therapy OR exercise) AND (stroke OR "cerebrovascular accident" OR "brain infarct" OR "brain ischaemia" OR "brain ischemia" OR "brain hemorrhage" OR "brain haemorrhage") AND ("upper limb" OR "upper extremity" OR arm) AND (home OR community) AND (randomized OR randomised OR controlled) | 54 |

### Table S3. Data collection tool

| Data information | | | | Notes |
|---|---|---|---|---|
| **General Information** | | | | |
| Author | | | | As for APA in text citation |
| Year of publication | | | | |
| Title | | | | |
| Data extractor | | | | Reviewer initials |
| Date of data extraction | | | | |
| Study ID | | | | Trial registration number if available |
| Main, protocol or associated publication | | | | |
| | Notes if associated paper | | | |
| Journal of publication | | | | |
| Hyperlink to publication | | | | |
| Linked publications | Available | | | |
| | If yes provide detail | | | |
| Reference checking and citation searching | Completed | | | |
| | Number of references checked | | | |
| | Number of citations checked | | | |
| | Date citations checked | | | |
| **Study Design and Purpose** | | | | |
| Trial design | Trial stage | | | |
| | If other state | | | |
| | Randomisation design | | | |
| | If other state | | | |
| | Number of groups | | | Number of randomised groups |
| | Other trial features | | | E.g. process evaluation, qualitative study, economic evaluation |
| | Allocation ratio | | | To randomised groups |
| | Number of sites | | | Number of centres/ hospitals/ trusts, geographical regions etc. Please state |
| Aim | | | | State the aim |
| Objectives | | | | State any objectives if provided |
| **Participants** | | | | |

| | | | | |
|---|---|---|---|---|
| Eligibility Criteria | Inclusion criteria | | | |
| | Exclusion criteria | | | |
| PROGRESS factor (included in eligibility) | Location (Place of residence | Part of eligibility | | |
| | | If Yes detail | | |
| | Race, ethnicity, culture, language | Part of eligibility | | |
| | | If Yes detail | | |
| | Occupation | Part of eligibility | | |
| | | If Yes detail | | |
| | Gender, sex | Part of eligibility | | |
| | | If Yes detail | | |
| | Religion | Part of eligibility | | |
| | | If Yes detail | | |
| | Education | Part of eligibility | | |
| | | If Yes detail | | |
| | Socioeconomic status | Part of eligibility | | |
| | | If Yes detail | | |
| | Social capital | Part of eligibility | | Consider support from social relationships and networks. |
| | | If Yes detail | | |
| Personal Characteristics (included in eligibility) | Age | Part of eligibility | | |
| | | If Yes detail | | |
| | Type of stroke | Part of eligibility | | Ischaemic or haemorrhagic |
| | | If Yes detail | | |
| | Time since stroke | Part of eligibility | | |
| | | If Yes detail | | |
| | Previous stroke | Part of eligibility | | |
| | | If Yes detail | | |
| | Post stroke disability | Part of eligibility | | |
| | | If Yes detail | | |
| | Upper limb impairment | Part of eligibility | | Any detail on strength, ROM or impairment outcome scales (linked to ICF domain) |
| | | If Yes detail | | |
| | Upper limb activity | Part of eligibility | | Any detail about functional activity or activity related outcome scales (linked to ICF domain) |
| | | If Yes detail | | |

| | | | | |
|---|---|---|---|---|
| | Cognition | Part of eligibility | | |
| | | If Yes detail | | |
| | Co-morbidities | Part of eligibility | | |
| | | If Yes detail | | |
| | Previous use of technology | Part of eligibility | | |
| | | If Yes detail | | |
| Participant numbers | Total | | | |
| | Intervention | | | |
| | Control | | | |
| PROGRESS Indicators in baseline characteristics | Location (Place of residence | Country | | Only source data to be included with acknowledgement that calculations may be required at a later stage.<br>Exception is if individual participant data is provided then calculations will be made undertaken.<br>Unit of measurement to be recorded e.g. days or months, males or females etc.<br>Several format options for continuous data (mean (SD), median (IQR) and range) or categorical data (number or percentage) will initially be collected with the final format of choice influenced by the available data.<br>For outcome measures, if available MRS for overall disability, FMA-UE for impairment and ARAT for activity. |
| | | If multiple countries provide detail | Intervention | |
| | | | Control | |
| | | Urban/Rural/Inner city | Intervention | |
| | | | Control | |
| | Race, ethnicity, culture, language | Ethnicity | Intervention | |
| | | | Control | |
| | | Language | Intervention | |
| | | | Control | |
| | Occupation | Intervention | | |
| | | Control | | |
| | Gender, sex | Intervention | | |
| | | Control | | |
| | Religion | Intervention | | |
| | | Control | | |
| | Education | Intervention | | |
| | | Control | | |
| | Socioeconomic status | Intervention | | |
| | | Control | | |
| | Social capital | Intervention | | |
| | | Control | | |
| Personal baseline characteristics | Age (in years) | Intervention | | |
| | | Control | | |
| | Type of stroke | Intervention | | |
| | | Control | | |

| | | | | |
|---|---|---|---|---|
| | Time since stroke (in days or months) | Intervention | | |
| | | Control | | |
| | Previous stroke | Intervention | | |
| | | Control | | |
| | Disability | Intervention | | |
| | | Control | | |
| | Upper limb impairment | Intervention | | |
| | | Control | | |
| | Upper limb activity | Intervention | | |
| | | Control | | |
| | Cognition | Intervention | | |
| | | Control | | |
| | Co-morbidities | Intervention | | |
| | | Control | | |
| | Previous use of technology | Intervention | | |
| | | Control | | |
| **Intervention based on TIDieR-Rehab** | | | | |
| Brief name | | | | Name of technology/ intervention (explain acronyms in full) or short statement of intervention. |
| Why | | | | Underpinning theory and mechanisms of impact (or hypothesised mechanisms). Consider active ingredients or components. |
| Who | Severity of upper limb impairment/activity limitation | | | What severity of upper limb impairment/activity limitation was the intervention intended to treat |
| | Upper limb focus | | | Which aspect of upper limb rehabilitation does it aim to influence |
| When | | | | What time since stroke was it intended to treat |
| What materials | Summary of technology | | | |
| | Detail of technology | | | |
| | Technology provided by study | | | Includes hardware and/or software and/or connectivity |
| | Other materials provided | | | Other physical and information materials provided to participants or to intervention providers |
| What procedures | Set up, familiarisation and training session | Took place | | For participants and those supporting the intervention e.g. carers, family, health professionals. |
| | | If Yes detail | | |

| | | | | |
|---|---|---|---|---|
| | During intervention | | | Details of what participants did and any additional follow up / sessions provided. |
| Who provided | Who | | | Who set up and/or delivered the intervention (provider) |
| | If other state | | | |
| | Skills experience and training | | | Pre-existing knowledge and skills, additional training, competence in order to provide intervention (not training in study methods/outcomes etc) |
| How and Where | Set up by provider | Mode of delivery | | |
| | | Where | | |
| | Follow-up by provider | Mode of delivery | | |
| | | Where | | |
| | Self-administered | Which components | | |
| | | Where | | |
| | Supervision/support from others | | | Include family, friends, carers, other professionals not directly providing intervention |
| | Co-interventions | | | Any co-interventions delivered alongside the study intervention including usual care/therapy |
| | Where intervention (excluding set up) took place | | | |
| How much* | Session duration | | | Total time of the intervention session - include units of measurement (minutes, hours) Includes time on task and time not on task. May need to differentiate different parts of intervention e.g. home practice and follow up sessions. |
| | Essential elements amount (minutes and/or repetitions) | | | Total time active/ on task in a single session. This can be reported in time (minutes/hours) or repetitions. This may consist of several specific tasks and the time/repetitions of each individual task should be recorded |
| | Frequency | | | Number of sessions per day and/or week, over the intervention length |
| | Intervention length (duration) | | | Total length of intervention including units of measurement (weeks, months) |
| How challenging* | Difficulty | | | How hard the task is. Includes the absolute challenge (difficulty based on task), relative challenge (difficulty |

| | | | | |
|---|---|---|---|---|
| | | | | of task relative to patients' ability) or the perceived challenge of each task(subjective experience of task) |
| | Intensity | | | How much work (or work per time) is performed in each session. Work e.g. repetitions or work per time e.g. repetitions per minute for each task |
| Regression/progression | | | | Of the dose parameters, how much and how challenging. Include how these decisions are made if reported. |
| Personalisation | Needs | | | Provision of supplementary strategies, resources or expertise to enable the delivery of essential elements of the intervention |
| | Preferences | | | Person-centred adaptions to the intervention to promote experience, engagement and adherence |
| Protocol deviations | | | | |
| How well<br>Note assessment of fidelity covered in outcomes and results section | Strategies to maintain/improve fidelity | | | |
| Harms<br>Note assessment of fidelity covered in outcomes and results section | | | | |
| Control group | What | | | Brief description of control |
| | Dose matched | | | |
|  | Additional detail on dose matching |  |  | For example, if intervention was dose matched but not usual care. |
| **Outcomes** | | | | |
| Primary Outcome | | | | |
| Impairment of motor function measurement tools | | | | List all tools used to measure domain |
| Activity limitation measurement tools | | | | List all tools used to measure domain |
| Adverse event/harm measurement tools | | | | List all tools used to measure domain such as tools to measure spasticity, fatigue, pain etc. |
| Timing of outcome measures | Baseline (always 0) | | | Report time units i.e., weeks/months |
| | Midpoint | | | |

| | | | | |
|---|---|---|---|---|
| | Post Intervention | | | |
| | Follow up 1 | | | |
| | Follow up 2 | | | |
| Adverse event/harm– non-systematic collection | | | | This is adverse event reporting. |
| Fidelity | Was fidelity assessed | | | |
| | If yes, how was it assessed | | | |
| Economic evaluation | Was an economic evaluation made | | | |
| | Type of evaluation | | | Cost effectiveness, cost utility, cost benefit or cost consequence. |
| | Measurement of cost and perspective of cost | | | |
| | Measurement of benefit | | | |
| Other outcomes | | | | List other outcome measurement tools used. |
| **Results$** | | | | |
| Type of analysis | Type | | | |
| | If other state | | | |
| | Management of missing data | | | State how missing data managed |
| For each impairment, activity and adverse event tool | Tool | | | State tool from which data extracted. Use overall tool if total and subscale scores are reported. If only subscales are reported, report each subscale separately. If a total score and subscale provide information about a different impairment/activity domain then report. Note range of movement or muscle strength for individual upper limb joints/ muscle groups will not be extracted however grip strength will be extracted. |
| | Units of measurement if appropriate | | | |
| | Baseline score | Number of participants | Intervention | |
| | | | Control | |
| | | Intervention | Mean/Median/N | Choose mean over median at all times if available. |
| | | | SD/IQR/% | |
| | | Control | Mean/Median/N | |
| | | | SD/IQR/% | |

| | | | | |
|---|---|---|---|---|
| | | Notes | | |
| | Post score | Number of participants | Intervention | |
| | | | Control | |
| | | Intervention | Mean/Median/N | |
| | | | SD/IQR/% | |
| | | Control | Mean/Median/N | |
| | | | SD/IQR/% | |
| | | Notes | | |
| | Change score to post | Intervention | Change | Include any detail provided about change including mean(SD) etc and any statistical tests and p values |
| | | Control | Change | |
| | Group difference post | Measurement used | | What is being compared between groups e.g. change score or outcome score |
| | | Difference | | |
| | | Statistical analysis | | |
| | Follow up score | Number of participants | Intervention | |
| | | | Control | |
| | | Intervention | Mean/Median/N | |
| | | | SD/IQR/% | |
| | | Control | Mean/Median/N | |
| | | | SD/IQR/% | |
| | | Notes | | |
| | Change score to follow-up | Time period of change | | |
| | | Intervention | Change | As above |
| | | Control | Change | |
| | Group difference to follow-up | Measurement used | | As above |
| | | Difference | | Include any detail provided |
| | | Statistical analysis | | |
| Adverse events | Total | | | |
| | Adverse events | | | |
| | Serious adverse events | | | |
| | Detail if available | | | |
| Fidelity | Measures assessed | | | |
| | Participant numbers in analysis | | | For intervention and control as appropriate |
| | Intervention | | | Provide any detail of measures assessed |

| | | | | |
|---|---|---|---|---|
| | Control | | | |
| Economic evaluation | Participant numbers in analysis | | | Give both groups |
| | Intervention | Cost total | | Indicate units |
| | | Economic outcome | | Indicate time period if given |
| | Control | Cost total | | |
| | | Economic outcome | | |
| | Cost difference | | | Include any detail provided including statistical tests and p values |
| **Other** | | | | |
| Key conclusions | | | | |
| Notes | | | | |

**Abbreviations**

ARAT: Action Research Arm Test, ICF: International Classification of Functioning, Disability and Health[1], IRQ: Interquartile Range, FMA-UE: Fugl-Meyer Assessment for Upper Extremity, MRS: Modified Rankin Scale, ROM: Range of Motion, SD: Standard Deviation

TIDieR-Rehab: Template for Intervention Description and Replication: Extension for Rehabilitation[2,3]

**Notes**

*The TIDieR-Rehab elements of "how much" and "how challenging" influence the dose of the intervention. The items under this section are informed by bringing information together from the TIDieR-Rehab checklist and the Dose articulation framework [4]

[$] If there is more than one control group i.e. a sham and control group, or more than one follow-up time results from both groups/time periods will be reported

Table S4. Summary of the participant characteristics of included studies

| Study | Number n | Gender Female n (%) | Age (years) Mean (SD) *Median (IQR) | Type of stroke Ischaemic n(%) | Time since stroke Mean (SD) *Median (IQR (range when stated)) | Impairment in upper limb function Mean (SD) *Median (IQR) | Limitation in upper limb activity Mean (SD (CI when stated)) *Median (IQR) | Cognition Mean (SD) *Median (IQR) |
|---|---|---|---|---|---|---|---|---|
| Adie [5] | 235<br>E=117<br>C=118 | E=51 (44%)<br>C=53 (45%) | E=66.8(14.6)<br>C=68.0(11.9) | E=104 (89%)<br>C=105(89%) | Days<br>E=57.3(48.3)<br>C=56.3(50.1) | Not reported | ARAT<br>E=41.2(15.9)<br>C=41.0(16.6) | MOCA<br>E= 23.8(4.5)<br>C=23.5(5.2) |
| Alon [6] | 15<br>E=7<br>C=8 | E=4(57%)<br>C=37.5% | Not reported | Not reported | Days<br>E=18.0 (6.2)<br>C=15.6(3.9) | modified FMA-UE<br>E= 23.9 (7.4)<br>C=21.9 (7.5) | BBT<br>E=5.9(6.0)<br>C=5.3(6.2) | MMSE<br>E=27.4(1.9)<br>C=26.5(3.0) |
| Butcher [7] | 24<br>E=16<br>C=8 | E=13(81%)<br>C=4 (50%) | E=66.5 (15)<br>C=64.6(13.6) | E=10 (62.5%)<br>E=5 (62.5%) | Days<br>E=17 (13.9)<br>C=16(6.7) | FMA-UE<br>E= 36.1 (18.1)<br>C= 41.5(18.6) | ARAT<br>E=29.5 (21.0)<br>C=36.1(22.7) | Not reported |
| Cramer [8] | 124<br>E=62<br>C=62 | E=14 (23%)<br>C=20 (32%) | E=62(14)<br>C=60(13) | E=54(87.1%)<br>C=52(83.9%) | Days<br>E=132(65)<br>C=129(59) | FMA-UE<br>E= 42.8(7.8)<br>C= 42.7(8.7) | BBT<br>E=21.3(13.3)<br>C=23.8(12.7) | MOCA<br>E= 24.9(4.1)<br>C=24.4(5.0) |
| Da-Silva [9] | 33<br>E=14<br>C=19 | E=8 (57%)<br>C=12 (63%) | *E=73 (65–80)<br>*C=69 (61–80) | E=13 (93%)<br>C=18(95%) | Days<br>*E=27 (13–48)<br>*C=26 (18–33) | MI<br>*E= 77 (54–84)<br>*C= 51 (38–70) | ARAT<br>*E=37(16-46)<br>*C=15(2-35) | Not reported |
| Emmerson [10] | 62<br>E=30<br>C=32 | E=13 (43%)<br>C=13 (41%) | E=68 (15)<br>C=63(18) | E=22 (79%)<br>C=27 (90%) | Days<br>*E=122 (77–193)<br>*C=133(58-228) | Grip strength (kg)<br>E=10.5(11.0)<br>C=10.8(10.4) | WMFT(time in secs)<br>E=39(44)<br>C=49(47) | Not reported |
| Fletcher-Smith [11] | 40<br>E=20<br>C=20 | E=13 (65%)<br>C=7 (35%) | *E=75(66.5-80)<br>*E=67(59-84) | E=17(85%)<br>C=18(90%) | Days<br>*E=1.5 (range:0-3)<br>*C=2 (range: 1-3) | NIHSS arm<br>*E=3(2-4)<br>*C=4(2-4) | ARAT<br>*E=0 (0-6)<br>*C=0(0-5.5) | MOCA:<br>*E=19.5(16-23)<br>*C=22(19-27) |
| Standen [12] | 27<br>E=17<br>C=10 | E=9 (53%)<br>C=2 (20%) | E=59(12.03)<br>C=63(14.06) | Not reported | Weeks<br>*E=22(16-59.5).<br>*C=12(7.75-20.25). | Not reported | WMFT (time in seconds)<br>*E=2.60(1.65-6.00)<br>*C=3.34(1.90-4.92) | Not reported |

| | | | | | | | | |
|---|---|---|---|---|---|---|---|---|
| Swanson [13] | 27<br>E=14<br>C=13 | E=0 (0%)<br>C=4 (31%) | E=50.3(10.9)<br>C=52(8.7) | E=11 (79%)<br>C=9 (69%) | Weeks<br>E=9.7(4.5)<br>C=10.1(5.1) | FMA-UE<br>E= 36.7 (15.4)<br>C= 35.18 (14.5) | BBT<br>E=25.4 (17.6)<br>C=23.5(14.8) | Not reported |
| Wei [14] | 84<br>E=32<br>C1=25<br>C2=27 | E=7 (22%)<br>C1=5(20%)<br>C2=4 (15%) | E=59.19(11.25)<br>C1=60.44(10.38)<br>C2=63.11(10.27) | E=21(66%)<br>C1=21(84%)<br>C2=20(74%) | Days<br>E=47.75(21.93)<br>C1=61.08(41.26)<br>C2=53.67(41.16) | FMA-UE<br>E=52.03(13.95)<br>C1=51.24(16.74)<br>C2=50.78(17.59) | ARAT<br>E=34.00(17.84)<br>C1=36.12(21.84)<br>C2=35.96(22.59) | MMSE<br>E=26.33(3.31)<br>C1=26.87(3.20)<br>C2=27.17(3.00) |
| Wilson [15] | 17<br>E=10<br>C=7 | E=3 (30%)<br>C=2 (29%) | E=69.9 (13.8)<br>C=77.3 (8.9) | E=9(90%)<br>C=5(71%) | Days<br>E=137.5 (152.4)<br>C=107.4 (56.4) | Not reported | BBT<br>E=23.5 (9.6)<br>C=15.7 (10.8) | MOCA<br>E=18.5 (5.1)<br>C=17.0 (7.8) |
| Wolf [16] | 99<br>E=51<br>C=48 | E=26 (51%)<br>C=17(35%) | E=59.1(14.1)<br>C=54.7(12.2) | Not reported | Days<br>E=115.5(53.1)<br>C=127.1(46.2) | FMA-UE<br>E= 34.1(12.1)<br>C= 33.3(12.0) | ARAT<br>E=34.4 (95%CI: 24.7, 44.0)<br>C=31.1 (95%CI: 22.1,40.1) | Not reported |

**Abbreviations**

ARAT: Action Research Arm Test, BBT: Box and Block Test, C: Comparator group, CI: Confidence Interval, E: Experimental group, FMA-UE: Fugl-Meyer Assessment for Upper Extremity, IQR: Interquartile Range, MI: Motricity Index, MMSE: Mini Mental State Examination, MOCA: Montreal Cognitive Assessment, n: Number, NIHSS: National Institute of Health Stroke Scale, SD: Standard Deviation, WMFT: Wolf motor function test

**Notes**

*median and IQR reported

### Table S5. PROGRESS factors reporting

| Factor | | | Reporting | |
|---|---|---|---|---|
| | | Sub group | n(%) | References |
| Progress factor | Location (Place of residence) | N/A | 1 (8%) | [14] |
| | Race, ethnicity, culture, language | Ethnicity | 4 (33%) | [7,8,13,16] |
| | | Language | 0 (0%) | N/A |
| | Occupation/ Employment | N/A | 1 (8%) | [5] |
| | Gender, sex | N/A | 12 (100%) | [5–16] |
| | Religion | N/A | 0 (0%) | N/A |
| | Education | N/A | 1 (8%) | [14] |
| | Socioeconomic status | N/A | 0 (0%) | N/A |
| | Social capital | N/A | 0 (0%) | N/A |

Table S6 Study data included in meta-analysis

a) comparing the effects of virtual reality and conventional exercise programmes on motor impairment using the Fugl-Meyer Assessment for Upper Extremity

| Study | Experimental | | | Comparator | | |
|---|---|---|---|---|---|---|
| | Mean | SD | Sample | Mean | SD | Sample |
| Cramer[8] | 7.86* | 6.68* | 62 | 8.36* | 7.04* | 62 |
| Swanson[13] | 43.2 | 16.3 | 14 | 36.64 | 14.8 | 11 |
| Wolf[16] | 43.4 | 42.91† | 47 | 42.9 | 42.4† | 45 |

b) comparing the effects of virtual reality and conventional exercise programmes on combined measures of activity limitation (Action Research Arm Test and Box and Block Test)

| Study | Measurement tool | Experimental | | | Comparator | | |
|---|---|---|---|---|---|---|---|
| | | Mean | SD | Sample | Mean | SD | Sample |
| Adie[5] | ARAT | 47.6 | 14.2 | 101 | 49 | 13.6 | 108 |
| Cramer[8] | BBT | 30.8 | 13.3 | 62 | 32.6 | 15.4 | 62 |
| Swanson[13] | BBT | 28.9 | 17.6 | 14 | 24.5 | 15.8 | 11 |
| Wilson[15] | BBT | 34.7 | 13.8 | 10 | 16 | 9.4 | 7 |
| Wolf[16] | ARAT | 39.5 | 37.81† | 47 | 39.9 | 38.61† | 45 |

**Abbreviations**

ARAT: Action Research Arm Test, BBT: Box and Block Test, SD: Standard deviation

**Notes**

*change from baseline data (post intervention measurement data not available and acceptable to use change from baseline as combining data using mean difference [17]

†calculated from confidence interval

### Table S7. GRADE evidence table

| Virtual reality compared to conventional exercise therapy for upper limb rehabilitation at home in the first acute and subacute phases after stroke | | | | | | | |
|---|---|---|---|---|---|---|---|
| Population: Adult participants with upper limb deficit as a result of a stroke within the previous six months | | | | | | | |
| Setting: Experimental intervention, targeting the upper limb, delivered in the home setting | | | | | | | |
| Experimental intervention: Virtual reality | | | | | | | |
| Comparator Intervention: Conventional exercise therapy | | | | | | | |
| **Outcome** | **Design** | **Risk of bias** | **Inconsistency** | **Indirectness** | **Imprecision** | **Publication bias (see also supplementary figure S2)** | **Overall judgement** |
| Post intervention impairment of motor function measured by FMA-UE | RCT | Yes downgrade one level<br>Reason:<br>One or more risk of bias domains were judged as unclear in all included studies | Yes<br>Reasons:<br>Although no heterogeneity indicated by $I^2$, small study numbers may compromise the reliability of estimate<br>Point estimate of individual studies and pooled studies didn't meet MCID but upper boundaries of CIs of 2 of 3 studies met the MCID. Raises doubts about finding of no effect. | No<br>Reason:<br>No indirectness associated with PICO | No<br>Reason:<br>Small CI and MCID does not fall within CI | No<br>Reason:<br>Limited information from funnel plot due to small study numbers | Low |
| Post intervention limitation of activity measured by either the ARAT or BBT | RCT | Yes downgrade one level<br>Reason:<br>One or more risk of bias domains were judged as unclear in all included studies | Yes downgrade one level<br>Reason: Heterogeneity was indicated by $I^2$<br>Point estimate of 4 of 5 individual studies and pooled studies didn't meet MCID but point estimate of one study met the MCID and the upper boundaries of CIs of 2 of 5 studies also met the MCID. Raises doubts about finding of no effect. | No<br>Reason:<br>No indirectness associated with PICO | No<br>Reason:<br>Small CI and MCID does not fall within CI | No<br>Reason:<br>Limited information from funnel plot due to small study numbers | Low |

**Abbreviations**

ARAT: Action Research Arm Test, BBT: Box and Block Test, CI: Confidence Interval, FMA-UE: Fugl-Meyer Assessment for Upper Extremity, MCID: Minimal Clinically Important Difference, PICO: Population, Intervention, Comparator, Outcome (criteria), RCT: Randomised Controlled Trial

### Table S8. Harms and adverse events

| Study | Harms | | Adverse events | | | |
|---|---|---|---|---|---|---|
| | Outcome measures | Summary of outcomes | Reporting | Adverse events n | Serious adverse events n | Relation to study interventions |
| Adie [5] | None | N/A | Participant record in daily diary | Not reported | 46 | None related |
| Alon [6] | None | N/A | Recorded but no detail on method | 0 | 0 | N/A |
| Butcher [7] | Fatigue: FSS-7<br>Spasticity: MMAS<br>Arm pain: Yes/No and VAP | No significant differences reported between groups in fatigue, spasticity or pain prevalence<br>VAS pain data reported but not analysed | Therapist record post intervention | E= 5<br>C=0 | E=1<br>C=2 | 5 AEs in intervention definitely (1 finger redness), probably (1 carpel tunnel symptoms) and possibly (3 pain and migraine) related<br>1 SAE in intervention unlikely related<br>2 SAEs in comparator unrelated<br>No significant difference between groups in the number of AEs |
| Cramer [8] | None | N/A | Therapist record every 3rd supervised session. | E=10<br>C=7 | E=1<br>C=6 | All SAEs unrelated. 6 AEs in intervention (arm and shoulder pain) and 5 in comparator (fatigue and arm and shoulder pain) reasonably or definitely related |
| Da-Silva [9] | Arm pain: NRS.<br>Fatigue: NRS | Group specific pain and fatigue data reported but not analysed | Therapist record twice weekly at review sessions (only SAEs formally recorded) | Not reported | 8 | None related |
| Emmerson [10] | None | N/A | Recorded but no detail on method | 0 | 0 | N/A |
| Fletcher-Smith [11] | Pain: SPIN.<br>Spasticity: stretch induced activation of muscles | Group specific pain data reported but not analysed.<br>Spasticity reported as data combined for both groups and not analysed. | Not recorded | N/A | N/A | N/A |
| Standen [12] | None | N/A | Not recorded | N/A | N/A | N/A |
| Swanson [13] | Spasticity: MAS.<br>Arm pain: VAP | No significant differences reported between groups in spasticity or pain<br>In the intervention group no significant changes was found | Recorded but no detail on method | 0 | 0 | N/A |

| | | | | | | |
|---|---|---|---|---|---|---|
| | | from baseline to post intervention | | | | |
| Wei [14] | None | N/A | Therapist record at weekly telephone call (experimental and sham groups) | 0 | 0 | N/A |
| Wilson [15] | None | N/A | Recorded but no detail on method | 0 | 0 | N/A |
| Wolf [16] | Spasticity : MAS (identified in protocol) | No published data | Therapist record at weekly telephone call / email contact | E=6<br>C= 5 | 0 | Not reported |

**Abbreviations**

AE: Adverse Event, C: Comparator, E: Experimental, FFS-7: Fatigue Severity Scale, MAS: Modified Ashworth scale , MMAS: Modified, Modified Ashworth Scale, NRS: Numeric Rating Scale, SAE: Serious Adverse Event, SPIN: Scale of Pain Intensity, VAP: Visual Analog Pain

### Table S9: Economic Evaluation

| Study | Economic evaluation | |
|---|---|---|
| | Completed | Summary |
| Adie [5] | Cost (health and social care perspective) per quality-adjusted life year | High probability estimate that the Wii™ was more expensive and less effective than usual care. |
| Alon [6] | No | N/A |
| Butcher [7] | Cost (participant, health and social care perspective) per quality- adjusted life year | Not reported |
| Cramer [8] | No | N/A |
| Da-Silva [9] | No | N/A |
| Emmerson [10] | No | N/A |
| Fletcher-Smith [11] | Cost (health and societal perspective) per quality- adjusted life year | Uncertainty on all cost-effectiveness. Data indicates that electrical stimulation had higher outcomes and lower costs than usual care. |
| Standen [12] | No | N/A |
| Swanson [13] | No | N/A |
| Wei [14] | No | N/A |
| Wilson [15] | No | N/A |
| Wolf [16] | No | N/A |

### Figure S1. RoB 2 assessment for meta-analyses

a) RoB 2 for studies comparing the effects of interactive technologies and conventional exercise programmes on motor impairment using the Fugl-Meyer Assessment for Upper Extremity

| Study | Technology group | Outcome | D1 | D2 | D3 | D4 | D5 | Overall |
|---|---|---|---|---|---|---|---|---|
| Cramer | Virtual reality | FMA-UE | + | + | + | + | ! | ! |
| Swanson | Virtual reality | FMA-UE | ! | + | + | + | ! | ! |
| Wolf | Virtual reality | FMA-UE | + | ! | + | + | + | ! |

+ Low risk
! Some concerns
- High risk

D1 Randomisation process
D2 Deviations from the intended interventions
D3 Missing outcome data
D4 Measurement of the outcome
D5 Selection of the reported result

b) RoB 2 for studies comparing the effects of interactive technologies and conventional exercise programmes on combined measures of activity limitation (Action Research Arm Test and Box and Block Test)

| Study | Technology group | Outcome | D1 | D2 | D3 | D4 | D5 | Overall |
|---|---|---|---|---|---|---|---|---|
| Adie | Virtual reality | ARAT | + | + | + | ! | + | ! |
| Cramer | Virtual reality | BBT | + | + | + | + | ! | ! |
| Swanson | Virtual reality | BBT | ! | + | + | + | ! | ! |
| Wilson | Virtual reality | BBT | + | + | + | + | ! | ! |
| Wolf | Virtual reality | ARAT | + | ! | + | + | + | ! |

+ Low risk
! Some concerns
- High risk

D1 Randomisation process
D2 Deviations from the intended interventions
D3 Missing outcome data
D4 Measurement of the outcome
D5 Selection of the reported result

Figure S2 . Funnel plot for meta-analyses

a) Funnel plot for studies comparing the effects of interactive technologies and conventional exercise programmes on motor impairment using the Fugl-Meyer Assessment for Upper Extremity

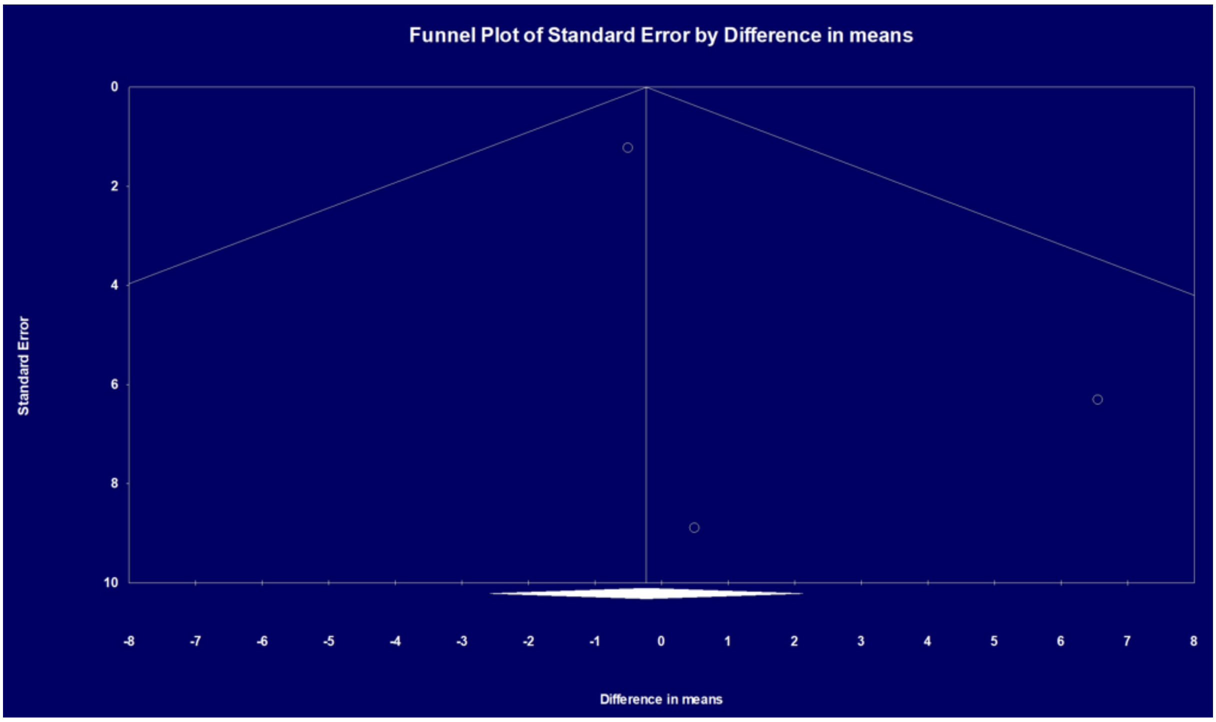

b) Funnel plot for studies comparing the effects of interactive technologies and conventional exercise programmes on combined measures of activity limitation (Action Research Arm Test and Box and Block Test)

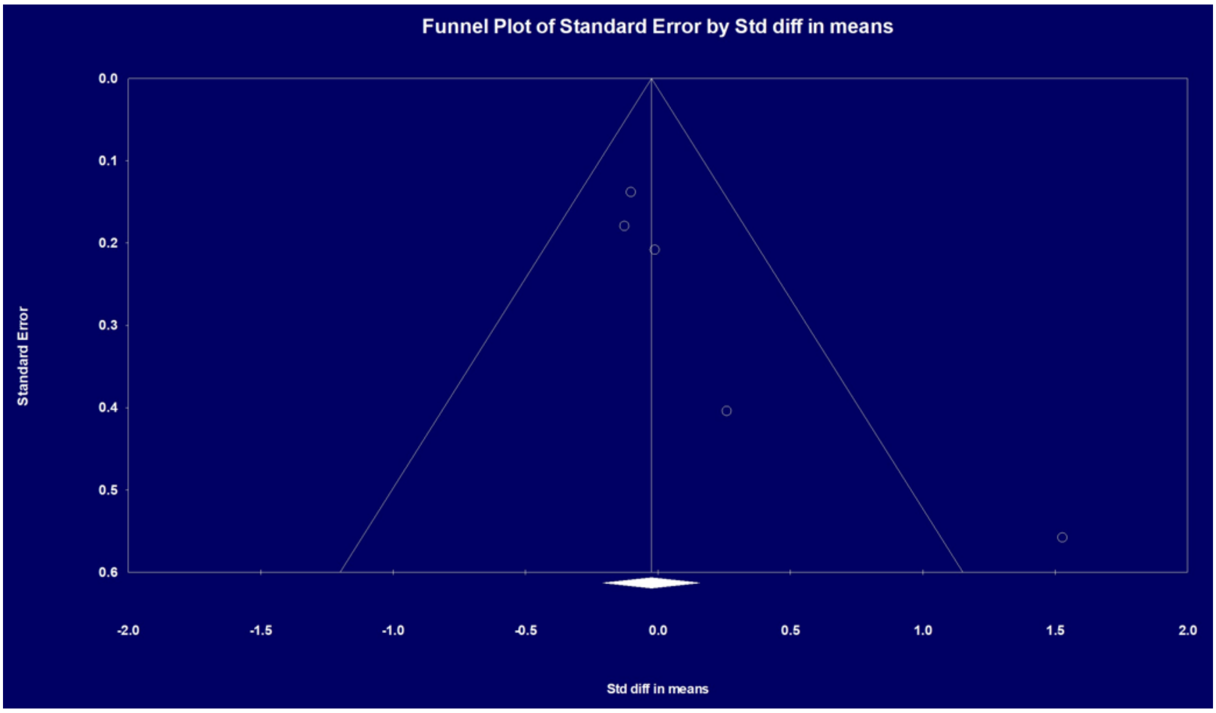

### References

1. World Health Organisation. *International classification of functioning, disability and health (ICF)*. Geneva: World Health Organisation, 2001 Available from www.who.int/standards/classifications/international-classification-of-functioning-disability-and-health.

2. Hoffmann TC, Glasziou PP, Boutron I, et al. Better reporting of interventions: template for intervention description and replication (TIDieR) checklist and guide. *BMJ* 2014; 348: g1687.

3. Signal N, Gomes E, Olsen S, et al. Enhancing the reporting quality of rehabilitation interventions through an extension of the Template for Intervention Description and Replication (TIDieR): the TIDieR-Rehab checklist and supplementary manual. *BMJ Open* 2024; 14: e084320.

4. Hayward KS, Churilov L, Dalton EJ, et al. Advancing stroke recovery through improved articulation of nonpharmacological intervention dose. *Stroke* 2021; 52: 761–769.

5. Adie K, Schofield C, Berrow M, et al. Does the use of Nintendo Wii Sports™ improve arm function? Trial of Wii™ in Stroke: a randomized controlled trial and economics analysis. *Clin Rehabil* 2017; 31: 173–185.

6. Alon G, Levitt AF and McCarthy PA. Functional electrical stimulation enhancement of upper extremity functional recovery during stroke rehabilitation: a pilot study. *Neurorehabil Neural Repair* 2007; 21: 207–215.

7. Butcher T, Warland A, Stewart V, et al. Rehabilitation using virtual gaming for Hospital and hOMe-Based training for the Upper limb in acute and subacute Stroke (RHOMBUS II): results of a feasibility randomised controlled trial. *BMJ Open* 2025; 15: e089672.

8. Cramer SC, Dodakian L, Le V, et al. Efficacy of home-based telerehabilitation vs in-clinic therapy for adults after stroke: a randomized clinical trial. *JAMA Neurol* 2019; 76: 1079–1087.

9. Da-Silva RH, Moore SA, Rodgers H, et al. Wristband Accelerometers to motiVate arm Exercises after Stroke (WAVES): a pilot randomized controlled trial. *Clin Rehabil* 2019; 33: 1391–1403.

10. Emmerson KB, Harding KE and Taylor NF. Home exercise programmes supported by video and automated reminders compared with standard paper-based home exercise programmes in patients with stroke: a randomized controlled trial. *Clin Rehabil* 2017; 31: 1068–1077.

11. Fletcher-Smith JC, Walker D-M, Allatt K, et al. The ESCAPS study: a feasibility randomized controlled trial of early electrical stimulation to the wrist extensors and flexors to prevent post-stroke complications of pain and contractures in the paretic arm. *Clin Rehabil* 2019; 33: 1919–1930.

12. Standen P, Threapleton K, Richardson A, et al. A low cost virtual reality system for home based rehabilitation of the arm following stroke: a randomised controlled feasibility trial. *Clin Rehabil* 2017; 31: 340–350.

13. Swanson VA, Johnson C, Zondervan DK, et al. Optimized home rehabilitation technology reduces upper extremity impairment compared to a conventional home exercise program: a randomized, controlled, single-blind trial in subacute stroke. *Neurorehabil Neural Repair* 2023; 37: 53–65.

14. Wei WXJ, Fong KNK, Chung RCK, et al. "Remind-to-Move" for promoting upper extremity recovery using wearable devices in subacute stroke: a multi-center randomized controlled study. *IEEE Trans Neural Syst Rehabil Eng* 2019; 27: 51–59.

15. Wilson PH, Rogers JM, Vogel K, et al. Home-based (virtual) rehabilitation improves motor and cognitive function for stroke patients: a randomized controlled trial of the Elements (EDNA-22) system. *J NeuroEngineering Rehabil* 2021; 18: 165.

16. Wolf SL, Sahu K, Bay RC, et al. The HAAPI (Home Arm Assistance Progression Initiative) trial: a novel robotics delivery approach in stroke rehabilitation. *Neurorehabil Neural Repair* 2015; 29: 958–968.

17. Deeks JJ, Higgins JPT, Altman DG, et al. Chapter 10: Analysing data and undertaking meta-analyses [last updated November 2024]. In: Higgins JPT, Thomas J, Chandler J, et al. (eds). *Cochrane handbook for systematic reviews of interventions.* Version 6.5 (updated August 2024). Cochrane, 2024. Available from www.cochrane.org/authors/handbooks-and-manuals/handbook/current.
